# Effect of chronic sedative-hypnotic use on sleep architecture and brain oscillations in older adults with chronic insomnia

**DOI:** 10.1101/2024.09.12.24313583

**Authors:** Loïc Barbaux, Aurore A. Perrault, Nathan E. Cross, Oren M. Weiner, Mehdi Essounni, Florence B. Pomares, Lukia Tarelli, Margaret McCarthy, Antonia Maltezos, Dylan Smith, Kirsten Gong, Jordan O’Byrne, Victoria Yue, Caroline Desrosiers, Doris Clerc, Francis Andriamampionona, David Lussier, Suzanne Gilbert, Cara Tannenbaum, Jean-Philippe Gouin, Thien Thanh Dang-Vu

## Abstract

**Rationale:** High rates of insomnia in older adults lead to widespread benzodiazepine (BZD) and benzodiazepine receptor agonist (BZRA) use, even though chronic use has been shown to disrupt sleep regulation and impact cognition. Little is known about sedative-hypnotic effects on NREM slow oscillations (SO) and spindles, including their coupling, which is crucial for memory, especially in the elderly.

**Objectives:** Our objective was to investigate the effect of chronic sedative-hypnotic use on sleep macro-architecture, EEG relative power, as well as SO and spindle characteristics and coupling.

**Methods:** One hundred and one individuals (66.05 ± 5.84 years, 73% female) completed a one-night study and were categorized into three groups: good sleepers (GS, n=28), individuals with insomnia (INS, n=26) or individuals with insomnia who chronically use either BZD or BZRA to manage their insomnia difficulties (MED, n=47; dose equivalent in Diazepam: 6.1 ± 3.8 mg/week). We performed a comprehensive comparison of sleep architecture, EEG relative spectrum, and associated brain oscillatory activities, focusing on NREM brain oscillations crucial for sleep-dependent memory consolidation (i.e., SO and spindles) and their temporal coupling.

**Results:** Chronic use of BZD/BZRA worsened sleep architecture and spectral activity compared to older adults with and without insomnia disorder. The use of BZD/BZRAs also altered the characteristics of sleep-related brain oscillations and their synchrony. An exploratory interaction model suggested that BZD use exacerbated sleep alterations compared to BZRA, and higher BZD/BZRA dosage worsened alteration in sleep micro-architecture and EEG spectrum.

**Conclusions:** Our results suggest that chronic use of sedative-hypnotics is detrimental to sleep when compared to drug-free GS and INS. Such alteration of sleep regulation – at the macro and micro-architectural levels - may contribute to the reported association between sedative-hypnotic use and cognitive impairment in older adults.

**STATEMENT OF SIGNIFICANCE:** Widespread use of sedative-hypnotics is driven by high insomnia rates among older adults. Chronic use can disrupt sleep and cognitive function, however, its impact on sleep regulation – at the macro and micro-architecture levels - is not well understood. We assessed the effect of chronic sedative-hypnotic use in older adults using a between-group design involving good sleepers, individuals with insomnia disorder who do not take any pharmacological treatment to manage their symptoms and individuals with insomnia disorder who chronically use sedative-hypnotics as a sleep aid. We performed a comprehensive comparison of sleep architecture, EEG relative spectrum, and associated NREM brain oscillations crucial for sleep-dependent memory consolidation (i.e., SO and spindles) and their temporal coupling. We showed that chronic use of sedative-hypnotics is detrimental to sleep regulation – at the macro and micro level - compared to drug-free GS and INS, and this may contribute to the reported link between sedative-hypnotic use and cognitive impairment in older adults.

## INTRODUCTION

Insomnia complaints are one of the most common sleep disturbances in the general population^1^ and exhibit a higher prevalence among older individuals compared to young adults^2,3^. Insomnia disorder is defined by complaints of difficulties initiating and/or maintaining sleep, despite having adequate opportunities and circumstances for sleep, occurring at least three times per week for more than three months^4^. Importantly, a diagnosis of insomnia disorder also requires complaints about daytime functioning, such as fatigue, impaired concentration and mood disruption^5,6^. Insomnia disorder not only impairs quality of life and health but also increases the risk for cognitive decline and dementia^7–11^, thereby representing a major health issue in the aging population.

While cognitive-behavioural therapy for insomnia (CBTi), a multimodal psychological intervention, is considered the gold standard management of insomnia, its access remains challenging^12–14^. Pharmacological treatment is still the most widely accessible and used treatment option for insomnia, with a higher prevalence in the elderly^15–17^. The prolonged use of sedative-hypnotics, defined as a duration of at least three weeks^18^, is particularly common among seniors^19–22^. Benzodiazepines (BZDs, e.g., Diazepam, Clonazepam, Nitrazepam, Oxazepam, Lorazepam, Temazepam) or benzodiazepine receptor agonists (Z-drugs or BZRAs, e.g. Zopiclone) are among the most prescribed class of drugs to manage insomnia complaints. They enhance the inhibitory activity by acting as a gamma-aminobutyric acid (GABA) neurotransmitter GABA_A_ receptor agonists and binding specifically to its α subunits^23^. They are used for their myorelaxant, anxiolytics and sedative-hypnotics properties (i.e., shorten sleep latency, promote sleep continuity)^24^. While BZDs range from short- to long-acting hypnotics (e.g., with a half-life of up to 100 hours), BZRAs are usually shorter-acting (e.g., with a half-life of up to 6 hours), with fewer sedative and myorelaxant side effects^25–27^. Sedative-hypnotic use has been shown to reduce wake duration in placebo-controlled older adults with insomnia disorder when used for one month, concurrently extending their overall sleep duration^28,29^. However, when compared to older adults with and without insomnia disorder, older adults chronically using BZD exhibited a higher arousal density ^30^. Prolonged sedative-hypnotic use in both adults with and without insomnia disorder has been shown to be detrimental to sleep quality by disrupting sleep architecture (including a reduction in time spent in deep sleep (N3) and REM sleep while increasing duration in N2 sleep)^31^. Frequently prescribed^25^, the chronic use of sedative-hypnotics has been also associated with an accelerated decline in both cognitive and physical health^32–35^.

Changes in sleep architecture have been associated with alterations in sleep electroencephalogram (EEG) spectral rhythms. Indeed, compared to older adults with and without insomnia disorder, BZD users exhibited less theta activity and increased beta and sigma power^30,36^. However, BZD users showed an increase in spindle density^37^. During NREM, BZD use has been shown to suppress slow wave activity (SWA), which combines slow oscillation (SO) and delta EEG activity, thus reducing its power^37^. Regarding the intrinsic relationship between those NREM rhythms (i.e., temporal synchrony, phase-amplitude coupling (PAC)), a study on single-night use of Zolpidem in healthy young adults revealed a greater phase angle measure of SO-spindle coupling compared to a placebo condition^38,39^, suggesting that spindles were clustered in the up-state of the SO phase closer to a positive peak. However, the overnight coupling strength between the SO phase and sigma amplitude (i.e., modulation index; MI) exhibited inconsistencies. While a single use of Zolpidem showed comparable effects to a placebo during the NREM period^39,40^, the coupling strength was found to be higher than a placebo in a daytime nap study^38^.

Therefore, further investigation is required to determine whether chronic use of sedative-hypnotics alters interactions between NREM rhythms, particularly in older adults, requires further investigation. Given the diversity in acting duration among sedative-hypnotics, their effects on sleep architecture can vary significantly. Furthermore, sleep fragmentation and duration have been characterized only in a between-group design involving middle-aged (55-65 years old) adults with insomnia complaints using BZDs for prolonged periods compared to healthy sleepers^37^, and older adults with insomnia complaints using BZRAs in placebo-controlled studies^28,29^. These study designs, including three elderly populations (55-80 years old) - healthy sleepers, individuals with insomnia with and without chronic BZD use - mainly focused on describing sleep EEG spectral rhythms and did not assess the effect of BZRAs on EEG spectrum^30,36^. Finally, the description of the intrinsic relationship between SO and spindle focused only on a single use of BZRAs and did not investigate the elderly population^38–40^.

In summary, our comprehension of the chronic impact of BZDs and BZRAs on sleep macro-structure, sleep EEG spectral rhythms, and NREM characteristics such as spindle and SO, as well as their coupling, including the effects of dosage and duration of sedative-hypnotic exposure, and the specific effects of BZDs and BZRAs remains incomplete. Here, we first aimed to further characterize the effect of chronic use of BZD/BZRA on sleep regulation in older adults using a between-group design involving good sleepers, individuals with insomnia disorder who do not take any pharmacological treatment to manage their symptoms and individuals with insomnia disorder who chronically use either BZD or BZRA as sleep-aids. We performed a comprehensive comparison of sleep architecture, EEG relative spectrum, and associated brain oscillatory activities. Specifically, we focused on NREM brain oscillations crucial for sleep-dependent memory consolidation (i.e., SO and spindles) and their temporal coupling. We hypothesized that chronic use of sedative-hypnotics would have a more significant impact on sleep macrostructure compared to drug-free insomnia disorder, resulting in reduced N3 and REM duration, prolonged N2 duration, but greater sleep fragmentation. Regarding EEG spectrum, we anticipated reduced activity in low-frequency bands (i.e., delta and theta power) and greater activity in high-frequency bands such as sigma power indicative of increased spindle activity. We also expected that both spindle and SO characteristics would be impacted, thus altering their coupling. As part of an exploratory analysis, we investigated the differences between BZDs and BZRAs and their effects on the same sleep measures. Additionally, we examined whether sedative-hypnotics dose and duration could mediate these effects. We hypothesized that increased exposure to sedative-hypnotics would lead to greater sleep disruption, with BZD expected to have a more detrimental effect than BZRA.

## MATERIAL & METHODS

### Participants

We investigated 3 groups of older adults: good sleepers (GS), individuals with insomnia disorder who do not use sleeping medication to manage their symptoms (INS) and individuals with insomnia disorder who are chronic sedative-hypnotics users (MED). The INS group were not under any type of sedative-hypnotic treatment at the time of assessment. The MED group was either using BZD (Diazepam, Clonazepam, Nitrazepam, Oxazepam, Lorazepam, Temazepam) or BZRA (Zopiclone) drugs. Data used in this dataset were collected in the scope of different projects conducted in the laboratory and published^41,42^ or registered (ISRCTN13983243, ISRCTN10037794) elsewhere. All groups had similar recruitment process through online and print advertisements posted in the community and from physician referral. Prospective participants underwent screening via a telephone-based checklist to assess inclusion and exclusion criteria, followed by a semi-structured individual interview. Eligible participants underwent a screening polysomnographic (PSG) recording to exclude the presence of additional sleep disorders that could contribute to symptoms of insomnia (i.e. sleep apnea).

Older adults included in the GS group were self-defined good sleepers with no sleep complaints and no sleep dissatisfaction. The INS and MED groups consisted of participants meeting the 3rd version of the International Classification of Sleep Disorder (ICSD) diagnostic criteria for insomnia disorder for at least 3 months^43^. The ICSD-3 criteria for insomnia disorder are defined as self-reported difficulties initiating sleep (i.e., sleep onset latency, SOL > 30min), difficulties maintaining sleep (i.e., WASO > 30min), and/or early morning awakenings (i.e., final awakening time earlier than desired by at least 30 min), for at least 3 times a week and for more than 3 months, combined with complaints of daytime functioning. Participants in the MED group had to meet a criterion of sedative-hypnotic use (regardless of the dosage): BZD or BZRA had to be prescribed for insomnia and to be used for more than 3 nights a week for more than 3 weeks. Averaged consumed doses of BZD and BZRA were converted into Diazepam Equivalent Dose, according to the Equivalence Table of BZD in the Ashton Manuel Supplement^44^.

For all groups, exclusion criteria were as follows: being under 55 y.o or over 80 y.o, major cardiovascular condition or intervention, recent severe infection, medical or unstable condition that could impair physical, psychological, or cognitive abilities (e.g., Parkinson’s or Alzheimer’s disease), poor cognitive function (defined by a Mini Mental State Examination – score ≤24^45^ or a Montreal Cognitive Assessment (MoCA) – < 23^46^), medical conditions likely to affect sleep (e.g., epilepsy, multiple sclerosis, chronic pain, stroke, active cancer), sleep disorders (e.g., moderate or severe sleep apnea (apnea-hypopnea index (AHI) greater than 15 events per hour), bruxism, restless legs syndrome or periodic limb movement (PLM) disorder defined by an index during sleep > 15/h), night shifts or a change in time zone in the last 6 weeks and history of alcoholism or drug abuse. Specifically for individuals in the GS and INS groups, exclusion criteria also included using any sleep-inducing medication (prescribed or over-the-counter) or having received any sleep-related intervention (e.g., cognitive-behavioral therapy for insomnia).

All participants signed a written informed consent form, which was approved by the Concordia University Human Research Ethics Committee and the Comité d’Éthique de la Recherche of the Institut Universitaire de Gériatrie de Montréal. One hundred and one participant were found eligible for the study and distributed as follow: twenty-eight participants in the GS group, twenty-six participants in the INS group as well as forty-seven included in the MED group. Participant demographics are presented in Table 1.

**Table 1.**
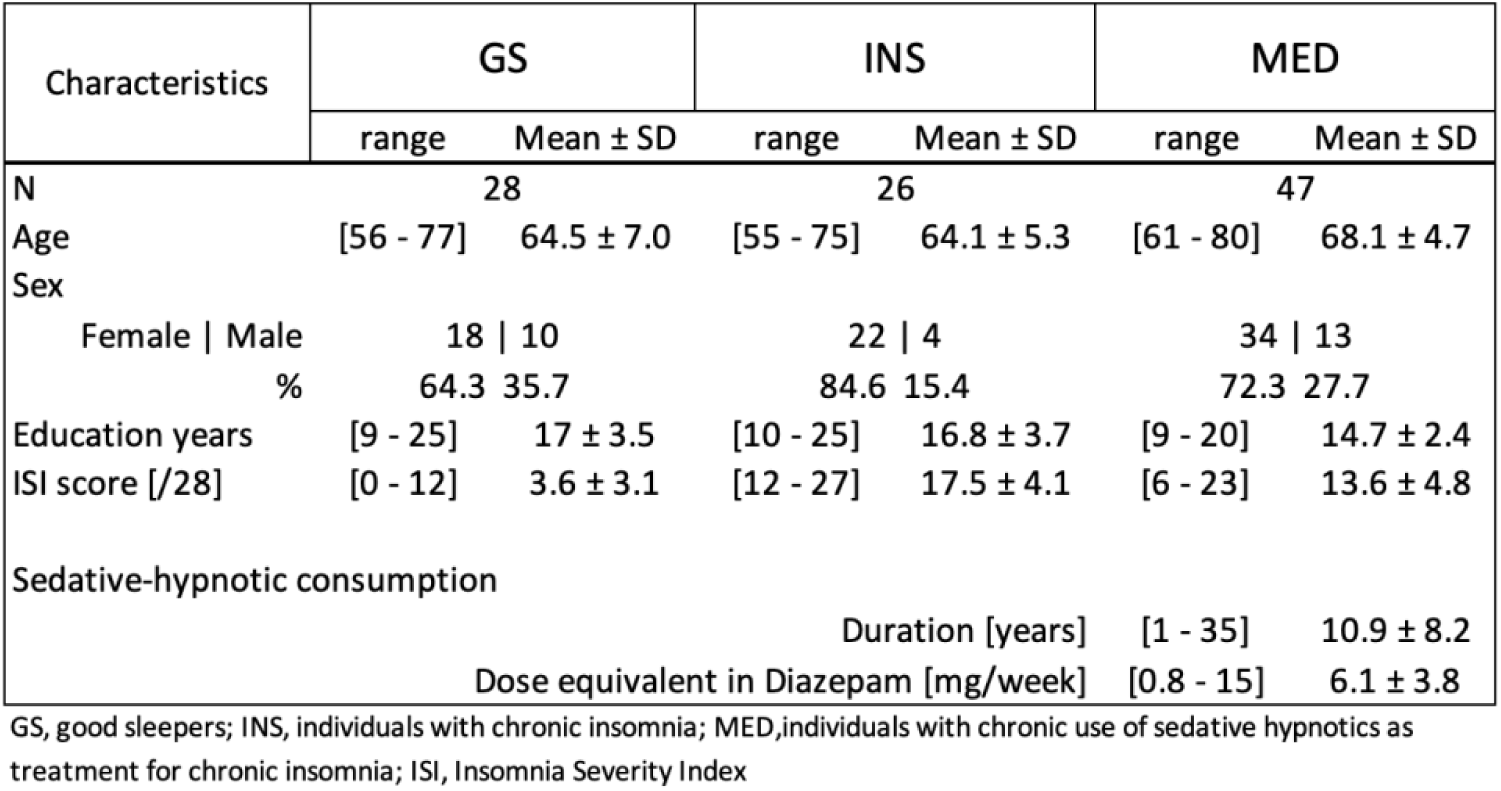
Demographics.

### Study protocol

Once eligible, participants underwent a screening PSG, which also served as a habituation night. Depending on the specific projects they were recruited for, participants then completed at least one experimental night and subsequently filled out the Insomnia Severity Index (ISI) questionnaire the following morning. For the present study, sleep characteristics were extracted from the first experimental night.

### Insomnia Severity Index (ISI)

Participants completed the ISI questionnaire, a 7 Likert-scale used to assess the nature, severity, and impact of current insomnia symptoms^47^. The total ISI score ranges from 0 to 28, with higher scores indicating more severe insomnia. Its overall Cronbach’s α was 0.90.

### Polysomnographic (PSG) recording

Whole-night PSG recordings were used for each night, including EEG, electromyogram (chin and legs EMG), electrooculogram (EOG), and electrocardiogram (ECG). During the screening night, the PSG setup included an oximeter, thoracic and abdominal belts, oral-nasal thermistor, and nasal cannula. From the habituation night, we computed the apnea-hypopnea index and periodic leg movement index to exclude potential sleep disorders. For the experimental night, the EEG montage included 13 electrodes (Fz, F3, F4, Cz, C3, C4, Pz, P3, P4, O1, O2, M1, M2) positioned on the scalp according to the 10-20 system AASM guidelines. EEG signal was recorded by a Somnomedics amplifier (SomnoMedics, Germany) at a sampling rate of 512 Hz, referenced to Pz online for monitoring and to contralateral mastoids (M1 and M2) offline for analysis.

### EEG analysis

All sleep scoring and analyses were conducted using the Wonambi python toolbox (https://wonambi-python.github.io) and the Seapipe python toolbox (https://github.com/nathanecross/seapipe). Sleep stages (N1, N2, N3, REM) and wake 30-s epochs scoring as well as the detection of arousal and artefact events were visually scored according to the AASM rules^43^. From the scoring, we computed the calculation of the following: total sleep period (TSP; from sleep onset to final awakening), total sleep time (TST; sum of the time spent in different sleep stages), sleep onset latency (SOL; from light off to the first epoch of sleep), wake after sleep onset (WASO; sum of the nocturnal wake epochs), time spent in bed (TIB; from light off to light on), sleep efficiency (SE; TST/TIB*100), arousal density (number per 30 s), stage switch index (SSI; number of transitions between sleep stages per hour), sleep fragmentation index (SFI; number of transitions from deep to lighter sleep stages per hour) as well as the % of time spent in each stage (%/TSP).

The EEG spectrum power density average (30 sec of time resolution with artefact excluded) was computed using a Fast Fourier Transformation and Welch’s method (Overlapping: 50%; Resolution: 0.25 Hz). Mean relative power for the following frequency ranges was calculated as absolute power divided by broad spectrum (0.5-35Hz) total power: SO (0.25-1.25 Hz), delta (0.5-4 Hz), theta (4-7.75 Hz), alpha (8-11 Hz), sigma (11.25-16 Hz), beta (low: 16.25-19 Hz and high: 19.25-35 Hz).

For spindle detection, the highest center frequency peak (integral of the Gaussian fit) over the sigma range specific for each subject was obtained^48,49^. Using those participant-specific adapted sigma ranges, we determined with a 2Hz bandwidth the highest peak in the 10 to 13 Hz range for midline frontal (Fz – slow spindle) and the 13 to 16 Hz range for midline parietal (Pz - fast spindle) on artefact-free derivations accounting for spindle frequency gradient^50,51^. Spindles were automatically detected using a validated algorithms^52–54^ and implemented in the Wonambi and Seapipe toolboxes. The spindle detection algorithm consisted of computing the root-mean-square (RMS) of the participant-adapted sigma band with a 0.5 sec overlap window and smoothed with Gaussian filter^55,56^. RMS were identified as a spindle event when values exceeded a threshold at 2 SD above the mean peak amplitude. Detection criteria included a spindle duration ranging from 0.5 to 3 sec.

SO events were detected on a fixed band-pass FIR filter from 0.16 to 1.25 Hz on artefact-free EEG recordings of Fz using a published algorithm^57,58^ and implemented in the Wonambi and Seapipe toolboxes. SO candidates were flagged using a duration criteria of two positive-to-negative zero crossings within 0.8 to 2 sec. Then, trough-to-peak amplitudes between two positive-to-negative zero crossings were computed on SO candidates and only the top 25% highest amplitudes were considered as SO in the study.

For SOs and spindles detected in NREM (N2 + N3), we extracted the following characteristics: density (i.e., mean number of spindles/SO per epoch of 30 s), amplitude (µV), duration (sec) and peak frequency (Hz).

Spindle and SO coupling was assessed using three complementary measures: event co-occurrence, preferred coupling phase, and the modulation index as we previously published^42,59^.

SO-spindle temporal co-occurrence was determined via the intersection-union rule. SO+ is defined as the proportion of the SO that co-occurs with spindles. Spindle+ is defined as the count of spindles linked with SO divided by the total count of spindles. The intersection/union threshold was set at 10% overlap duration between SO and spindle.

We investigated SO (0.5 to 1.25 Hz) and participant-specific adapted sigma band (Fz: 10-13Hz; Pz: 13-15Hz) phase-amplitude coupling based on each SO event detected across the whole night. Each SO event was filtered with a 2-second buffer on each side to catch filter edge effects. Using a Hilbert transformation and FIR filter, we extracted instantaneous phase time series within SO frequency bands and amplitude time series within sigma frequency bands. After removing the buffer, we paired the instantaneous amplitude time series with the corresponding phase values and categorized them into 18 bins^60^. We then computed the average amplitude for each phase bin, resulting in a single distribution that represents the average amplitude across each SO phase. These average amplitudes in each phase bin were z-scored independently for each SO event^61^. The phase bin with the highest average amplitude of sigma activity was identified for each SO event. The preferred coupling phase (CP) was determined using circular statistics to find the mean direction across all NREM SO events.

The MI quantifies the relationship between the SO phase and the amplitude of participant-specific adapted sigma bands, revealing the strength of phase-amplitude coupling^60^. MI values range from 0 to 1: a value of 0 indicates a uniform distribution of amplitudes across all phase bins, suggesting no preferential coupling between phase and amplitude. In contrast, a value of 1 indicates that the signal’s amplitude is consistently highest at a specific phase angle (CP), indicating stronger and more consistent phase-amplitude coupling. The MI was calculated as (Hmax - H) / Hmax, where H is the Shannon entropy of the phase-amplitude probability distribution, and Hmax the maximum distribution entropy^60^. We employed a permutation-based method to evaluate the significance of the observed coupling strength compared to a randomized phase-amplitude coupling distribution^62^. All SO events detected across the whole night were concatenated into equal segments. For each segment, we performed permutations, where the SO phase time series were shuffled with respect to their corresponding participant-specific adapted sigma band amplitude. We recalculated the raw MI to establish a random distribution of raw MI. Subsequently, we computed z-scores for the observed MI with respect to the null distribution. Finally, these z-scores were averaged across all segments to derive the normalized MI.

Supplementary analyses at the midline central electrode (Cz) were conducted to compute the average EEG spectrum power, detect SO and spindles, and investigate their temporal co-occurrence and coupling. These findings can be found in Supplemental Results.

Further analyses per sleep stage N2 and N3 separately, along with the use of other methods for spindle detection^53,54^ and SO detection^57^ are also presented in Supplemental Results.

### Statistical analyses

Statistical analyses were performed using RStudio 1.2.50 (RStudio, Inc., Boston, MA) and R package (ggplot2, sjstats, sjmisc, forcats, emmeans, rstatix). Normality of distribution was checked with Shapiro tests and homogeneity of variance was tested with Levene tests.

Our main objective was to comprehensively characterize the effect of chronic sedative-hypnotics use on sleep compared to drug-free insomnia and good sleepers. We first investigated differences in macro-architecture (including wake duration, SOL, duration in sleep stages, sleep efficiency and markers of sleep fragmentation). Additionally, we analyzed differences in EEG relative spectrum power, particularly delta and sigma power amplitude, as well as SO and spindle events density and amplitude, and phase-amplitude coupling measures, such as the modulation index. Analyses of variance (ANOVA) with Group as a between-subject factor (GS vs INS vs MED) and unpaired-posthoc test were computed to investigate the difference between groups on our sleep-derived measures. We performed non-parametric tests (Kruskal-Wallis test and Dunn test) only when the variance or normality of distribution was not homogeneous. All analyses were conducted while adjusting for age. Between-group effect sizes were calculated using Hedge’s g (which corrects for small sample sizes). Watson circular statistic test for non-uniformity of circular data was performed to test whether spindles had a to the SO phase^63^.

As an exploratory analysis, we investigated the distinctive effect between sedative-hypnotic classes (BZD vs BZRA) on the same sleep characteristics, and whether sedative-hypnotic exposure could mediate these effects. Analyses of variance (ANOVA) with sedative-hypnotic class as a between-subject factor (BZD vs BZRA) was used to investigate the difference between groups on our sleep-derived measures. We performed non-parametric tests (Wilcoxon test) only when the variance or normality of distribution was not homogeneous. Analyses were conducted while adjusting for sedative-hypnotics equivalent dose. Additionally, non-parametric Spearman correlations were computed to test the relationship between our sleep-derived measures, and both treatment duration and medication equivalent dose in Diazepam on the 47 elderly participants with chronic use of BZD or BZRA.

The level of significance was set to a p-value of <.05 and p-values were adjusted for multiple comparisons (Benjamini-Hochberg/FDR correction). For significant results, both raw (*p*) and adjusted p-values (*q*) were reported when necessary.

## RESULTS

One hundred and one individuals (66.05 ± 5.84 years, 73% female) were categorized into three groups: good sleepers (GS, n=28), individuals with insomnia (INS, n=26) or individuals with insomnia who chronically use sedative-hypnotics (either BZD or BZRA) to manage their insomnia difficulties (MED, n=47; dose equivalent in Diazepam: 6.1 ± 3.8 mg/week); see Table 1 for demographics and Table S1 for the various sedative-hypnotics used. We found that participants in the MED group were older compared to both GS and INS groups (all *p<.*02), while no difference was found regarding sex ratio (Fisher, *p=.25*). Models reporting the effects of age on our sleep-related measures were detailed in Table S2-S4. The GS group displayed no clinically significant insomnia (ISI score: 3.6 ± 3.1), while both the INS and the MED groups exhibited higher insomnia severity (ISI score > 8) (Kruskal-Wallis, *p<.*001; see Figure S1). The MED group reported a lower ISI score compared to the INS group (INS group: 17.5 ± 4.1; MED group: 13.6 ± 4.8; *p<.*002).

### Chronic use of BZD and BZRA affects sleep architecture

There was a Group effect for SE (*F*(2,97)=16.5, *p<.001, q*=.001), where both INS and MED displayed lower SE compared to the GS group (all *p*<.001, all *q*<.002) (Figure 1A). We also found a Group*Stage interaction (*F*(8,9215)=220.75; *p*<.001) on the proportion of time spent in each sleep stage (over TSP) with significant main effects of Stage (*F*(4,95)=10034; *p*<.001) and Group (*F*(2,97)=53; *p*<.001). Specifically, we found that the MED group spent more time in N1 and N2 and less time in N3 compared to both INS and GS groups (all *p*<.001). The MED group also increased their time spent awake compared to the GS (*p*<.001) but not the INS group (Figure 1B). Meanwhile, no significant difference was observed between the three groups for REM duration (Figure 1B). In line with the differences in NREM stages, we found a main effect of Group on latency to N3 (*F*(2,97)=13, *p*=.002 *p*=.001, *q*=.002) and REM (*F*(2,97)=12.2, *p*=.002, *q*=.003) as MED displayed longer latencies to both stages compared to the INS and GS groups (all *p*=.004, *q*=.01). However, we did not find any significant effect for SOL (Figure 1C) or latency to N2. The INS group also exhibited more arousal than the GS and MED groups and lower SFI than the MED group (see Table 2 and Supplementary Results).

**Figure 1:**
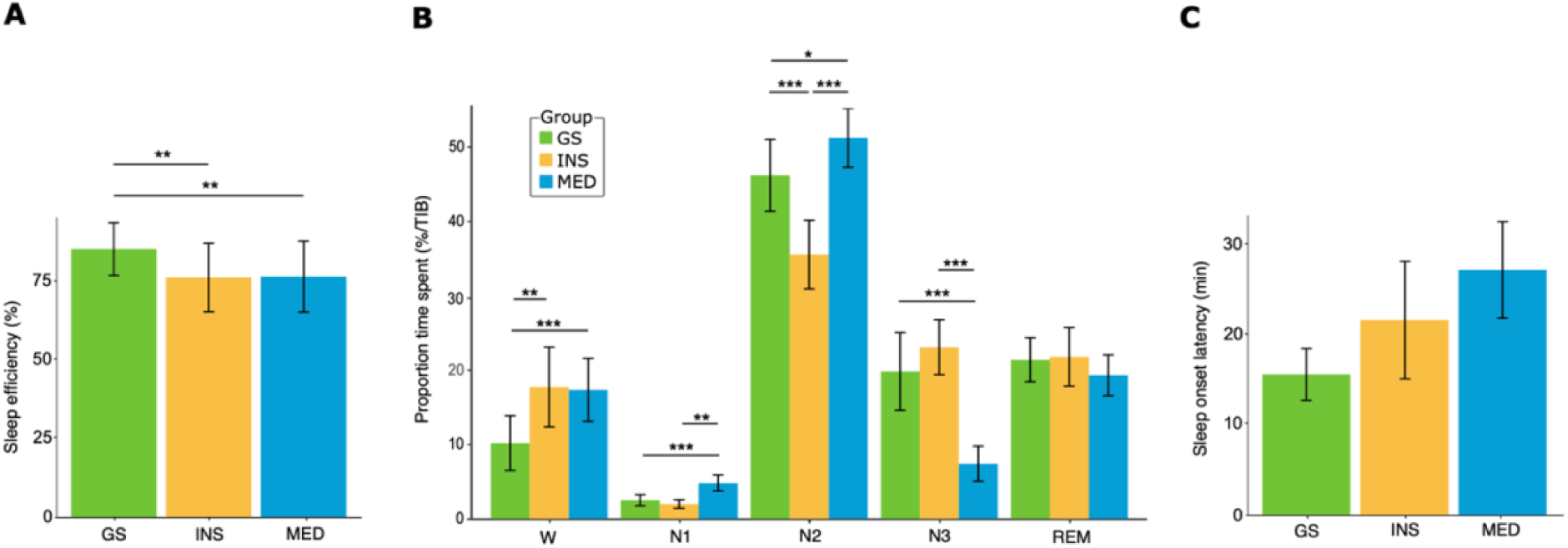
Chronic sedative-hypnotic use affects sleep architecture. For the GS (green), the INS (orange), and the MED (blue) group: (A) *Mean sleep efficiency (%) (± SD)* (B) *Mean proportion time spent in wake and sleep stages (% over TIB) (± SD)* (C) *Mean SOL duration (min) (± SD) Asterisks represent significance (p): *<0.05; **<0.01; ***<0.001*

**Table 2.**
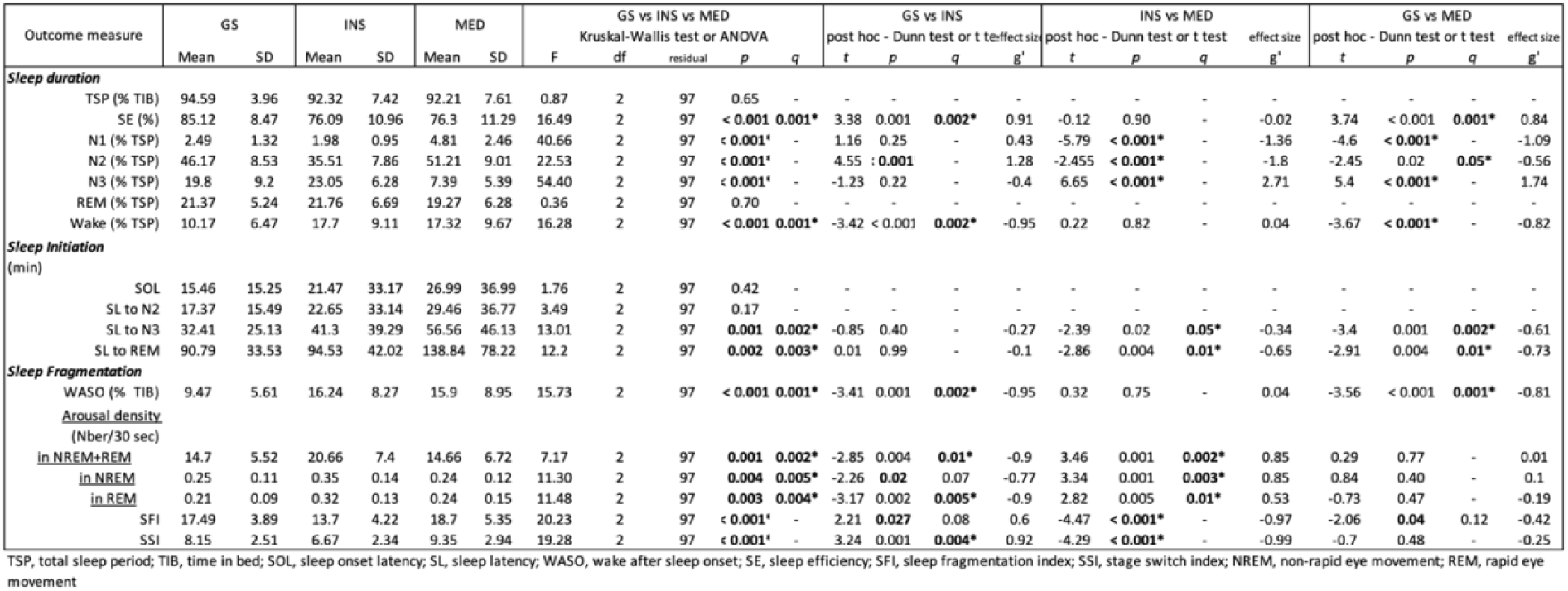
Effects of chronic sedative-hypnotic use on sleep architecture.

In summary, participants with insomnia (MED and INS) showed changes in sleep architecture, resulting in lower SE with or without chronic sedative-hypnotic use. Additionally, chronic sedatives-hypnotic use worsened sleep quality by prolonging light sleep (N1 and N2) duration and reducing deep sleep (N3), with increased latencies to sleep stages N3 and REM.

### Chronic use of BZD and BZRA alters spectral activity

Power analysis of sleep EEG revealed Group effects in specific spectral bands (see Table S5-S6 and Supplementary Results). Compared to both the INS and GS groups, MED exhibited a lower relative power in the theta band over the frontal (Fz; *F*(2,97)=11.8, *p*=.003 *q*=.005) and posterior (Pz; *F*(2,97)=15.6, *p*<.001, *q*=.002) electrodes in both NREM sleep (N2 and N3 combined) and REM sleep (Figure 2). The MED group also displayed a higher relative power in the frontal sigma (Fz; *F*(2,97)=15.3, *p*<.001, *q*=.001) and lower beta bands (Fz; *F*(2,97)=18.4, *p*<.002, *q*=.001) during NREM (Figure 2A), as well as greater relative power in frontal and parietal high beta bands in NREM sleep (Fz-*F*(2,97)=16.9, *p*<.003, *q*=.001 - Figure 2A; Pz - *F*(2,97)=8.7, *p*=.013, *q*=.044 - Figure 2B).

**Figure 2:**
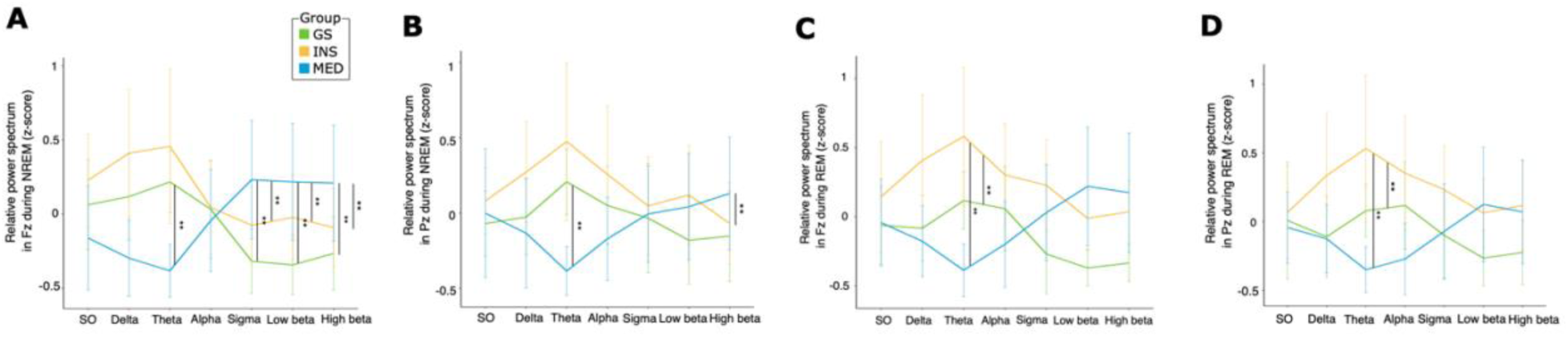
Chronic sedative-hypnotic use alters spectral activity For the GS group (green), the INS group (orange), and the MED (blue) group: (A) *Mean relative spectral activity in Fz during NREM (z-score)* (B) *Mean relative spectral activity in Pz during NREM (z-score)* (C) *Mean relative spectral activity in Fz during REM (z-score)* (D) *Mean relative spectral activity in Pz during REM (z-score) Asterisks represent significance (p): *<0.05; **<0.01; ***<0.001*

We did not find any significant Group effects for relative power in the SO, delta and alpha frequencies. During REM, the MED group displayed less theta relative power than the two other groups (all *p*≤.001, all *q*≤.002) in both frontal (Figure 2C) and parietal (Figure 2D) regions.

### Both spindles and SOs are altered with chronic use of BZD and BZRA

During NREM sleep, a Group effect was observed on frontal spindle density (*F*(2,97)=8.4, *p*=.01, *q*=.04) with the MED group exhibiting greater spindle density compared to the INS group only (*p*=.004, *q*=.01). However, there was no significant Group effect on spindle density detection on the posterior channel (Pz – *F*(2,97)=1.3, *p*=.28). Spindle amplitude was found lower in the MED group compared to both INS and GS groups (*F*(2,97)=7.8, *p*=.02, *q*=.04) (all *p*<.02, all *q*<.07) in the frontal region. In the parietal region, spindle amplitude was lower in the MED group compared to the INS group only (*F*(2,97)=7.7, *p*=.02, *q*=.08), but this difference did not survive multiple comparisons correction. We found no significant effect of Group on other spindle characteristics (Figure 3A and Table S7-S8). We observed similar findings using other published methods (Ray, Lacourse; see Supplemental Material Table S9).

**Figure 3:**
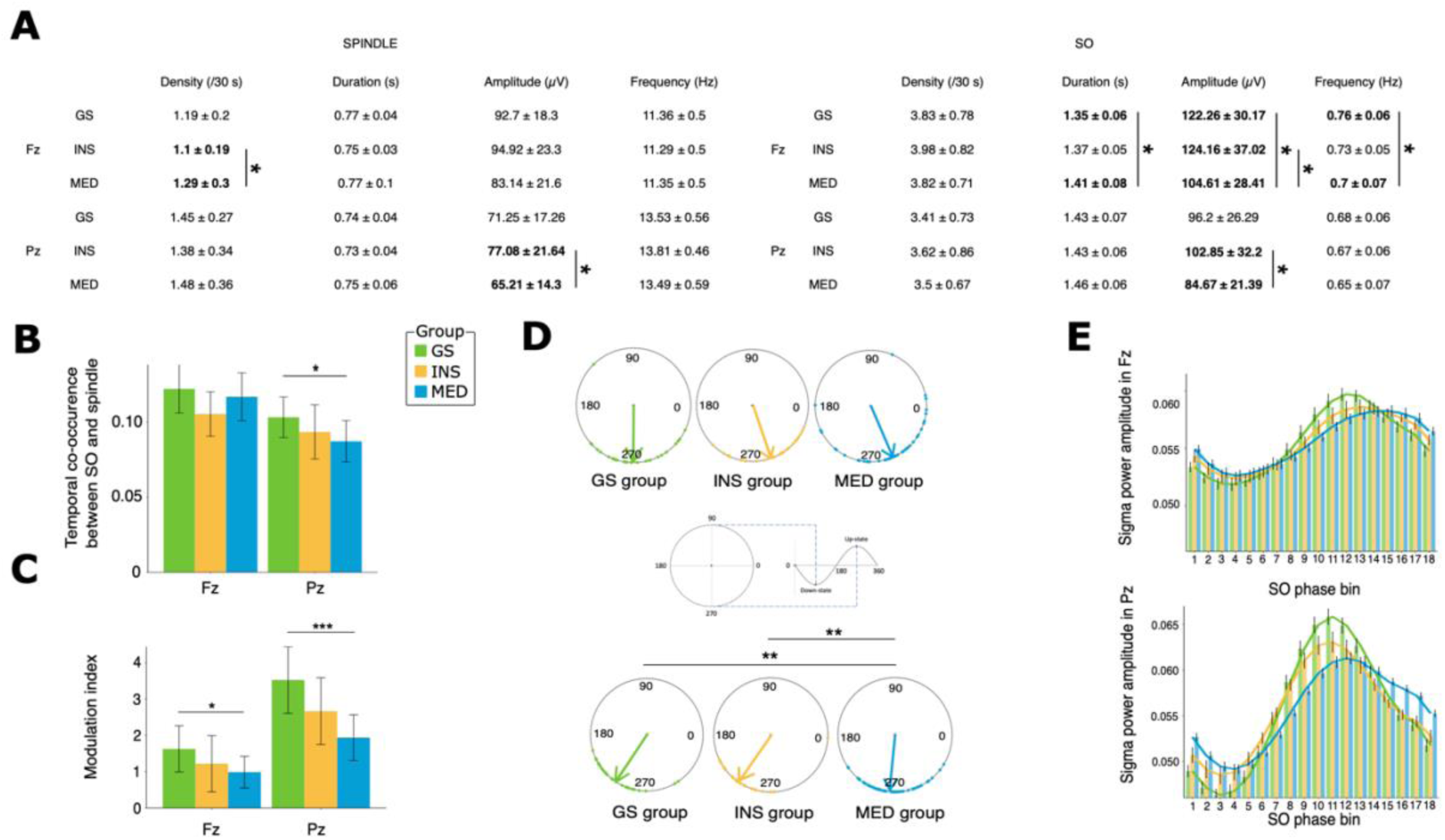
Chronic sedative-hypnotic use impact on SO-spindle and their association. For the GS group (green), the INS group (orange), and the MED (blue) group: (A) *Mean spindle (up) and slow oscillation (down) characteristics (± SD)* (B) *Mean temporal association between SO and spindle (± SD)* (C) *Mean Modulation index (± SD)* (D) *Preferred-phase coupling polar plot for Fz (up) and Pz (down) during NREM. Arrows represents the average coupling preferred-phase for each group*. (E) *Phase-amplitude histograms for Fz (up) and Pz (down)*. *Each bar represent the average sigma power mean amplitude (µV) (± SD) across 18 bins, where each bin represent 20°*. *Asterisks represent significance (p): *<0.05; **<0.01; ***<0.001*

We found a Group effect on frontal SO characteristics, including SO duration (*F*(2,97)=9.6, *p*=.01 – Figure 3A, right), amplitude (*F*(2,97)=9.5, *p*=.01) and frequency (*F*(2,97)=16.2, *p*<.001, *q*=.001). Specifically, SO duration was longer in the MED group compared to the GS group (*p*=.003, *q*=.01) but not to the INS (*p*=.03, *q*=.1) groups. The MED group displayed lower SO amplitude compared to both GS and INS (all *p*=.01, all *q*≤.04) groups. The MED group also exhibited a slower SO peak frequency compared to both GS (*p*<.001) and INS (*p*=.03, *q*=.09) groups. Similar trends were found in SOs detected on parietal region, although these did not pass corrections for multiple comparisons (Table S7-S8). We did not find any significant Group effect on SO density in both frontal and parietal region.

Using a more rigid SO detection algorithm based on fixed amplitude criteria (Massimini2004), we observed that the MED group had longer SO durations in both the frontal and parietal regions compared to the GS and INS groups (all p<.02) (Table S10). Additionally, we noticed a lower SO density in the frontal area for the MED group compared to both GS and INS groups (all p<.05).

### Chronic BZD and BZRA use affected SO-spindle association

The only significant effect on the temporal co-occurrence of SO and spindles was a Group effect in Pz (*F*(2,97)=7, *p*=.03, *q*=.06 – Figure 3B) - driven by the MED group displaying lower co-occurrence compared to the GS group (*p*=.008, *q*=.02), although it did not pass corrections for multiple comparisons (Table S11). We did not find any significant Group effect on the temporal co-occurrence of spindles and SOs in frontal regions (Table S11).

We found a Group effect in frontal on the PAC between SO and sigma (MI; *F*(2,97)=6.2, *p*=.04 – Figure 3C). Post-hoc tests revealed that the MED group displayed lower coupling strength (MI) compared to the GS group (*p*=.014, *q*=.04), but no difference was found between the MED and INS groups, as well as between the GS and the INS group. The same Group effect was observed in the parietal channel Pz (*F*(2,97)=17.9, *p*<.001 – Figure 3C) driven again by differences between the MED and GS groups (*p*<.001).

Furthermore, we found a Group effect on the coupling phase in the parietal regions (Figure 3D) driven by the MED group which presented a greater (delayed) preferred coupling phase (CP) compared to both the GS (*W*= .4, *p*<.01) and INS (*W*= .3, *p*<.01) groups. Phase-amplitude histograms (Figure 3E) represent this delay in SO-sigma in the MED group.

Furthermore, we investigated which of the previously described changes in SO and spindle characteristics were associated with alterations in their coupling strength (MI). We found that the coupling strength (MI) was positively associated with spindle amplitude in both frontal (*p*=.03, *q*=.08, *r*= .31) and Pz (all *q*<.003, all *r>* .49) (Figure S2 and Table S12). SO amplitude was also found associated with MI in both frontal and parietal regions (all *q*≤.003, *r*≥ .49), although this effect was not specific to the MED group, as it was shared among the three groups.

In summary, the MED group displayed a lower temporal co-occurrence of SOs and spindles, including a weaker coupling strength (MI) and a later preferred coupling phase when compared to good sleepers.

### Effect of the use and type of chronic sedative-hypnotic exposure on sleep

A subgroup analysis was conducted within the MED group, including first both BZDs and BZRAs users (n=47) (Table S13-S16). Significant correlations were observed between higher sedative-hypnotic dosage and increased SOL (*p*=.003, *q*=.01, *r=*.45), as well as latency to reach stage N2 (*p*=.002, *q*=.01, *r=*.44), and N3 (*p*<.001, q<.001, *r=*.51 – Figure 4A), and while it did not survive corrections for multiple comparisons, decreased TST (*p*=.02, *q*=.07, *r=*-.33) and greater latency to REM (*p*=.04, *q*=.08, *r=*.31) (Table S13). Additionally, we observed a correlation between higher sedative-hypnotic dosage and increased relative power in high-frequency during NREM, including sigma (*p*=.004, *q*=.01, *r=*.41 – Figure 4B) and beta (*p*=.003, *q*=.01, *r=*.42 – Figure 4C) bands (Table S14). We observed a correlation between a longer duration of sedative-hypnotic consumption and decreased posterior spindle duration (Pz; *p*=.004, *q*=.02, *r*= -.41 – Figure 4D) but no other sleep variables were associated with duration of use (Table S15).

**Figure 4:**
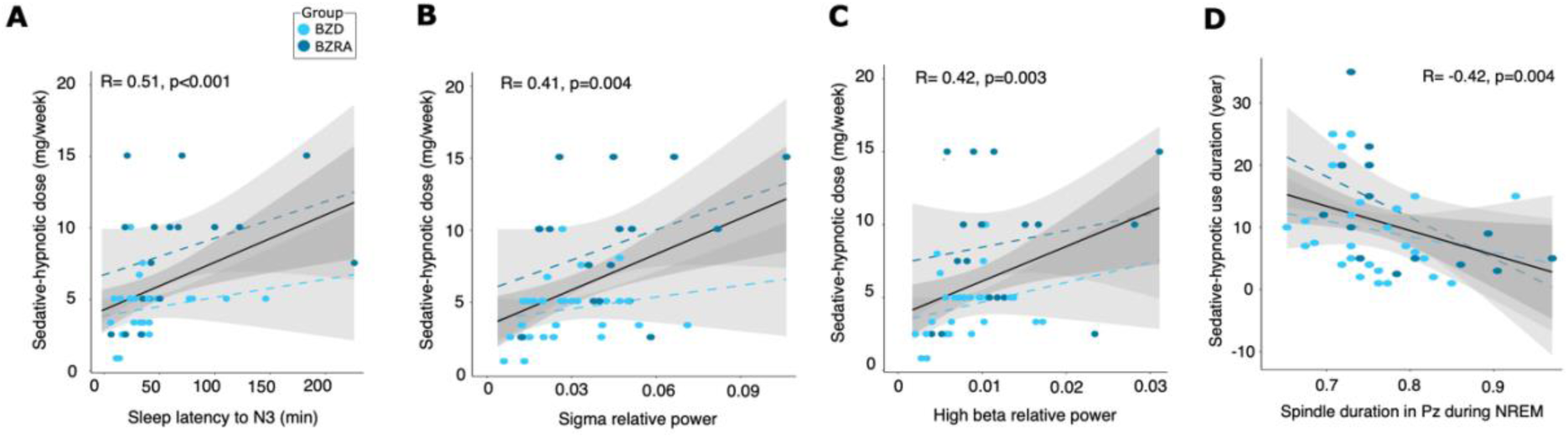
Impact of sedative-hypnotic dose and duration. For the BZD (darkblue) and the BZRA group (lightblue): (A) *Scatterplot showing correlation between the change in the dosage and the change in the sleep latency to N3* (B) *Scatterplot showing correlation between the change in the dosage and the change in the sigma relative power* (C) *Scatterplot showing correlation between the change in the dosage and the change in the high beta relative power* (D) *Scatterplot showing correlation between the change in the sedative-hypnotic use duration and the change in the spindle duration in Pz*

We then categorized participants in the MED group based on their type of sedative-hypnotic class, into either the BZD (n=18; dose equivalent in Diazepam: 8.75 ± 4.39 mg/week) or the BZRA (n=29; dose equivalent in Diazepam: 4.41 ± 2.01 mg/week) group, while adjusting for sedative-hypnotic equivalent dose. We did not find any significant group effect on age, sex or duration of hypnotic consumption between the two groups (all *p*>.05), but we found a group effect driven by BZD which displayed greater sedative-hypnotics dose equivalent in diazepam (*p*=.002) compared to the BZRA group. We observed that the BZD group displayed lower TST (*F*(2,44)=6.1; *p*=0.02, *q*=0.2) and SE (*F*(2,44)=4.4; *p*=0.04, *q*=0.24), as well as SL to N3 (*W*(2,44)=143; *p*=0.03, *q*=0.2) compared to the BZRA group, but that did not pass multiple comparison corrections (Table S13). We found a group effect on high-frequency bands in the frontal area driven by the BZD group, which displayed greater beta (all *p*<.02, all *q*=.07) relative power compared to the BZRA group, although these did not pass corrections for multiple comparisons (Table S14). No significant differences were found between BZD and BZRA regarding spindle characteristics, and SO-spindle coupling (Tables S15-S16).

Additional analyses were conducted to balance age between groups and evaluate the impact of BZRA on sleep quality, by comparing the same GS and INS groups with a third group consisting solely of BZRA users (BZRA group). Chronic BZRA use extended the duration of lighter sleep stages (N1, N2), reduced deep sleep, and increased arousal density compared to the INS group (Tables S17). The BZRA group also showed greater high-frequency bands (sigma, beta), although it did not pass corrections for multiple comparisons (Tables S18). Spindle density was greater in the frontal area and lower in the parietal area, but these findings also did not survive corrections for multiple comparisons (Tables S19).

In summary, the alteration in sleep architecture and spectral activity became more pronounced as the dosage of sedative-hypnotics increased. However, the characteristics of SOs and spindles, including their temporal co-occurrence and coupling strength, showed no association with medication dosage. The duration of sedative-hypnotic use was only associated with a decrease in posterior spindle duration (Pz). Additionally, chronic use of BZD displayed more pronounced changes in sleep architecture and spectral activity compared to BZRA.

## DISCUSSION

This study investigated the effect of chronic sedative-hypnotic use on sleep architecture in older adults, including good sleepers, individuals with insomnia who do not take any pharmacological treatment for sleep and individuals with insomnia who chronically use BZD/BZRA for insomnia management. We found that chronic use of use BZD/BZRA influenced sleep regulation without enhancing sleep duration or reducing its fragmentation. Alterations in relative spectral power were evident, particularly in theta range and in sigma and beta activity. Chronic sedative-hypnotic use also altered spindle and SO characteristics and related phase-amplitude coupling. Finally, we found that higher doses of BZD/BZRA use correlated with worsened sleep parameters.

Drug-free older adults with insomnia (INS) displayed lower efficiency, together with greater wake duration and sleep fragmentation when compared to good sleepers (GS), which is consistent with typical insomnia complaints. Surprisingly, the use of chronic sedative-hypnotics (MED) did not improve sleep patterns as overall wake duration, as well as sleep fragmentation were not significantly different when comparing INS and MED groups. Furthermore, the chronic use of BZD/BZRA degraded sleep architecture more than insomnia alone: increasing N1 and N2 duration, reducing N3 duration, and extending latency in both N3 and REM stages. Those current findings are comparable to previous studies with similar study designs and clinical groups^30,36^ where the chronic use of BZD (BZRA was not studied) greatly altered sleep architecture compared to drug-free insomnia and good sleepers^30,36,37^. It has been shown that sedative-hypnotics increase sigma EEG activity by potentializing the intra-thalamic inhibition, with a parallel increase in N2 duration^64–66^. In contrast, one study found that acute use of BZRA increased N3 duration, however it was conducted in young good sleepers only^67^. Reduction in time spent in N3 has been linked to impaired cognitive performance^68^, reduced cortical volume^69^, as well as a reduction in cerebral metabolism and altered brain clearance^70^. Thus, BZD/BZRA related-sleep architecture alterations may increase the vulnerability of older adults.

Alterations in sleep architecture due to chronic use of sedative hypnotics were also captured in the differences in EEG spectral power and brain oscillation characteristics. Consistent with the literature on BZD/BZRA use in middle-aged to older adults with and without insomnia, we found a significant reduction in the relative theta power during both NREM and REM sleep compared to GS and INS^30,36,37,71,72^. Theta activity generation involves GABAergic neurons^73–75^. Consequently, the inhibitory effects induced by sedative-hypnotics may result in a lack of theta activity.

Moreover, we found a reduction in the ratio between slower to higher EEG frequencies in chronic users of sedative-hypnotics. A previous study found a reduction in absolute delta activity associated with chronic BZD (not BZRA) use in the elderly, which was not observed in our groups^30^. Despite the absence of change in delta power, we found that SO characteristics were altered by the chronic use of sedative-hypnotics compared to both INS and GS.

Specifically, SO amplitude was reduced in both frontal and parietal regions while SO duration was longer, and frequency was slower in the frontal region. The current findings align with a study demonstrating that acute BZRA use in both schizophrenia patients and healthy controls resulted in increased SO duration and decreased SO amplitude across the whole scalp^76^. Importantly, we observed that the choice of SO detection had an impact on the quantity and characteristics of detected SO events in this sample of older adults. The detection method using an adapted amplitude threshold (top 25% of SOs with the highest amplitude^58^) identified a higher number of SO events in older adults compared to the detection with a rigid threshold (SOs with peak-to-peak amplitude > 140 µV^57^). The adapted detection method^58^ was especially more effective in identifying interindividual variations in SOs among older adults, which may have functional implications for daytime functioning, such as memory consolidation. It is well established that slow wave activity (SWA; present during N3 and comprising SO and delta waves) plays a crucial role in synaptic and cellular sleep-dependent memory consolidation^77^. Thus, these findings of impaired slow oscillations in chronic users of sedative-hypnotics suggest that these disruptions might interfere with sleep-dependent memory consolidation.

Furthermore, we found that chronic sedative-hypnotics altered other NREM brain oscillations such as spindles and related sigma power. Specifically, slow spindle density increased while the amplitudes of both slow and fast spindles were reduced. Previous studies have observed an increase in sigma activity as well as slow spindle density, in healthy young adults following acute BZRA administration^76,78^, and schizophrenia outpatients^79^. However, an increase in spindle amplitude has also been reported^78^. Therefore, the pharmacological effects of sedative hypnotics on spindle density appear not influenced by the duration of sedative-hypnotic use, while spindle amplitude may be associated with tolerance effects. This suggests also that the pharmacological impact on spindle characteristics might interfere with synchronous neuronal activity among thalamocortical neurons. Given that communication between thalamocortical neurons and thalamic reticular neurons is crucial for spindle and SO synchronization^80^, such chronic use of sedative-hypnotics may also disrupt the synchronization of NREM oscillations also called coupling.

While spindle and SO characteristics are essential for sleep-dependent memory consolidation, their coupling appears critical. Spindle temporally linked with SO showed a greater calcium activity in pyramidal neurons, compared to isolated spindle and SO which may promote long-term synaptic dendritic plasticity^81^. In the present study, while we found that chronic use of sedative-hypnotics increased spindle density, it also weakened the temporal link between SO and spindles. Specifically, it delayed the SO-sigma phase-amplitude coupling and the preferred phase of this coupling in both the frontal and parietal regions. The present findings align with the existing literature showing that the consistency of SO phase locking with spindles is reduced following acute use of BZRA in schizophrenia outpatients compared to healthy controls^76^. A study found that BZRA increased the phase-amplitude coupling between SO and spindle as verbal memory consolidation improvement^38^. However, the population involved in the study consisted of healthy young good sleepers with acute use of sedative-hypnotics^38^, whereas the present study focused on older adults with chronic use. As synchronization between SOs and spindles mediate memory consolidation^58,62,77,82,83^, this desynchronization between NREM oscillations induced by chronic use of sedative-hypnotic may have significant detrimental implications for plasticity and memory processes, which is especially concerning in an elderly population.

Our results suggest that chronic use of sedative-hypnotics seems detrimental to sleep compared to drug-free GS and INS. Such alteration of sleep regulation – at the macro and micro-level - may explain the reported strong link between sedative-hypnotic use and cognitive impairment in older adults^33^. Indeed, chronic use has been demonstrated to accelerate cognitive decline and morbidity^84–86^. Future studies should assess whether altered SO and spindle characteristics and their coupling due to chronic use of sedative-hypnotics might relate to memory performance. More importantly, other approaches whether pharmacological^87–89^ or non-pharmacological^90–92^ need to be considered to address insomnia complaints while preserving the integrity of the intrinsic mechanisms related to cognitive functioning.

To our knowledge, no previous study has directly and systematically compared sleep macro- and micro-architecture between BZD and BZRA users. It would be expected that BZRA might be less detrimental because of their usually shorter-acting pharmacokinetic effect. Indeed, our findings indicate that chronic use of BZD decreases sleep efficiency and increases latency to deep sleep, as well as elevating power in high-frequency EEG more than BZRA, with a dose-dependent effect, regardless of exposure duration. Since this effect was not attributed to the higher dosage of BZDs compared to BZRAs, the specifically pronounced effect of BZDs may be explained by their longer-acting activity in the nervous system. Additionally, BZRA use did not significantly affect sleep latency or spindle and SO characteristics, although their coupling was disrupted, and this may be due to the shorter-acting activity of BZRAs. This suggests that shorter-acting medications might have a lesser negative impact on NREM brain oscillations in older adults compared to long-acting medications, with the effect being more influenced by pharmacokinetics than by the duration of use. This may have impacts on the development of neurodegeneration. Indeed, studies have found associations between sedative-hypnotic use and increased risk of developing dementia, increasing specifically with cumulative dose and exposure and when long-acting medication was used^34,93–96^. However, conflicting evidence exists at higher doses^35,97,98^. Future studies should further clarify the discrepancies in these findings and investigate the differing effects of these medications on sleep.

Some limitations of this study may affect the interpretability of the findings. First, our sample size was relatively limited, especially when comparing individuals using BZRA and those using BZDs. Secondly, the use of BZD/BZRA was determined through self-reporting, which may not accurately represent actual medication consumption as measured by objective assessments. The potential discrepancy could result in inaccuracies in our analysis of the dosage and duration of sedative-hypnotic use. Third, we were unable to examine sex differences among our participants due to the predominance of females over males. Understanding these differences is essential, as they can influence sleep needs and responses to treatments. Fourth, due to the light off/on protocol differences between the projects, we did not report total sleep time in minutes. Indeed, the strict wake-up time in one of the projects might have hindered Group effect on sleep duration. Finally, the data is cross-sectional and future studies should investigate sleep both before and after the use of sedative-hypnotics.

In conclusion, chronic use of sedative-hypnotics in older adults worsened sleep architecture and spectral activity compared to older adults both with and without insomnia disorder. The use of these drugs also altered the characteristics of sleep-related brain oscillations and their synchrony. The current findings may provide insight into the observed impact of chronic use of sedative-hypnotics on health outcomes, including cognitive function.

## Data Availability

All data produced in the present study are available upon reasonable request to the authors

## ACKNOWLEDGMENTS

This research was supported by grants from the Canadian Institutes of Health Research (MOP 142191, PJT 153115) to TDV and JPG, from the Natural Sciences and Engineering Research Council of Canada to TDV, from the Centre de Recherche de l’Institut Universitaire de Gériatrie de Montréal and from Concordia University to TDV. LB has been supported by the CIHR-SPOR Chair in Innovative, Patient-Oriented, Behavioural Clinical Trials, Concordia University. We acknowledge the contributions of the following students who assisted in participants’ recruitment, data collection and data preprocessing: Jean-Louis Zhao, Romain Perera, Ali Salimi, Cristina Bata, Jennifer Suliteanu, Kazem Habibi, Brian Hodhod, Alex Hillcoat, Kajamathy Subramaniam, Elizaveta Frolova, Emma-Maria Phillips, Rachel Hu, Loren Bies, Meaghan Pawlowski, Aminata Baldé, Alexandros Hadjinicolaou, Elissa Pierre, Laurence Vo Buu, Maryam Aboutiman, Shira Azoulay, Corina Lazarenco, Julia Lumia, Laurianne Bastien, Kenza Eddebbarh, Laurie Truchon, Rosette Tamaddon, Katherine Chhuon, Marissa Likoudis, Sabrina Di Francesco, Meghan Couture, Ilyas Mameri-Arab, Edith San-Jose Jones, Despina Bolanis, Shant Donabedian, Samuel Gillman, Eric Lachapelle, Katherine Walker, Roxanne Carbone, Léa Homer, Julia Giraud, Lilia Seguin, Mathilde Reyt, and all the volunteers. We also thank our sleep technologists Madeline Dickson and Elinah Mozhentiy from the Clinique SomnoMed for their contribution to the setup of sleep recordings. Finally, we would like to thank the participants for giving their time and energy to this research study.

## AUTHORS CONTRIBUTION

Conceptualization: LB, AAP, TDV, NEC

Recruitment: LB, ME, OMW, DC, CD, LT

Data collection: LB, ME, OMW, AAP, FBP, LT, MM, AM, DS, KG, JOB

Project coordination: LB, ME, AAP, OMW, LT, FBP

Data curation: LB, AAP, NEC, ME, OMW, LK, MM, AM, DS, VY, MM, KG

Formal analysis: LB Methodology: NEC, AAP, JOB

Visualization: LB, AAP, NEC

Interpretation: LB, AAP, NEC, TDV

Supervision: AAP, NEC, TDV

Funding: TDV, JPG

Writing - original draft: LB, AAP, NEC, TDV

Editing and reviewing: OMW, JPG, CT, SG, DL, CD, FA, JOB, FBP, ME, LT, MM, AM, DS, KG

## ABBREVIATIONS

AASM: American Academy of Sleep Medicine

AHI: Apnea-hypopnea index

BZD: Benzodiazepine

BZRA: Benzodiazepine Receptor Agonist or Z-drugs

CBTi: Cognitive-behavioral therapy for insomnia

CP: Absolute coupling phase distance from SO up-state

ECG: Electrocardiogram

EEG: Electroencephalogram

EMG: Electromyogram

EOG: Electrooculogram

GABA: Gamma-aminobutyric acid

GS: group involving good sleepers

Hz: Hertz

ICSD: International Classification of Sleep Disorder

INS: group involving participant with insomnia

ISI: Insomnia severity index

MED: group involving participant with insomnia and chronic sedative-hypnotic use

MI: Modulation index

MINI: Mini-international neuropsychiatric interview

MMSE: Mini-mental state examination

MoCA: Montreal cognitive assessment

NREM: Non-rapid-eye-movement

N1: NREM stage 1

N2: NREM stage 2

N3: NREM stage 3

PLM: Periodic limb movement

PSG: Polysomnography

REM: Rapid-eye-movement

SD: Standard deviation

SE: Sleep efficiency

sec: Second(s)

SFI: Sleep fragmentation index

SO: Slow oscillation

SL: Sleep latency

SOL: Sleep onset latency

SSI: Stage switch index

TIB: Time spent in bed

TST: Total sleep time

WASO: Wake after sleep onset

μV: microvolts

