## Supplementary materials for "Effect of chronic sedative-hypnotic use on sleep architecture and brain oscillations in older adults with chronic insomnia"

Barbaux et al.

#### Table des matières


**Table S1 – Dosage and type of sedatives-hypnotics used** 3

**Table S2: Age effect on sleep architecture** 3

**Table S3: Age effect on spectral activity during NREM** 4

**Table S4: Age effect on SOs and spindles, their association and SO-sigma PAC during NREM** 5

**Figure S1: Insomnia Severity Index score Distribution Across Groups** 6

**Chronic use of sedative-hypnotics affects sleep architecture.** 6

**Table S5 – Chronic sedative-hypnotic use affects spectral activity during NREM** 7

**Table S6 – Chronic sedative-hypnotic use affects spectral activity during REM** 8

**Chronic use of sedative-hypnotics alters spectral activity** 8

**Table S7 – Chronic sedative-hypnotic use affects SOs and spindles during NREM** 9

**Table S8: Chronic sedatives-hypnotic use affects SOs and spindles during N2 and N3** 9

**Table S9 – Effect of chronic sedative-hypnotic use on spindles using Moelle, Ray and Lacourse detection algorithms during NREM** 10

**Table S10 – Effect of chronic sedative-hypnotic use on SOs using Staresina and Massimini detection algorithms during NREM** 10

**Table S11 – Chronic sedative-hypnotic use affects SO-spindle association and SO-sigma PAC during NREM** 11

**Table S12 : Association between the Modulation Index and changes in SOs and spindles characteristics during NREM** 11

**Figure S2: Chronic sedative-hypnotic use is associated with alteration in the modulation index** 12

**Subgroup analysis: effect on the use and type of chronic sedative-hypnotic exposure on sleep.** 12

**Table S13 – Effect of chronic sedative-hypnotic exposure on sleep architecture** 13

**Table S14 – Effect of chronic sedative-hypnotic exposure on spectral activity during NREM** 14

**Table S15 – Effect of chronic sedative-hypnotic exposure on SOs and spindles during NREM** 14

**Table S16 – Effect of chronic sedative-hypnotic exposure on SO-spindle association and SO-sigma PAC during NREM** 15

**Table S17 – Effect of chronic BZRA use on sleep architecture** 15

**Table S18 – Effect of chronic BZRA use on spectral activity during NREM** 16

**Table S19 – Effect of chronic BZRA use on SOs and spindles, their co-occurrence and SO-sigma PAC during NREM** 16

**Chronic use of BZRA alters sleep quality** 17

#### Table S1 – Dosage and type of sedatives-hypnotics used

| Medication type | Dose equivalent in Diazepam (mg/ week) | | N |
|---|---|---|---|
| BZD | | 18 | |
| **Diazepam** | **10** | | 1 |
| Clonazepam | 0.5 | | 2 |
| Nitrazepam | 10 | | 2 |
| Oxazepam | 20 | | 6 |
| Lorazepam | 1 | | 6 |
| Temazepam | 20 | | 1 |
| BZRA | | 29 | |
| Zopiclone | 15 | | 29 |

BZD, benzodiazepine; BZRA, benzodiazepine receptor agonist

#### Table S2: Age effect on sleep architecture

| AGE EFFECT | N = 101 GS gp: n=28; INS gp: n=26; MED gp: n=47 | | | | | | N = 79 GS gp: n=28; INS gp: n=25; BZRA gp: n=26 | | | | | |
|---|---|---|---|---|---|---|---|---|---|---|---|---|
| | Spearman correlation test or ANCOVA | | | | | | Spearman correlation test or ANCOVA | | | | | |
| Oucome measure | GS vs INS | | INS vs MED | | GS vs MED | | GS vs INS | | INS vs BZRA | | GS vs MED | |
| | *p* | *r* | *p* | *r* | *p* | *r* | *p* | *r* | *p* | *r* | *p* | *r* |
| Sleep architecture | | | | | | | | | | | | |
| ***Sleep duration*** | | | | | | | | | | | | |
| TSP (% TIB) | **0.002** | -0.40 | 0.87 | - | 0.14 | - | **0.004** | -0.39 | 0.53 | - | **0.01** | -0.35 |
| SE (%) | **0.01** | -0.33 | 0.47 | - | **0.001** | -0.43 | **0.02** | -0.31 | 0.64 | - | **0.001** | -0.57 |
| N1 (% TSP) | 0.14 | - | 0.11 | - | **0.005** | 0.32 | 0.14 | - | 0.21 | - | **0.001** | 0.42 |
| N2 (% TSP) | ***F* (2,97)=22.5; *p* =0.01** | | | | | | ***F* (2,75)=18.8; *p* <0.001** | | | | | |
| N3 (% TSP) | **0.02** | -0.32 | **0.001** | -0.42 | **0.001** | -0.49 | ***F* (2,75)=17.9; *p* <0.001** | | | | | |
| REM (% TSP) | ***F* (2,97)=9.7; *p* =0.002** | | | | | | ***F* (2,75)=12.3; *p* <0.001** | | | | | |
| Wake (% TSP) | 0.06 | - | 0.30 | - | **0.001** | 0.46 | 0.09 | - | 0.83 | - | **0.001** | 0.54 |
| ***Sleep Initiation*** (min) | | | | | | | | | | | | |
| SOL | 0.06 | - | 0.53 | - | 0.20 | - | 0.11 | - | 0.63 | - | 0.15 | - |
| SL to N2 | 0.14 | - | 0.44 | - | 0.18 | - | 0.25 | - | 0.52 | - | 0.16 | - |
| SL to N3 | 0.10 | - | 0.21 | - | **0.01** | 0.28 | 0.15 | - | 0.60 | - | **0.04** | 0.27 |
| SL to REM | 0.79 | - | 0.24 | - | 0.07 | - | 0.79 | - | 0.60 | - | 0.17 | - |
| ***Sleep Fragmentation*** | | | | | | | | | | | | |
| WASO (% TIB) | 0.08 | - | 0.27 | - | **0.001** | 0.45 | 0.11 | - | 0.85 | - | **0.001** | 0.52 |
| Arousal density (Nber/30 sec) | | | | | | | | | | | | |
| in NREM+REM | *F* (2,97)=0.25; *p* =0.62 | | | | | | 0.25 | - | 0.90 | - | 0.85 | - |
| in NREM | 0.19 | - | 0.51 | - | 0.99 | - | 0.31 | - | 0.90 | - | 0.8 | - |
| in REM | 0.13 | - | 0.43 | - | 0.81 | - | 0.21 | - | 0.58 | - | 0.35 | - |
| SFI | **0.02** | 0.30 | 0.18 | - | **0.02** | 0.26 | **0.05** | 0.30 | 0.07 | - | **0.001** | 0.45 |
| SSI | **0.05** | 0.27 | 0.42 | - | 0.21 | - | **0.07** | - | 0.17 | - | **0.01** | 0.35 |

TSP, total sleep period; TIB, time in bed; SOL, sleep onset latency; SL, sleep latency; WASO, wake after sleep onset; SE, sleep efficiency; SFI, sleep fragmentation index; SSI, stage switch index; NREM, non-rapid eye movement; REM, rapid eye movement

Table S3: Age effect on spectral activity during NREM

| AGE EFFECT | N = 101 GS gp: n=28; INS gp: n=26; MED gp: n=47 | | | | | | N = 79 GS gp: n=28; INS gp: n=25; BZRA gp: n=26 | | | | | |
|---|---|---|---|---|---|---|---|---|---|---|---|---|
| Oucome measure | Spearman correlation test or ANCOVA | | | | | | Spearman correlation test or ANCOVA | | | | | |
| | GS vs INS | | INS vs MED | | GS vs MED | | GS vs INS | | INS vs BZRA | | GS vs MED | |
| | *p* | *r* | *p* | *r* | *p* | *r* | *p* | *r* | *p* | *r* | *p* | *r* |
| ***Relative spectrum*** | | | | | | | | | | | | |
| **Fz** | | | | | | | | | | | | |
| SO | 0.29 | - | 0.72 | - | 0.75 | - | 0.39 | - | 0.70 | - | 0.62 | - |
| Delta | 0.18 | - | **0.02** | -0.28 | **0.001** | -0.36 | 0.15 | - | 0.10 | - | **0.01** | -0.33 |
| Theta | 0.42 | - | 0.79 | - | 0.29 | - | *F* (2,75)=0.07; *p* =0.80 | | | | | |
| Alpha | 0.22 | - | 0.20 | - | 0.77 | - | 0.23 | - | 0.79 | - | 0.51 | - |
| Sigma | 0.87 | - | **0.003** | 0.34 | 0.05 | - | 0.748 | - | **0.04** | 0.28 | 0.52 | - |
| Low Beta | 0.07 | - | **<0.001** | 0.42 | **0.006** | 0.31 | 0.05 | - | **0.007** | 0.37 | 0.10 | - |
| High Beta | 0.06 | - | **0.002** | 0.36 | **0.009** | 0.30 | **0.038** | 0.28 | **0.02** | 0.32 | 0.09 | - |
| **Pz** | | | | | | | | | | | | |
| SO | *F* (2,97)=0.16; *p* =0.68 | | | | | | 0.87 | - | 0.98 | - | 0.59 | - |
| Delta | *F* (2,97)=2.5; *p* =0.12 | | | | | | *F* (2,75)=1.55; *p* =0.22 | | | | | |
| Theta | 0.69 | - | 0.64 | - | 0.19 | - | *F* (2,75)=0.35; *p* =0.56 | | | | | |
| Alpha | 0.89 | - | 0.097 | - | 0.79 | - | 0.82 | - | 0.43 | - | 0.64 | - |
| Sigma | 0.46 | - | **0.013** | 0.28 | 0.307 | - | 0.42 | - | 0.13 | - | 0.88 | - |
| Low Beta | 0.08 | - | **0.002** | 0.35 | **0.05** | - | *F* (2,75)=1.24; *p* =0.27 | | | | | |
| High Beta | **0.03** | 0.29 | **<0.001** | 0.40 | **0.01** | 0.29 | **0.03** | 0.30 | **0.02** | 0.31 | 0.12 | - |
| ***Index (/Beta)*** | | | | | | | | | | | | |
| **Fz** | | | | | | | | | | | | |
| SO | 0.10 | - | **0.001** | -0.37 | **0.01** | -0.29 | 0.054 | - | **0.01** | -0.34 | 0.106 | - |
| Delta | **0.04** | -0.27 | **<0.001** | -0.40 | **0.004** | -0.33 | **0.03** | -0.30 | **0.009** | -0.36 | 0.06 | - |
| Theta | **0.03** | -0.29 | **0.003** | -0.34 | **0.002** | -0.35 | **0.03** | -0.29 | 0.12 | - | **0.03** | -0.30 |
| Alpha | **0.003** | -0.39 | 0.06 | - | **0.03** | -0.24 | **0.002** | -0.41 | 0.10 | - | **0.04** | -0.27 |
| Sigma | **0.009** | -0.35 | 0.82 | - | 0.18 | - | **0.01** | -0.37 | 0.94 | - | 0.18 | - |
| Low Beta | *F* (2,97)=0.3; *p* =0.60 | | | | | | *F* (2,75)=2.16; *p* =0.15 | | | | | |
| High Beta | *F* (2,97)=0.3; *p* =0.61 | | | | | | *F* (2,75)=2.16; *p* =0.15 | | | | | |
| **Pz** | | | | | | | | | | | | |
| SO | 0.06 | - | **0.002** | -0.35 | 0.05 | - | 0.07 | - | **0.04** | -0.29 | 0.26 | - |
| Delta | 0.06 | - | **0.001** | -0.38 | **0.02** | -0.27 | 0.06 | - | **0.03** | -0.30 | 0.17 | - |
| Theta | **0.04** | -0.27 | **0.002** | -0.35 | **0.003** | -0.34 | **0.04** | -0.27 | 0.08 | - | **0.04** | -0.28 |
| Alpha | **0.04** | -0.27 | 0.18 | - | 0.09 | - | **0.04** | -0.28 | 0.28 | - | 0.10 | - |
| Sigma | 0.14 | - | 0.91 | - | 0.21 | - | 0.20 | - | 0.91 | - | 0.08 | - |
| Low Beta | 0.22 | - | 0.80 | - | 0.27 | - | 0.15 | - | 0.99 | - | 0.16 | - |
| High Beta | 0.22 | - | 0.80 | - | 0.27 | - | 0.15 | - | 0.99 | - | 0.16 | - |

SO, slow oscillation

Table S4: Age effect on SOs and spindles, their association and SO-sigma PAC during NREM

| AGE EFFECT | N = 101 GS gp: n=28; INS gp: n=26; MED gp: n=47 | | | | | | N = 79 GS gp: n=28; INS gp: n=25; BZRA gp: n=26 | | | | | |
|---|---|---|---|---|---|---|---|---|---|---|---|---|
| Oucome measure | Spearman correlation test or ANCOVA | | | | | | Spearman correlation test or ANCOVA | | | | | |
| | GS vs INS | | INS vs MED | | GS vs MED | | GS vs INS | | INS vs BZRA | | GS vs MED | |
| | *p* | *r* | *p* | *r* | *p* | *r* | *p* | *r* | *p* | *r* | *p* | *r* |
| ***Spindle characteristics (Moelle detection)*** | | | | | | | | | | | | |
| **Fz** | | | | | | | | | | | | |
| Density (Nber/30 sec) | 0.16 | - | 0.72 | - | 0.77 | - | *F* (2,75)=1.1; *p* =0.29 | | | | | |
| Duration (sec) | 0.42 | - | 0.76 | - | 0.80 | - | 0.39 | - | 0.94 | - | 0.45 | - |
| Amplitude (µV) | **0.04** | -0.27 | **0.001** | -0.38 | **0.001** | -0.36 | **0.03** | -0.29 | **0.01** | -0.35 | **0.02** | -0.33 |
| Frequency (Hz) | *F* (2,97)=0.01; *p* =0.94 | | | | | | *F* (2,75)=0.11; *p* =0.74 | | | | | |
| **Pz** | | | | | | | | | | | | |
| Density (Nber/30 sec) | 0.92 | - | 0.94 | - | 0.67 | - | *F* (2,75)=0.05; *p* =0.83 | | | | | |
| Duration (sec) | 0.52 | - | 0.855 | - | 0.57 | - | *F* (2,75)=0.01; *p* =0.93 | | | | | |
| Amplitude (µV) | **0.02** | -0.32 | **0.002** | -0.35 | **0.001** | -0.36 | ***F* (2,75)=10.1; *p* =0.002** | | | | | |
| Frequency (Hz) | ***F* (2,97)=4.6; *p* =0.03** | | | | | | 0.052 | - | 0.007 | -0.37 | 0.009 | -0.35 |
| ***SO characteristics (Staresina detection)*** | | | | | | | | | | | | |
| **Fz** | | | | | | | | | | | | |
| Density (Nber/30 sec) | 0.38 | - | **0.04** | -0.23 | **0.04** | -0.23 | *F* (2,75)=3.5; *p* =0.07 | | | | | |
| Duration (sec) | 0.859 | - | **0.01** | 0.29 | **0.001** | 0.37 | 0.98 | - | 0.09 | - | **0.02** | 0.32 |
| Amplitude (µV) | **0.02** | -0.32 | **<0.001** | -0.42 | **<0.001** | -0.43 | ***F* (2,75)=13.7; *p<0.001*** | | | | | |
| Frequency (Hz) | 0.29 | - | **0.01** | -0.28 | **<0.001** | -0.47 | 0.19 | - | 0.18 | - | **0.001** | -0.45 |
| **Pz** | | | | | | | | | | | | |
| Density (Nber/30 sec) | 0.19 | - | 0.24 | - | 0.08 | - | 0.14 | - | 0.11 | - | **0.04** | -0.28 |
| Duration (sec) | 0.17 | - | 0.11 | - | 0.07 | - | 0.08 | - | 0.13 | - | 0.14 | - |
| Amplitude (µV) | **0.002** | -0.40 | **0.001** | -0.38 | **<0.001** | -0.39 | ***F* (2,75)=12.8; *p<0.001*** | | | | | |
| Frequency (Hz) | 0.33 | - | **0.01** | -0.28 | **0.009** | -0.29 | 0.18 | - | **0.02** | -0.32 | **0.04** | -0.2 |
| ***Temporal association*** | | | | | | | | | | | | |
| **Fz** | | | | | | | | | | | | |
| Recall | 0.38 | - | 0.46 | - | 0.58 | - | *F* (2,75)=0.02; *p* =0.89 | | | | | |
| Precision | *F* (2,97)=0.14; *p* =0.71 | | | | | | *F* (2,75)=0.02; *p* =0.88 | | | | | |
| **Pz** | | | | | | | | | | | | |
| Recall | 0.87 | - | 0.40 | - | 0.19 | - | 0.84 | - | 0.21 | - | **0.04** | -0.28 |
| Precision | *F* (2,97)=3.2; *p* =0.08 | | | | | | ***F* (2,75)=4.3; *p* =0.04** | | | | | |
| ***SO-sigma PAC*** | | | | | | | | | | | | |
| **Fz** | | | | | | | | | | | | |
| Modulation Index | 0.13 | - | 0.76 | - | 0.55 | - | 0.12 | - | 0.97 | - | 0.91 | - |
| **Pz** | | | | | | | | | | | | |
| Modulation Index | 0.47 | - | **0.001** | -0.38 | **0.003** | -0.33 | 0.34 | - | **0.008** | -0.36 | **0.04** | -0.28 |

SO, slow oscillation; PAC, phase-amplitude coupling

#### Figure S1: Insomnia Severity Index score Distribution Across Groups

Asterisks represent significance (*p*): *<0.05; **<0.01; ***<0.001

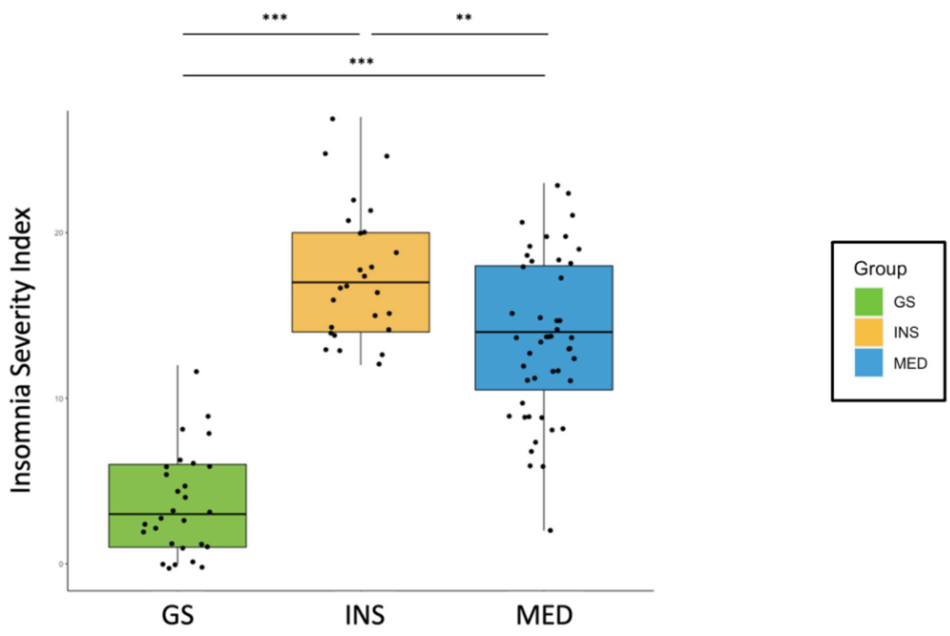


#### Chronic use of sedative-hypnotics affects sleep architecture.

There was no significant difference in TSP $F(2,97)=.87$, $p=.7$), both MED and INS groups spent more time awake (WASO) compared to GS $F(2,97)=15.7$, $p<.001$, $q=.001$) (all $p \leq .001$, all $q<.002$). We found a Group effect ($F(2,97)=20.2$, $p<.001$) where both GS ($p=.03$, $q=.08$) and MED ($p<.0001$) presented greater SFI compared to the INS group. Furthermore, we found a Group effect ($F(2,97)=7.2$, $p=.001$, $q=.002$) where both the GS and MED groups shown lower total arousal density compared to the INS (all $p \leq .004$, $q \leq .01$) group. The same Group effect was found when taking arousal density in NREM and REM individually.

#### Table S5 – Chronic sedative-hypnotic use affects spectral activity during NREM

| Outcome measure | GS | | INS | | MED | | GS vs INS vs MED Kruskal-Wallis test or ANOVA | | | | | GS vs INS post hoc - Dunn test or t t | | | effect siz | INS vs MED post hoc - Dunn test or t test | | | effect size | GS vs MED post hoc - Dunn test or t test | | | effect size |
|---|---|---|---|---|---|---|---|---|---|---|---|---|---|---|---|---|---|---|---|---|---|---|---|
| | Mean | SD | Mean | SD | Mean | SD | F | df | residual | *p* | *q* | *t* | *p* | *q* | g' | *t* | *p* | *q* | g' | *t* | *p* | *q* | g' |
| ***Relative spectrum*** | | | | | | | | | | | | | | | | | | | | | | | |
| **Fz** | | | | | | | | | | | | | | | | | | | | | | | |
| SO | 0.59 | 0.1 | 0.59 | 0.1 | 0.61 | 0.1 | 1.68 | 2 | 97 | 0.43 | - | - | - | - | - | - | - | - | - | - | - | - | - |
| Delta | 0.76 | 0.07 | 0.78 | 0.08 | 0.74 | 0.08 | 5.17 | 2 | 97 | 0.08 | - | - | - | - | - | - | - | - | - | - | - | - | - |
| Theta | 0.08 | 0.03 | 0.07 | 0.03 | 0.06 | 0.03 | 11.80 | 2 | 97 | 0.003 | **0.005*** | 1.01 | 0.31 | - | 0.21 | 2.09 | **0.036** | 0.11 | 0.51 | 3.3 | 0.001 | **0.003*** | 0.77 |
| Alpha | 0.04 | 0.02 | 0.03 | 0.02 | 0.05 | 0.03 | 2.42 | 2 | 97 | 0.29 | - | - | - | - | - | - | - | - | - | - | - | - | - |
| Sigma | 0.02 | 0.01 | 0.02 | 0.01 | 0.03 | 0.02 | 15.30 | 2 | 97 | <0.001 | **0.001*** | -0.29 | 0.77 | - | -0.11 | -3.02 | 0.003 | **0.008*** | -0.82 | -3.42 | 0.001 | **0.002*** | -0.89 |
| Low Beta | 0.02 | 0.008 | 0.018 | 0.009 | 0.025 | 0.025 | 18.43 | 2 | 97 | <0.002 | **0.001*** | -0.18 | 0.86 | - | -0.09 | -3.4 | 0.001 | **0.002*** | -0.68 | -3.68 | **<0.001*** | - | -0.75 |
| High Beta | 0.03 | 0.017 | 0.033 | 0.017 | 0.045 | 0.04 | 16.99 | 2 | 97 | <0.003 | **0.001*** | 0.28 | 0.78 | - | 0.08 | -3.52 | <0.001 | **0.001*** | -0.76 | -3.28 | 0.001 | **0.003*** | -0.72 |
| **Cz** | | | | | | | | | | | | | | | | | | | | | | | |
| SO | 0.52 | 0.11 | 0.53 | 0.11 | 0.54 | 0.09 | 0.88 | 2 | 97 | 0.64 | - | - | - | - | - | - | - | - | - | - | - | - | - |
| Delta | 0.73 | 0.07 | 0.73 | 0.08 | 0.71 | 0.07 | 0.06 | 2 | 97 | 0.94 | - | - | - | - | - | - | - | - | - | - | - | - | - |
| Theta | 0.1 | 0.03 | 0.09 | 0.03 | 0.08 | 0.03 | 9.54 | 2 | 97 | 0.008 | **0.019*** | 0.48 | 0.63 | - | 0.11 | 2.2 | **0.03** | 0.08 | 0.59 | 2.83 | 0.005 | **0.01*** | 0.7 |
| Alpha | 0.04 | 0.02 | 0.04 | 0.02 | 0.05 | 0.03 | 0.89 | 2 | 97 | 0.64 | - | - | - | - | - | - | - | - | - | - | - | - | - |
| Sigma | 0.03 | 0.01 | 0.03 | 0.01 | 0.04 | 0.02 | 8.83 | 2 | 97 | 0.012 | **0.021*** | 0.09 | 0.92 | - | 0.03 | -2.47 | **0.04*** | - | -0.71 | -2.46 | 0.01 | **0.04*** | -0.69 |
| Low Beta | 0.005 | 0.002 | 0.005 | 0.002 | 0.007 | 0.003 | 14.05 | 2 | 97 | <0.001 | **0.003*** | -0.54 | 0.58 | - | -0.13 | -2.71 | **0.02*** | - | -0.73 | -3.42 | 0.001 | **0.002*** | -0.84 |
| High Beta | 0.009 | 0.004 | 0.009 | 0.004 | 0.013 | 0.006 | 14.63 | 2 | 97 | <0.002 | **0.003*** | -0.01 | 0.99 | - | 0 | -3.11 | **0.006*** | - | -0.77 | -3.23 | 0.001 | **0.004*** | -0.77 |
| **Pz** | | | | | | | | | | | | | | | | | | | | | | | |
| SO | 0.56 | 0.12 | 0.57 | 0.11 | 0.6 | 0.12 | *1.35* | 2 | 97 | *0.26* | - | - | - | - | - | - | - | - | - | - | - | - | - |
| Delta | 0.69 | 0.08 | 0.72 | 0.1 | 0.69 | 0.08 | *0.73* | 2 | 97 | *0.48* | - | - | - | - | - | - | - | - | - | - | - | - | - |
| Theta | 0.11 | 0.05 | 0.1 | 0.05 | 0.07 | 0.04 | 15.65 | 2 | 97 | <0.001 | **0.002*** | 1.23 | 0.22 | - | 0.32 | 2.35 | **0.019** | 0.055 | 0.63 | 3.82 | **<0.001*** | - | 1 |
| Alpha | 0.05 | 0.02 | 0.04 | 0.03 | 0.04 | 0.03 | 1.09 | 2 | 97 | 0.58 | - | - | - | - | - | - | - | - | - | - | - | - | - |
| Sigma | 0.04 | 0.02 | 0.04 | 0.02 | 0.05 | 0.04 | 3.04 | 2 | 97 | 0.22 | - | - | - | - | - | - | - | - | - | - | - | - | - |
| Low Beta | 0.004 | 0.002 | 0.004 | 0.002 | 0.006 | 0.004 | 5.75 | 2 | 97 | 0.06 | - | - | - | - | - | - | - | - | - | - | - | - | - |
| High Beta | 0.009 | 0.004 | 0.007 | 0.007 | 0.01 | 0.007 | 8.74 | 2 | 97 | 0.013 | **0.044*** | 0.89 | 0.37 | - | 0.29 | -2.81 | 0.005 | **0.01*** | -0.68 | -1.87 | 0.06 | - | -0.48 |
| ***Index (/Beta)*** | | | | | | | | | | | | | | | | | | | | | | | |
| **Fz** | | | | | | | | | | | | | | | | | | | | | | | |
| SO | 80.27 | 61.4 | 76.57 | 42.93 | 53.66 | 44.27 | 12.05 | 2 | 97 | 0.002 | **0.006*** | -0.37 | 0.71 | - | 0.07 | 3.03 | 0.002 | **0.007*** | 0.52 | 2.68 | 0.007 | **0.022*** | 0.51 |
| Delta | 97.15 | 49.52 | 98.81 | 52.13 | 62.88 | 38.31 | 17.87 | 2 | 97 | **<0.001*** | - | -0.22 | 0.82 | - | -0.03 | 3.57 | **<0.001*** | - | 0.81 | 3.41 | 0.001 | **0.002*** | 0.79 |
| Theta | 9.35 | 4.03 | 9.15 | 6.37 | 4.63 | 3.2 | 32.95 | 2 | 97 | **<0.001*** | - | 0.88 | 0.37 | - | 0.04 | 4.12 | **<0.001*** | - | 0.98 | 5.22 | **<0.001*** | - | 1.32 |
| Alpha | 4.78 | 4.64 | 4.16 | 2.6 | 3.37 | 2.51 | 5.49 | 2 | 97 | 0.06 | - | - | - | - | - | - | - | - | - | - | - | - | - |
| Sigma | 2.11 | 0.74 | 2.17 | 0.64 | 2.19 | 0.79 | 0.09 | 2 | 97 | 0.96 | - | - | - | - | - | - | - | - | - | - | - | - | - |
| Low Beta | 0.33 | 0.04 | 0.35 | 0.03 | 0.34 | 0.05 | 0.63 | 2 | 97 | 0.53 | - | - | - | - | - | - | - | - | - | - | - | - | - |
| High Beta | 0.67 | 0.04 | 0.65 | 0.03 | 0.66 | 0.05 | 0.63 | 2 | 97 | 0.53 | - | - | - | - | - | - | - | - | - | - | - | - | - |
| **Cz** | | | | | | | | | | | | | | | | | | | | | | | |
| SO | 51.39 | 40.2 | 47.03 | 29.33 | 33.43 | 18 | 8.53 | 2 | 97 | 0.01 | **0.03*** | -0.09 | 0.93 | - | 0.12 | 2.43 | 0.02 | **0.045*** | 0.59 | 2.42 | 0.016 | **0.046*** | 0.62 |
| Delta | 66.01 | 32.41 | 61.46 | 25.46 | 44.36 | 24.9 | 13.91 | 2 | 97 | 0.001 | **0.003*** | 0.2 | 0.84 | - | 0.15 | 2.92 | 0.004 | **0.01*** | 0.67 | 3.25 | 0.001 | **0.004*** | 0.76 |
| Theta | 8.13 | 3.24 | 7.77 | 3.93 | 4.72 | 2.94 | 27.67 | 2 | 97 | **<0.001*** | - | 0.55 | 0.58 | - | 0.1 | 3.94 | **<0.001*** | - | 0.91 | 4.71 | **<0.001*** | - | 1.1 |
| Alpha | 3.59 | 2.34 | 3.4 | 1.7 | 2.85 | 2.17 | 6.43 | 2 | 97 | **0.04** | 0.07 | -0.03 | 0.97 | - | 0.09 | 2.08 | **0.04** | 0.11 | 0.27 | 2.12 | **0.03** | 0.10 | 0.33 |
| Sigma | 2.41 | 0.89 | 2.33 | 0.68 | 2.21 | 0.6 | 0.33 | 2 | 97 | 0.84 | - | - | - | - | - | - | - | - | - | - | - | - | - |
| Low Beta | 0.35 | 0.04 | 0.36 | 0.04 | 0.34 | 0.05 | 0.51 | 2 | 97 | 0.60 | - | - | - | - | - | - | - | - | - | - | - | - | - |
| High Beta | 0.65 | 0.04 | 0.64 | 0.04 | 0.66 | 0.05 | 0.51 | 2 | 97 | 0.60 | - | - | - | - | - | - | - | - | - | - | - | - | - |
| **Pz** | | | | | | | | | | | | | | | | | | | | | | | |
| SO | 57.64 | 45.81 | 61.63 | 45 | 51.74 | 51.06 | 4.13 | 2 | 97 | 0.13 | - | - | - | - | - | - | - | - | - | - | - | - | - |
| Delta | 65.86 | 33.86 | 73.26 | 34.67 | 54.8 | 35.6 | 7.62 | 2 | 97 | 0.02 | **0.044*** | -0.85 | 0.39 | - | -0.21 | 2.63 | 0.008 | **0.03*** | 0.52 | 1.73 | 0.08 | - | 0.31 |
| Theta | 9.23 | 3.27 | 9.04 | 5.28 | 4.84 | 3.13 | 34.78 | 2 | 97 | **<0.001*** | - | 0.86 | 0.38 | - | 0.04 | 4.26 | **<0.001*** | - | 1.03 | 5.35 | **<0.001*** | - | 1.37 |
| Alpha | 3.66 | 1.69 | 3.72 | 2.71 | 2.75 | 2.17 | 12.88 | 2 | 97 | 0.002 | **0.005*** | 0.44 | 0.65 | - | -0.03 | 2.65 | 0.008 | **0.02*** | 0.4 | 3.22 | 0.001 | **0.004*** | 0.45 |
| Sigma | 3.1 | 1.25 | 3.01 | 0.96 | 2.82 | 1 | 0.28 | 2 | 97 | 0.87 | - | - | - | - | - | - | - | - | - | - | - | - | - |
| Low Beta | 0.35 | 0.04 | 0.37 | 0.04 | 0.34 | 0.06 | 6.57 | 2 | 97 | 0.037 | **0.05*** | -1.57 | 0.11 | - | -0.52 | 2.56 | 0.01 | **0.03*** | 0.5 | 0.83 | 0.40 | - | 0.09 |
| High Beta | 0.65 | 0.04 | 0.63 | 0.04 | 0.66 | 0.06 | 6.57 | 2 | 97 | 0.037 | **0.05*** | 1.57 | 0.11 | - | 0.52 | -2.56 | 0.01 | **0.03*** | -0.5 | -0.83 | 0.40 | - | -0.09 |

SO, slow oscillation

#### Table S6 – Chronic sedative-hypnotic use affects spectral activity during REM

| Outcome measure | GS | | INS | | MED | | GS vs INS vs MED<br>Kruskal-Wallis test or ANOVA | | | | | GS vs INS<br>post hoc - Dunn test or t test | | | effect size | INS vs MED<br>post hoc - Dunn test or t test | | | effect size | GS vs MED<br>post hoc - Dunn test or t test | | | effect size |
|---|---|---|---|---|---|---|---|---|---|---|---|---|---|---|---|---|---|---|---|---|---|---|---|
| | Mean | SD | Mean | SD | Mean | SD | F | df | residual | *p* | *q* | *t* | *p* | *q* | g' | *t* | *p* | *q* | g' | *t* | *p* | *q* | g' |
| ***Relative power spectrum*** | | | | | | | | | | | | | | | | | | | | | | | |
| **Fz** | | | | | | | | | | | | | | | | | | | | | | | |
| SO | 0.44 | 0.13 | 0.4 | 0.1 | 0.46 | 0.14 | 4.13 | 2 | 97 | 0.23 | - | - | - | - | - | - | - | - | - | - | - | - | - |
| Delta | 0.57 | 0.06 | 0.59 | 0.09 | 0.57 | 0.1 | 0.15 | 2 | 97 | 0.86 | - | - | - | - | - | - | - | - | - | - | - | - | - |
| Theta | 0.15 | 0.05 | 0.15 | 0.06 | 0.1 | 0.05 | 21.31 | 2 | 97 | **<0.001*** | - | 0.66 | 0.51 | - | 0.07 | 3.37 | 0.001 | **0.002*** | 0.91 | 4.16 | **<0.001*** | - | 1.09 |
| Alpha | 0.07 | 0.03 | 0.06 | 0.03 | 0.06 | 0.03 | 3.09 | 2 | 97 | 0.25 | - | - | - | - | - | - | - | - | - | - | - | - | - |
| Sigma | 0.04 | 0.02 | 0.04 | 0.02 | 0.05 | 0.02 | 3.38 | 2 | 97 | 0.25 | - | - | - | - | - | - | - | - | - | - | - | - | - |
| Low Beta | 0.01 | 0.01 | 0.02 | 0.01 | 0.03 | 0.03 | 6.15 | 2 | 97 | **0.046** | 0.11 | -0.6 | 0.55 | - | -0.35 | -1.63 | 0.10 | - | -0.38 | -2.33 | **0.02** | 0.05 | -0.51 |
| High Beta | 0.04 | 0.02 | 0.04 | 0.03 | 0.06 | 0.05 | 4.83 | 2 | 97 | 0.21 | - | - | - | - | - | - | - | - | - | - | - | - | - |
| **Cz** | | | | | | | | | | | | | | | | | | | | | | | |
| SO | 0.39 | 0.16 | 0.35 | 0.12 | 0.39 | 0.15 | 2.26 | 2 | 97 | 0.42 | - | - | - | - | - | - | - | - | - | - | - | - | - |
| Delta | 0.54 | 0.06 | 0.56 | 0.08 | 0.55 | 0.1 | 0.60 | 2 | 97 | *0.74* | - | - | - | - | - | - | - | - | - | - | - | - | - |
| Theta | 0.17 | 0.05 | 0.17 | 0.06 | 0.13 | 0.05 | 4.15 | 2 | 97 | **0.02** | *0.13* | -0.23 | 0.82 | - | -0.06 | 3.27 | <0.001 | **0.003*** | 0.84 | 3.27 | 0.001 | **0.004*** | 0.8 |
| Alpha | 0.07 | 0.03 | 0.07 | 0.03 | 0.07 | 0.04 | 2.04 | 2 | 97 | 0.42 | - | - | - | - | - | - | - | - | - | - | - | - | - |
| Sigma | 0.05 | 0.02 | 0.05 | 0.02 | 0.06 | 0.03 | 2.92 | 2 | 97 | 0.41 | - | - | - | - | - | - | - | - | - | - | - | - | - |
| Low Beta | 0.02 | 0.01 | 0.02 | 0.01 | 0.03 | 0.04 | 5.05 | 2 | 97 | *0.28* | - | - | - | - | - | - | - | - | - | - | - | - | - |
| High Beta | 0.04 | 0.02 | 0.04 | 0.02 | 0.06 | 0.04 | 3.70 | 2 | 97 | 0.37 | - | - | - | - | - | - | - | - | - | - | - | - | - |
| **Pz** | | | | | | | | | | | | | | | | | | | | | | | |
| SO | 0.42 | 0.16 | 0.4 | 0.13 | 0.47 | 0.15 | 4.36 | 2 | 97 | 0.11 | - | - | - | - | - | - | - | - | - | - | - | - | - |
| Delta | 0.5 | 0.08 | 0.54 | 0.1 | 0.55 | 0.11 | 2.25 | 2 | 97 | 0.11 | - | - | - | - | - | - | - | - | - | - | - | - | - |
| Theta | 0.16 | 0.05 | 0.15 | 0.07 | 0.1 | 0.05 | 20.84 | 2 | 97 | **<0.001*** | - | 0.57 | 0.57 | - | 0.07 | 3.38 | 0.001 | **0.002*** | 0.88 | 4.1 | **<0.001*** | - | 1.07 |
| Alpha | 0.1 | 0.06 | 0.09 | 0.05 | 0.07 | 0.05 | 5.77 | 2 | 97 | 0.06 | - | - | - | - | - | - | - | - | - | - | - | - | - |
| Sigma | 0.06 | 0.03 | 0.06 | 0.02 | 0.07 | 0.03 | 0.4 | 2 | 97 | 0.67 | - | - | - | - | - | - | - | - | - | - | - | - | - |
| Low Beta | 0.02 | 0.01 | 0.02 | 0.01 | 0.02 | 0.03 | 2.32 | 2 | 97 | 0.31 | - | - | - | - | - | - | - | - | - | - | - | - | - |
| High Beta | 0.03 | 0.02 | 0.03 | 0.02 | 0.05 | 0.04 | 2.13 | 2 | 97 | 0.34 | - | - | - | - | - | - | - | - | - | - | - | - | - |
| ***Beta ratio*** | | | | | | | | | | | | | | | | | | | | | | | |
| **Fz** | | | | | | | | | | | | | | | | | | | | | | | |
| SO | 11.18 | 8.06 | 9.02 | 5.6 | 9.63 | 8.13 | 1.31 | 2 | 97 | 0.51 | - | - | - | - | - | - | - | - | - | - | - | - | - |
| Delta | 13.64 | 6.74 | 12.45 | 5.58 | 11.75 | 10.14 | 5.69 | 2 | 97 | 0.06 | - | - | - | - | - | - | - | - | - | - | - | - | - |
| Theta | 3.53 | 2.14 | 3.15 | 1.96 | 1.82 | 1.42 | 23.53 | 2 | 97 | **<0.001*** | - | 0.89 | 0.37 | - | 0.19 | 3.4 | 0.001 | **0.002*** | 0.81 | 4.45 | **<0.001*** | - | 0.99 |
| Alpha | 1.53 | 1.02 | 1.3 | 0.92 | 0.97 | 0.67 | 9.33 | 2 | 97 | 0.009 | **0.03*** | 0.9 | 0.36 | - | 0.23 | 1.88 | 0.06 | - | 0.43 | 2.92 | 0.003 | **0.01*** | 0.68 |
| Sigma | 0.85 | 0.33 | 0.82 | 0.28 | 0.77 | 0.38 | 2.33 | 2 | 97 | 0.31 | - | - | - | - | - | - | - | - | - | - | - | - | - |
| Low Beta | 0.3 | 0.07 | 0.3 | 0.07 | 0.31 | 0.09 | 0.18 | 2 | 97 | 0.91 | - | - | - | - | - | - | - | - | - | - | - | - | - |
| High Beta | 0.7 | 0.07 | 0.7 | 0.07 | 0.69 | 0.09 | 0.18 | 2 | 97 | 0.91 | - | - | - | - | - | - | - | - | - | - | - | - | - |
| **Cz** | | | | | | | | | | | | | | | | | | | | | | | |
| SO | 11.12 | 17.26 | 7.83 | 6.18 | 7.66 | 6.48 | 1.29 | 2 | 97 | *0.62* | - | - | - | - | - | - | - | - | - | - | - | - | - |
| Delta | 12.21 | 7.99 | 11.43 | 5.35 | 10.33 | 8.2 | 4.17 | 2 | 97 | *0.29* | - | - | - | - | - | - | - | - | - | - | - | - | - |
| Theta | 3.44 | 1.85 | 3.31 | 1.77 | 2.13 | 1.43 | 15.89 | 2 | 97 | <0.001 | **0.002*** | 0.35 | 0.72 | - | 0.07 | 3.01 | 0.003 | **0.008*** | 0.75 | 3.53 | <0.001 | **0.001*** | 0.81 |
| Alpha | 1.53 | 0.9 | 1.38 | 0.73 | 1.07 | 0.67 | 7.95 | 2 | 97 | **0.02** | 0.06 | 0.62 | 0.53 | - | 0.18 | 1.87 | 0.19 | - | 0.44 | 2.65 | 0.008 | **0.02*** | 0.6 |
| Sigma | 0.92 | 0.31 | 0.91 | 0.26 | 0.87 | 0.37 | 1.29 | 2 | 97 | *0.62* | - | - | - | - | - | - | - | - | - | - | - | - | - |
| Low Beta | 0.3 | 0.05 | 0.31 | 0.06 | 0.32 | 0.09 | 0.94 | 2 | 97 | 0.62 | - | - | - | - | - | - | - | - | - | - | - | - | - |
| High Beta | 0.7 | 0.05 | 0.69 | 0.06 | 0.68 | 0.09 | 0.94 | 2 | 97 | 0.62 | - | - | - | - | - | - | - | - | - | - | - | - | - |
| **Pz** | | | | | | | | | | | | | | | | | | | | | | | |
| SO | 14.79 | 25.85 | 10.41 | 8.36 | 11.9 | 11.95 | 0.008 | 2 | 97 | 0.99 | - | - | - | - | - | - | - | - | - | - | - | - | - |
| Delta | 13.66 | 11.6 | 13.11 | 6.02 | 13.38 | 13.1 | 1.87 | 2 | 97 | 0.39 | - | - | - | - | - | - | - | - | - | - | - | - | - |
| Theta | 3.71 | 1.83 | 3.6 | 2.54 | 2.03 | 1.42 | 23.01 | 2 | 97 | **<0.001*** | - | 0.79 | 0.42 | - | 0.05 | 3.41 | 0.001 | **0.001*** | 0.82 | 4.39 | **<0.001*** | - | 1.05 |
| Alpha | 2.4 | 1.49 | 2.12 | 1.6 | 1.37 | 0.86 | 12.69 | 2 | 97 | **<0.001*** | - | 0.66 | 0.50 | - | 0.18 | 2.48 | 0.01 | **0.039*** | 0.63 | 3.29 | 0.001 | **0.003*** | 0.9 |
| Sigma | 1.33 | 0.51 | 1.29 | 0.51 | 1.12 | 0.47 | 4.69 | 2 | 97 | 0.09 | - | - | - | - | - | - | - | - | - | - | - | - | - |
| Low Beta | 0.33 | 0.05 | 0.35 | 0.05 | 0.35 | 0.07 | 1.04 | 2 | 97 | 0.59 | - | - | - | - | - | - | - | - | - | - | - | - | - |
| High Beta | 0.67 | 0.05 | 0.65 | 0.05 | 0.65 | 0.07 | 1.04 | 2 | 97 | 0.59 | - | - | - | - | - | - | - | - | - | - | - | - | - |

SO, slow oscillation

#### Chronic use of sedative-hypnotics alters spectral activity

In the frontal region (Fz), we found in frontal a group effect for SO- ($F$(2,97)=12.1, $p$=.002, $q$=.006), delta- ($F$(2,97)=17.9, $p$<.001) and theta-beta ratio ($F$(2,97)=32.9, $p$<.001), where both GS (all $p$≤.007, $q$≤.02) and INS (all $p$≤.002, $q$≤.007) were greater compared to MED group. In the parietal region (Pz), we found a group effect on delta- ($F$(2,97)=7.6, $p$=.02 $q$=.044), theta- ($F$(2,97)=34.5, $p$<.001), alpha- ($F$(2,97)=12.9, $p$=.002, $p$=.005), low and high (all $F$(2,97)=6.6, $p$=.037, $q$=.05) beta ratio, where the MED group displayed lower delta- ($p$=.008, $q$=.03) and low beta-ratio ($p$=.01, $q$=.03), as well as greater high-beta ratio ($p$=.01, $q$=.03) compared to INS and lower theta- and alpha-beta ratio compared to both GS (all $p$≤.001, all $q$≤.004) and INS (all $p$≤.008, all $q$≤.02).

During REM, we found that MED group displayed less theta and theta beta ratio than the two other groups (all $p$≤.001, all $q$≤.002), as alpha beta ratio in parietal (all $p$≤.01, all $q$≤.04); and frontal regions when compared to GS ($p$=.003, $q$=.01).

#### Table S7 – Chronic sedative-hypnotic use affects SOs and spindles during NREM

| Outcome measure | GS | | INS | | MED | | GS vs INS vs MED<br>Kruskal-Wallis test or ANOVA | | | | | GS vs INS<br>post hoc - Dunn test or t test | | | effect size | INS vs MED<br>post hoc - Dunn test or t test | | | effect size | GS vs MED<br>post hoc - Dunn test or t test | | | effect size |
|---|---|---|---|---|---|---|---|---|---|---|---|---|---|---|---|---|---|---|---|---|---|---|---|
| | Mean | SD | Mean | SD | Mean | SD | F | df | residual | p | q | t | p | q | g' | t | p | q | g' | t | p | q | g' |
| ***Spindle characteristics*** | | | | | | | | | | | | | | | | | | | | | | | |
| **Fz** | | | | | | | | | | | | | | | | | | | | | | | |
| Density (Nber/30 sec) | 1.19 | 0.2 | 1.1 | 0.19 | 1.29 | 0.3 | 8.43 | 2 | 97 | 0.01 | **0.04*** | 1.21 | 0.23 | - | 0.41 | -2.85 | 0.004 | **0.01*** | -0.79 | -1.54 | 0.12 | - | -0.41 |
| Duration (sec) | 0.77 | 0.04 | 0.75 | 0.03 | 0.77 | 0.1 | 2.85 | 2 | 97 | 0.24 | - | - | - | - | - | - | - | - | - | - | - | - | - |
| Amplitude (µV) | 92.7 | 18.3 | 94.92 | 23.3 | 83.14 | 21.6 | 7.83 | 2 | 97 | 0.02 | **0.04*** | -0.02 | 0.98 | - | -0.11 | 2.3 | **0.02** | 0.07 | 0.53 | 2.33 | **0.02** | 0.06 | 0.46 |
| Frequency (Hz) | 11.36 | 0.5 | 11.29 | 0.5 | 11.35 | 0.5 | 0.22 | 2 | 97 | 0.81 | - | - | - | - | - | - | - | - | - | - | - | - | - |
| **Pz** | | | | | | | | | | | | | | | | | | | | | | | |
| Density (Nber/30 sec) | 1.45 | 0.27 | 1.38 | 0.34 | 1.48 | 0.36 | 0.74 | 2 | 97 | 0.48 | - | - | - | - | - | - | - | - | - | - | - | - | - |
| Duration (sec) | 0.74 | 0.04 | 0.73 | 0.04 | 0.75 | 0.06 | 1.09 | 2 | 97 | 0.58 | - | - | - | - | - | - | - | - | - | - | - | - | - |
| Amplitude (µV) | 71.25 | 17.26 | 77.08 | 21.64 | 65.21 | 14.3 | 7.69 | 2 | 97 | **0.02** | 0.08 | -0.76 | 0.45 | - | -0.29 | 2.61 | 0.01 | **0.03*** | 0.68 | 1.82 | 0.07 | - | 0.39 |
| Frequency (Hz) | 13.53 | 0.56 | 13.81 | 0.46 | 13.49 | 0.59 | 2.11 | 2 | 97 | 0.13 | - | - | - | - | - | - | - | - | - | - | - | - | - |
| ***SO characteristics*** | | | | | | | | | | | | | | | | | | | | | | | |
| **Fz** | | | | | | | | | | | | | | | | | | | | | | | |
| Density (Nber/30 sec) | 3.83 | 0.78 | 3.98 | 0.82 | 3.82 | 0.71 | 1.05 | 2 | 97 | 0.59 | - | - | - | - | - | - | - | - | - | - | - | - | - |
| Duration (sec) | 1.35 | 0.06 | 1.37 | 0.05 | 1.41 | 0.08 | 9.63 | 2 | 97 | **0.01*** | - | -0.64 | 0.53 | - | -0.2 | -2.11 | **0.03** | 0.10 | -0.58 | -2.89 | 0.003 | **0.01*** | -0.71 |
| Amplitude (µV) | 122.26 | 30.17 | 124.16 | 37.02 | 104.61 | 28.41 | 9.45 | 2 | 97 | **0.01*** | - | 0.05 | 0.96 | - | -0.06 | 2.48 | 0.01 | **0.04*** | 0.61 | 2.6 | 0.01 | **0.03*** | 0.6 |
| Frequency (Hz) | 0.76 | 0.06 | 0.73 | 0.05 | 0.7 | 0.07 | 16.24 | 2 | 97 | < 0.001 | **0.001*** | 1.5 | 0.13 | - | 0.46 | 2.18 | **0.03** | 0.09 | 0.57 | 3.95 | **< 0.001*** | - | 0.92 |
| **Pz** | | | | | | | | | | | | | | | | | | | | | | | |
| Density (Nber/30 sec) | 3.41 | 0.73 | 3.62 | 0.86 | 3.5 | 0.67 | 1.27 | 2 | 97 | 0.53 | - | - | - | - | - | - | - | - | - | - | - | - | - |
| Duration (sec) | 1.43 | 0.07 | 1.43 | 0.06 | 1.46 | 0.06 | 4.68 | 2 | 97 | 0.09 | - | - | - | - | - | - | - | - | - | - | - | - | - |
| Amplitude (µV) | 96.2 | 26.29 | 102.85 | 32.2 | 84.67 | 21.39 | 7.86 | 2 | 97 | **0.02** | 0.08 | -0.43 | 0.67 | - | -0.22 | 2.51 | 0.01 | **0.04*** | 0.7 | 2.08 | **0.04** | 0.11 | 0.49 |
| Frequency (Hz) | 0.68 | 0.06 | 0.67 | 0.06 | 0.65 | 0.07 | 5.61 | 2 | 97 | 0.06 | - | - | - | - | - | - | - | - | - | - | - | - | - |

SO, slow oscillation

#### Table S8: Chronic sedatives-hypnotic use affects SOs and spindles during N2 and N3

| Outcome measure | GS | | INS | | MED | | GS vs INS vs MED<br>Kruskal-Wallis test or ANOVA | | | | | GS vs INS<br>post hoc - Dunn test or t test | | | effect size | INS vs MED<br>post hoc - Dunn test or t test | | | effect size | GS vs MED<br>post hoc - Dunn test or t test | | | effect size |
|---|---|---|---|---|---|---|---|---|---|---|---|---|---|---|---|---|---|---|---|---|---|---|---|
| | Mean | SD | Mean | SD | Mean | SD | F | df | residual | p | q | t | p | q | g' | t | p | q | g' | t | p | q | g' |
| **N2** | | | | | | | | | | | | | | | | | | | | | | | |
| ***Spindle characteristics*** | | | | | | | | | | | | | | | | | | | | | | | |
| **Fz** | | | | | | | | | | | | | | | | | | | | | | | |
| Density (Nber/30 sec) | 1.17 | 0.24 | 1.04 | 0.24 | 1.28 | 0.24 | 14.67 | 2 | 97 | 0.003 | **0.01*** | 1.73 | 0.08 | - | 0.54 | -3.79 | **< 0.001*** | - | -0.99 | -1.9 | 0.06 | - | -0.45 |
| Duration (sec) | 0.78 | 0.04 | 0.76 | 0.06 | 0.77 | 0.05 | 2.46 | 2 | 97 | 0.39 | - | - | - | - | - | - | - | - | - | - | - | - | - |
| Amplitude (µV) | 90.82 | 17.61 | 95.19 | 33.54 | 82.02 | 21.33 | 6.13 | 2 | 97 | **0.047** | 0.09 | 0.39 | 0.69 | - | -0.16 | 1.77 | 0.07 | - | 0.5 | 2.26 | **0.024** | 0.07 | 0.43 |
| Frequency (Hz) | 11.49 | 0.53 | 11.51 | 0.49 | 11.41 | 0.46 | 0.17 | 2 | 97 | 0.84 | - | - | - | - | - | - | - | - | - | - | - | - | - |
| Cz | | | | | | | | | | | | | | | | | | | | | | | |
| Density (Nber/30 sec) | 1.27 | 0.25 | 1.19 | 0.27 | 1.33 | 0.3 | 5.19 | 2 | 97 | 0.15 | - | - | - | - | - | - | - | - | - | - | - | - | - |
| Duration (sec) | 0.74 | 0.04 | 0.74 | 0.04 | 0.75 | 0.04 | 0.56 | 2 | 97 | 0.76 | - | - | - | - | - | - | - | - | - | - | - | - | - |
| Amplitude (µV) | 82.83 | 17.31 | 108.41 | 67.97 | 78.98 | 34.58 | 9.86 | 2 | 97 | **0.03** | 0.12 | -1.05 | 0.29 | - | -0.52 | 3.02 | 0.003 | **0.008*** | 0.59 | 1.9 | 0.06 | - | 0.13 |
| Frequency (Hz) | 13.28 | 0.49 | 13.38 | 0.57 | 13.23 | 0.49 | 0.28 | 2 | 97 | 0.76 | - | - | - | - | - | - | - | - | - | - | - | - | - |
| Pz | | | | | | | | | | | | | | | | | | | | | | | |
| Density (Nber/30 sec) | 1.43 | 0.29 | 1.36 | 0.33 | 1.49 | 0.36 | 1.28 | 2 | 97 | 0.28 | - | - | - | - | - | - | - | - | - | - | - | - | - |
| Duration (sec) | 0.75 | 0.05 | 0.75 | 0.05 | 0.76 | 0.06 | 0.19 | 2 | 97 | 0.91 | - | - | - | - | - | - | - | - | - | - | - | - | - |
| Amplitude (µV) | 69.34 | 16.3 | 76.11 | 23.04 | 64.44 | 14.59 | 6.59 | 2 | 97 | **0.037** | 0.07 | -0.67 | 0.50 | - | -0.34 | 2.41 | 0.016 | **0.048*** | 0.64 | 1.7 | 0.08 | - | 0.32 |
| Frequency (Hz) | 13.59 | 0.57 | 13.95 | 0.47 | 13.52 | 0.58 | 3.6 | 2 | 97 | **0.031** | 0.07 | -2.354 | **0.021** | 0.06 | -0.33 | 0.533 | 0.002 | **0.007*** | -0.33 | 0.53 | 0.59 | - | 0.12 |
| ***SO characteristics*** | | | | | | | | | | | | | | | | | | | | | | | |
| Fz | | | | | | | | | | | | | | | | | | | | | | | |
| Density (Nber/30 sec) | 3.55 | 0.83 | 3.64 | 0.86 | 3.76 | 0.72 | 1.01 | 2 | 97 | 0.38 | - | - | - | - | - | - | - | - | - | - | - | - | - |
| Duration (sec) | 1.35 | 0.06 | 1.34 | 0.06 | 1.4 | 0.08 | 12.70 | 2 | 97 | 0.004 | **0.008*** | 0.05 | 0.96 | - | 0.06 | -2.94 | 0.003 | **0.01*** | -0.8 | -2.95 | 0.003 | **0.01*** | -0.75 |
| Amplitude (µV) | 110.01 | 26.19 | 106.26 | 29.19 | 100.09 | 27.22 | 3.22 | 2 | 97 | 0.26 | - | - | - | - | - | - | - | - | - | - | - | - | - |
| Frequency (Hz) | 0.76 | 0.06 | 0.78 | 0.06 | 0.7 | 0.08 | 21.24 | 2 | 97 | < 0.001 | **0.004*** | -0.86 | 0.38 | - | -0.31 | 4.19 | **< 0.001*** | - | 1.06 | 3.31 | 0.001 | **0.003*** | 0.79 |
| Cz | | | | | | | | | | | | | | | | | | | | | | | |
| Density (Nber/30 sec) | 3.37 | 0.77 | 3.28 | 0.97 | 3.62 | 0.67 | 4.80 | 2 | 97 | 0.09 | - | - | - | - | - | - | - | - | - | - | - | - | - |
| Duration (sec) | 1.37 | 0.07 | 1.37 | 0.06 | 1.42 | 0.07 | 10.3 | 2 | 97 | 0.015 | **0.032*** | 0.24 | 0.80 | - | 0.03 | -2.75 | 0.006 | **0.02*** | -0.7 | -2.54 | 0.01 | **0.03*** | -0.64 |
| Amplitude (µV) | 103.36 | 22.91 | 115.63 | 46.61 | 93.91 | 21.04 | 6.20 | 2 | 97 | **0.045** | 0.06 | -0.5 | 0.61 | - | -0.33 | 2.28 | **0.02** | 0.07 | 0.66 | 1.77 | 0.07 | - | 0.43 |
| Frequency (Hz) | 0.72 | 0.08 | 0.72 | 0.06 | 0.68 | 0.06 | 9.70 | 2 | 97 | 0.016 | **0.032*** | 0.39 | 0.69 | - | 0.14 | 2.3 | **0.02** | 0.07 | 0.61 | 2.8 | 0.005 | **0.02*** | 0.7 |
| Pz | | | | | | | | | | | | | | | | | | | | | | | |
| Density (Nber/30 sec) | 3.17 | 0.76 | 3.3 | 0.89 | 3.47 | 0.69 | 3.42 | 2 | 97 | 0.24 | - | - | - | - | - | - | - | - | - | - | - | - | - |
| Duration (sec) | 1.41 | 0.06 | 1.4 | 0.05 | 1.44 | 0.07 | 11.39 | 2 | 97 | 0.007 | **0.014*** | 0.5 | 0.61 | - | 0.13 | -3.01 | 0.003 | **0.01*** | -0.74 | -2.52 | 0.01 | **0.036*** | -0.58 |
| Amplitude (µV) | 85.76 | 21.26 | 89.67 | 29.33 | 81.19 | 20.89 | 2.01 | 2 | 97 | 0.37 | - | - | - | - | - | - | - | - | - | - | - | - | - |
| Frequency (Hz) | 0.7 | 0.06 | 0.71 | 0.06 | 0.65 | 0.07 | 14.39 | 2 | 97 | 0.003 | **0.012*** | -0.48 | 0.62 | - | -0.18 | 3.35 | 0.001 | **0.002*** | 0.84 | 2.88 | 0.004 | **0.01*** | 0.68 |
| **N3** | | | | | | | | | | | | | | | | | | | | | | | |
| ***Spindle characteristics*** | | | | | | | | | | | | | | | | | | | | | | | |
| **Fz** | | | | | | | | | | | | | | | | | | | | | | | |
| Density (Nber/30 sec) | 1.2 | 0.23 | 1.21 | 0.2 | 1.29 | 0.31 | 1.73 | 2 | 97 | 0.41 | - | - | - | - | - | - | - | - | - | - | - | - | - |
| Duration (sec) | 0.75 | 0.04 | 0.74 | 0.04 | 0.75 | 0.07 | 1.71 | 2 | 97 | 0.42 | - | - | - | - | - | - | - | - | - | - | - | - | - |
| Amplitude (µV) | 97.5 | 20.24 | 98.36 | 24.63 | 91.73 | 24.14 | 1.94 | 2 | 97 | 0.37 | - | - | - | - | - | - | - | - | - | - | - | - | - |
| Frequency (Hz) | 11.1 | 0.49 | 11.03 | 0.56 | 10.93 | 0.45 | 2.30 | 2 | 97 | 0.31 | - | - | - | - | - | - | - | - | - | - | - | - | - |
| Cz | | | | | | | | | | | | | | | | | | | | | | | |
| Density (Nber/30 sec) | 1.2 | 0.3 | 1.16 | 0.43 | 0.17 | 0.32 | 0.57 | 2 | 97 | 0.74 | - | - | - | - | - | - | - | - | - | - | - | - | - |
| Duration (sec) | 0.7 | 0.04 | 0.7 | 0.03 | 0.71 | 0.06 | 0.35 | 2 | 97 | 0.83 | - | - | - | - | - | - | - | - | - | - | - | - | - |
| Amplitude (µV) | 90.34 | 19.18 | 110.87 | 84.49 | 83.74 | 18.82 | 4.71 | 2 | 97 | 0.09 | - | - | - | - | - | - | - | - | - | - | - | - | - |
| Frequency (Hz) | 13 | 0.51 | 13.1 | 0.71 | 12.72 | 0.64 | 8.29 | 2 | 97 | **0.016** | 0.06 | -0.6 | 0.55 | - | -0.16 | 2.65 | 0.008 | **0.024*** | 0.56 | 2.04 | **0.041** | 0.12 | 0.47 |
| Pz | | | | | | | | | | | | | | | | | | | | | | | |
| Density (Nber/30 sec) | 1.45 | 0.27 | 1.38 | 0.34 | 1.48 | 0.36 | 1.16 | 2 | 97 | 0.55 | - | - | - | - | - | - | - | - | - | - | - | - | - |
| Duration (sec) | 0.74 | 0.04 | 0.73 | 0.04 | 0.75 | 0.06 | 0.31 | 2 | 97 | 0.85 | - | - | - | - | - | - | - | - | - | - | - | - | - |
| Amplitude (µV) | 71.25 | 17.26 | 77.08 | 21.64 | 65.21 | 14.3 | 5.09 | 2 | 97 | 0.08 | - | - | - | - | - | - | - | - | - | - | - | - | - |
| Frequency (Hz) | 13.53 | 0.56 | 13.81 | 0.46 | 13.49 | 0.59 | 8.52 | 2 | 97 | **0.01** | 0.056 | -1.63 | 0.10 | - | -0.46 | 2.92 | 0.004 | **0.01*** | 0.7 | 1.14 | 0.25 | - | 0.29 |
| ***SO characteristics*** | | | | | | | | | | | | | | | | | | | | | | | |
| Fz | | | | | | | | | | | | | | | | | | | | | | | |
| Density (Nber/30 sec) | 4.38 | 0.83 | 4.52 | 0.73 | 4.28 | 0.77 | 1.58 | 2 | 97 | 0.45 | - | - | - | - | - | - | - | - | - | - | - | - | - |
| Duration (sec) | 1.39 | 0.08 | 1.39 | 0.06 | 1.43 | 0.10 | 4.20 | 2 | 97 | 0.12 | - | - | - | - | - | - | - | - | - | - | - | - | - |
| Amplitude (µV) | 143.76 | 36.48 | 144.55 | 43.56 | 133.52 | 38.37 | 2.55 | 2 | 97 | 0.27 | - | - | - | - | - | - | - | - | - | - | - | - | - |
| Frequency (Hz) | 0.73 | 0.09 | 0.7 | 0.05 | 0.67 | 0.07 | 11.78 | 2 | 97 | 0.01 | **0.04*** | 0.94 | 0.34 | - | 0.34 | 2.16 | **0.03** | 0.09 | 0.59 | 3.28 | 0.001 | **0.003*** | 0.82 |
| Cz | | | | | | | | | | | | | | | | | | | | | | | |
| Density (Nber/30 sec) | 4.12 | 0.82 | 4.15 | 0.73 | 4.00 | 0.72 | 0.70 | 2 | 97 | 0.70 | - | - | - | - | - | - | - | - | - | - | - | - | - |
| Duration (sec) | 1.44 | 0.08 | 1.44 | 0.06 | 1.47 | 0.08 | 3.27 | 2 | 97 | 0.19 | - | - | - | - | - | - | - | - | - | - | - | - | - |
| Amplitude (µV) | 132.65 | 32.97 | 143.2 | 46.44 | 122.97 | 28.05 | 3.57 | 2 | 97 | 0.16 | - | - | - | - | - | - | - | - | - | - | - | - | - |
| Frequency (Hz) | 0.67 | 0.06 | 0.66 | 0.06 | 0.63 | 0.063 | 9.7 | 2 | 97 | **0.03** | 0.12 | 1.04 | 0.30 | - | 0.29 | 1.82 | 0.07 | - | 0.43 | 3.03 | 0.002 | **0.007*** | 0.71 |
| Pz | | | | | | | | | | | | | | | | | | | | | | | |
| Density (Nber/30 sec) | 3.89 | 0.79 | 4.1 | 0.79 | 3.80 | 0.75 | 2.38 | 2 | 97 | 0.30 | - | - | - | - | - | - | - | - | - | - | - | - | - |
| Duration (sec) | 1.47 | 0.08 | 1.47 | 0.07 | 1.50 | 0.07 | 5.23 | 2 | 97 | 0.07 | - | - | - | - | - | - | - | - | - | - | - | - | - |
| Amplitude (µV) | 113.78 | 32.12 | 118.89 | 36.44 | 106.90 | 24.80 | 0.21 | 2 | 97 | 0.81 | - | - | - | - | - | - | - | - | - | - | - | - | - |
| Frequency (Hz) | 0.63 | 0.06 | 0.63 | 0.06 | 0.61 | 0.05 | 3.34 | 2 | 97 | 0.19 | - | - | - | - | - | - | - | - | - | - | - | - | - |

SO, slow oscillation

#### Table S9 – Effect of chronic sedative-hypnotic use on spindles using Moelle, Ray and Lacourse detection algorithms during NREM

| Outcome measure | GS | | INS | | MED | | GS vs INS vs MED Kruskal-Wallis test or ANOVA | | | | | GS vs INS post hoc - Dunn test or t test | | | effect size | INS vs MED post hoc - Dunn test or t test | | | effect size | GS vs MED post hoc - Dunn test or t test | | | effect size |
|---|---|---|---|---|---|---|---|---|---|---|---|---|---|---|---|---|---|---|---|---|---|---|---|
| | Mean | SD | Mean | SD | Mean | SD | F | df | residual | p | q | t | p | q | g' | t | p | q | g' | t | p | q | g' |
| **MOELLE** | | | | | | | | | | | | | | | | | | | | | | | |
| ***Spindle characteristics*** | | | | | | | | | | | | | | | | | | | | | | | |
| **Fz** | | | | | | | | | | | | | | | | | | | | | | | |
| Count | 678.54 | 149.6 | 563,0 | 145.9 | 688.53 | 152.7 | 7.31 | 2 | 97 | 0.03 | **0.05*** | 2.1 | **0.04** | 0.11 | 0.77 | -2.6 | 0.01 | **0.03*** | -0.83 | -0.26 | 0.79 | - | -0.07 |
| Density | 1.19 | 0.2 | 1.1 | 0.19 | 1.29 | 0.3 | 8.43 | 2 | 97 | 0.01 | **0.05*** | 1.21 | 0.23 | - | 0.41 | -2.85 | 0.004 | **0.01*** | -0.79 | -1.54 | 0.12 | - | -0.41 |
| Duration | 0.77 | 0.04 | 0.75 | 0.03 | 0.77 | 0.1 | 2.85 | 2 | 97 | 0.24 | - | 1.63 | 0.10 | - | 0.49 | -1.31 | 0.19 | - | -0.39 | 0.52 | 0.60 | - | -0.02 |
| Amplitude | 92.7 | 18.3 | 94.92 | 23.3 | 83.14 | 21.6 | 7.83 | 2 | 97 | 0.02 | **0.05*** | -0.02 | 0.98 | - | -0.11 | 2.3 | **0.02** | 0.07 | 0.53 | 2.33 | **0.02** | 0.06 | 0.46 |
| Frequency | 11.36 | 0.5 | 11.29 | 0.5 | 11.35 | 0.5 | 0.22 | 2 | 97 | 0.81 | - | 0.59 | 0.56 | - | 0.15 | 0.072 | 0.56 | - | -0.14 | 0.072 | 0.94 | - | 0.02 |
| **Cz** | | | | | | | | | | | | | | | | | | | | | | | |
| Count | 721.21 | 177.89 | 598.77 | 178.97 | 700.87 | 164.83 | 4.33 | 2 | 97 | 0.11 | - | 1.71 | 0.08 | - | 0.68 | -1.96 | **0.05** | 0.15 | -0.59 | -0.05 | 0.96 | - | 0.12 |
| Density | 1.26 | 0.24 | 1.18 | 0.31 | 1.31 | 0.3 | 5.07 | 2 | 97 | 0.08 | - | 1 | 0.31 | - | 0.29 | -2.22 | **0.03** | 0.08 | -0.44 | -1.14 | 0.25 | - | -0.19 |
| Duration | 0.73 | 0.04 | 0.73 | 0.03 | 0.74 | 0.04 | 3.78 | 2 | 97 | 0.15 | - | 0.36 | 0.71 | - | 0.08 | -1.77 | 0.08 | - | -0.43 | -1.4 | 0.16 | - | -0.33 |
| Amplitude | 84.78 | 17.9 | 108.62 | 71.08 | 79.81 | 33.72 | 11.50 | 2 | 97 | 0.003 | **0.016*** | -1.09 | 0.27 | - | -0.46 | 3.25 | 0.001 | **0.003*** | 0.57 | 2.09 | **0.036** | 0.11 | 0.17 |
| Frequency | 13.21 | 0.43 | 13.27 | 0.57 | 13.18 | 0.5 | 1.87 | 2 | 97 | 0.39 | - | -0.95 | 0.34 | - | -0.12 | 1.35 | 0.17 | - | 0.17 | 0.3 | 0.76 | - | 0.06 |
| **Pz** | | | | | | | | | | | | | | | | | | | | | | | |
| Count | 826.68 | 177.15 | 703.12 | 207.86 | 796.6 | 222.7 | 4.7 | 2 | 97 | 0.10 | - | 2.14 | **0.03** | 0.10 | 0.63 | -1.53 | 0.13 | - | -0.43 | 0.87 | 0.38 | - | 0.14 |
| Density | 1.45 | 0.27 | 1.38 | 0.34 | 1.48 | 0.36 | 0.74 | 2 | 97 | 0.48 | - | 0.75 | 0.46 | - | 0.22 | -0.429 | 0.21 | - | -0.29 | -0.429 | 0.67 | - | -0.1 |
| Duration | 0.74 | 0.04 | 0.73 | 0.04 | 0.75 | 0.06 | 1.09 | 2 | 97 | 0.58 | - | 0.45 | 0.65 | - | 0.09 | -1.03 | 0.30 | - | -0.34 | -0.53 | 0.59 | - | -0.26 |
| Amplitude | 71.25 | 17.26 | 77.08 | 21.64 | 65.21 | 14.3 | 7.69 | 2 | 97 | **0.02** | 0.08 | -0.76 | 0.45 | - | -0.29 | 2.61 | 0.01 | **0.03*** | 0.68 | 1.82 | 0.07 | - | 0.39 |
| Frequency | 13.53 | 0.56 | 13.81 | 0.46 | 13.49 | 0.59 | 2.11 | 2 | 97 | 0.13 | - | -1.91 | 0.06 | - | -0.55 | 0.28 | 0.02 | **0.05*** | 0.58 | 0.28 | 0.78 | - | 0.06 |
| **RAY** | | | | | | | | | | | | | | | | | | | | | | | |
| ***Spindle characteristics*** | | | | | | | | | | | | | | | | | | | | | | | |
| **Fz** | | | | | | | | | | | | | | | | | | | | | | | |
| Count | 743.29 | 275.05 | 608.42 | 187.16 | 789.79 | 294.37 | 7.36 | 2 | 97 | 0.025 | **0.044*** | 1.77 | 0.07 | - | 0.56 | -2.7 | 0.007 | **0.02*** | -0.69 | -0.74 | 0.45 | - | -0.16 |
| Density | 1.29 | 0.44 | 1.2 | 0.31 | 1.48 | 0.5 | 5.53 | 2 | 97 | 0.06 | - | 0.87 | 0.38 | - | 0.25 | -2.29 | **0.02** | 0.07 | -0.62 | -1.35 | 0.17 | - | -0.38 |
| Duration | 0.73 | 0.05 | 0.71 | 0.04 | 0.75 | 0.07 | 7.61 | 2 | 97 | 0.02 | **0.044*** | 1.98 | **0.047** | 0.14 | 0.51 | -2.71 | 0.007 | **0.02*** | -0.61 | -0.52 | 0.60 | - | -0.24 |
| Amplitude | 89.35 | 18.03 | 90.76 | 22.43 | 79.37 | 21.11 | 8.06 | 2 | 97 | 0.02 | **0.044*** | 0.04 | 0.97 | - | -0.07 | 2.3 | **0.02** | 0.06 | 0.52 | 2.39 | 0.02 | **0.05*** | 0.49 |
| Frequency | 11.32 | 0.55 | 11.4 | 0.54 | 11.45 | 0.5 | 0.83 | 2 | 97 | 0.65 | - | -0.35 | 0.72 | - | -0.14 | -0.49 | 0.62 | - | -0.1 | -0.9 | 0.36 | - | -0.24 |
| **Cz** | | | | | | | | | | | | | | | | | | | | | | | |
| Count | 703.86 | 283.04 | 552.08 | 224.29 | 733.81 | 238.33 | 8.21 | 2 | 97 | 0.016 | **0.02*** | 1.45 | 0.14 | - | 0.58 | -2.86 | 0.004 | **0.01*** | -0.77 | -1.27 | 0.20 | - | -0.12 |
| Density | 1.22 | 0.42 | 1.08 | 0.44 | 1.38 | 0.43 | 9.05 | 2 | 97 | 0.01 | **0.017*** | 1.17 | 0.24 | - | 0.32 | -2.94 | 0.003 | **0.01*** | -0.68 | -1.67 | 0.09 | - | -0.36 |
| Duration | 0.71 | 0.05 | 0.69 | 0.04 | 0.73 | 0.06 | 9.83 | 2 | 97 | 0.007 | **0.017*** | 1.23 | 0.22 | - | 0.37 | -3.06 | 0.002 | **0.006*** | -0.72 | -1.74 | 0.08 | - | -0.36 |
| Amplitude | 84.51 | 17.61 | 101.79 | 45.43 | 76.44 | 15.35 | 11.67 | 2 | 97 | 0.003 | **0.015*** | -1.06 | 0.28 | - | -0.5 | 3.26 | 0.001 | **0.003*** | 0.85 | 2.13 | **0.03** | 0.09 | 0.49 |
| Frequency | 12.24 | 0.76 | 12.24 | 0.9 | 12.25 | 0.73 | 0.01 | 2 | 97 | 0.99 | - | 0.02 | 0.98 | - | 0 | 0.08 | 0.93 | - | -0.01 | 0.1 | 0.92 | - | -0.01 |
| **Pz** | | | | | | | | | | | | | | | | | | | | | | | |
| Count | 805.93 | 292.41 | 678.85 | 266.21 | 841.51 | 298.53 | 4.19 | 2 | 97 | 0.12 | - | 1.17 | 0.24 | - | 0.45 | -2.05 | **0.04** | 0.12 | -0.56 | -0.76 | 0.44 | - | -0.12 |
| Density | 1.41 | 0.48 | 1.32 | 0.48 | 1.58 | 0.52 | 5.27 | 2 | 97 | 0.07 | - | 0.69 | 0.49 | - | 0.19 | -2.19 | **0.03** | 0.08 | -0.5 | -1.45 | 0.14 | - | -0.32 |
| Duration | 0.73 | 0.06 | 0.72 | 0.05 | 0.76 | 0.09 | 4.33 | 2 | 97 | 0.11 | - | 0.44 | 0.66 | - | 0.16 | -1.91 | 0.056 | - | -0.5 | -1.46 | 0.14 | - | -0.37 |
| Amplitude | 71.07 | 17.48 | 76.06 | 20.01 | 64.81 | 12.88 | 7.19 | 2 | 97 | **0.027** | 0.11 | -0.68 | 0.49 | - | -0.26 | 2.51 | 0.01 | **0.035*** | 0.71 | 1.79 | 0.07 | - | 0.42 |
| Frequency | 12.91 | 0.66 | 13.04 | 0.84 | 12.9 | 0.87 | 2.60 | 2 | 97 | 0.27 | - | -1.59 | 0.11 | - | -0.17 | 1.14 | 0.25 | - | 0.16 | -0.65 | 0.51 | - | 0.02 |
| **LACOURSE** | | | | | | | | | | | | | | | | | | | | | | | |
| ***Spindle characteristics*** | | | | | | | | | | | | | | | | | | | | | | | |
| **Fz** | | | | | | | | | | | | | | | | | | | | | | | |
| Count | 426.07 | 242.93 | 393.08 | 233.96 | 636.62 | 382.33 | 9.98 | 2 | 97 | 0.007 | **0.02*** | 0.4 | 0.69 | - | 0.14 | -2.79 | 0.005 | **0.015*** | -0.71 | -2.4 | 0.016 | **0.048*** | -0.62 |
| Density | 0.74 | 0.41 | 0.77 | 0.45 | 1.22 | 0.75 | 10.38 | 2 | 97 | 0.006 | **0.02*** | -0.24 | 0.80 | - | -0.07 | -2.48 | 0.013 | **0.039*** | -0.67 | -2.82 | 0.005 | **0.01*** | -0.73 |
| Duration | 0.91 | 0.08 | 0.91 | 0.08 | 0.93 | 0.09 | 1.67 | 2 | 97 | 0.43 | - | 0.04 | 0.96 | - | 0 | -1.08 | 0.28 | - | -0.21 | -1.06 | 0.29 | - | -0.21 |
| Amplitude | 78.65 | 12.18 | 78.44 | 14.38 | 71.98 | 15.99 | 7.16 | 2 | 97 | 0.023 | **0.045*** | 0.09 | 0.93 | - | 0.02 | 2.13 | **0.03** | 0.09 | 0.41 | 2.28 | **0.02** | 0.07 | 0.45 |
| Frequency | 11.74 | 0.5 | 11.94 | 0.55 | 11.85 | 0.4 | 1.48 | 2 | 97 | 0.47 | - | -1.2 | 0.22 | - | -0.38 | 0.55 | 0.58 | - | 0.2 | -0.81 | 0.41 | - | -0.25 |
| **Cz** | | | | | | | | | | | | | | | | | | | | | | | |
| Count | 588.71 | 334.67 | 487.88 | 289.32 | 720.17 | 357.26 | 9.74 | 2 | 97 | 0.008 | **0.013*** | 0.68 | 0.49 | - | 0.32 | -2.88 | 0.004 | **0.01*** | -0.69 | -2.17 | **0.03** | 0.08 | -0.37 |
| Density | 1.02 | 0.53 | 0.95 | 0.57 | 1.37 | 0.69 | 9.69 | 2 | 97 | 0.008 | **0.013*** | 0.24 | 0.81 | - | 0.12 | -2.67 | 0.01 | **0.02*** | -0.64 | -2.46 | 0.01 | **0.04*** | -0.54 |
| Duration | 0.91 | 0.07 | 0.9 | 0.07 | 0.92 | 0.08 | 1.71 | 2 | 97 | 0.42 | - | -0.09 | 0.93 | - | 0.03 | -1.02 | 0.30 | - | -0.26 | -1.14 | 0.25 | - | -0.23 |
| Amplitude | 77.17 | 14.34 | 86.89 | 25.4 | 71.65 | 12.96 | 9.80 | 2 | 97 | 0.007 | **0.016*** | -1.24 | 0.21 | - | -0.47 | 3.06 | 0.002 | **0.006*** | 0.82 | 1.72 | 0.08 | - | 0.41 |
| Frequency | 12.7 | 0.56 | 12.76 | 0.72 | 12.63 | 0.55 | 1.32 | 2 | 97 | 0.51 | - | -0.3 | 0.76 | - | -0.08 | 1.08 | 0.27 | - | 0.2 | 0.76 | 0.44 | - | 0.13 |
| **Pz** | | | | | | | | | | | | | | | | | | | | | | | |
| Count | 736.86 | 425.57 | 626.42 | 392.33 | 777.94 | 447.74 | 1.86 | 2 | 97 | 0.39 | - | 0.68 | 0.49 | - | 0.27 | -1.36 | 0.17 | - | -0.35 | -0.62 | 0.53 | - | -0.09 |
| Density | 1.29 | 0.72 | 1.22 | 0.75 | 1.47 | 0.84 | 2.16 | 2 | 97 | 0.33 | - | 0.31 | 0.75 | - | 0.09 | -1.35 | 0.17 | - | -0.31 | -1.03 | 0.30 | - | -0.23 |
| Duration | 0.88 | 0.07 | 0.89 | 0.07 | 0.9 | 0.1 | 1.29 | 2 | 97 | 0.52 | - | -0.68 | 0.49 | - | -0.16 | -0.35 | 0.72 | - | -0.17 | -1.14 | 0.25 | - | -0.29 |
| Amplitude | 67.77 | 14.9 | 70.98 | 15.9 | 62.65 | 10.34 | 6.84 | 2 | 97 | **0.03** | 0.13 | -0.71 | 0.48 | - | -0.21 | 2.46 | 0.01 | **0.041*** | 0.65 | 1.72 | 0.08 | - | 0.41 |
| Frequency | 13.23 | 0.55 | 13.42 | 0.64 | 13.16 | 0.68 | 5.46 | 2 | 97 | 0.06 | - | -1.98 | **0.047** | 0.14 | -0.32 | 2.16 | **0.03** | 0.09 | 0.39 | -0.05 | 0.96 | - | 0.11 |

#### Table S10 – Effect of chronic sedative-hypnotic use on SOs using Staresina and Massimini detection algorithms during NREM

| Outcome measure | GS | | INS | | MED | | GS vs INS vs MED Kruskal-Wallis test or ANOVA | | | | | GS vs INS post hoc - Dunn test or t test | | | effect size | INS vs MED post hoc - Dunn test or t test | | | effect size | GS vs MED post hoc - Dunn test or t test | | | effect size |
|---|---|---|---|---|---|---|---|---|---|---|---|---|---|---|---|---|---|---|---|---|---|---|---|
| | Mean | SD | Mean | SD | Mean | SD | F | df | residual | p | q | t | p | q | g' | t | p | q | g' | t | p | q | g' |
| **STARESINA** | | | | | | | | | | | | | | | | | | | | | | | |
| ***SO characteristics*** | | | | | | | | | | | | | | | | | | | | | | | |
| **Fz** | | | | | | | | | | | | | | | | | | | | | | | |
| Count | 2181.89 | 455.25 | 2029.5 | 571.38 | 2061 | 513.33 | 0.76 | 2 | 97 | 0.47 | - | 1.09 | 0.28 | - | 0.29 | 0.99 | 0.80 | | -0.06 | 0.98 | 0.33 | | 0.24 |
| Density | 3.83 | 0.78 | 3.98 | 0.82 | 3.82 | 0.71 | 1.05 | 2 | 97 | 0.59 | - | -0.85 | 0.39 | - | -0.19 | 0.95 | 0.34 | | 0.21 | 0 | 0.99 | | 0.01 |
| Duration | 1.35 | 0.06 | 1.37 | 0.05 | 1.41 | 0.08 | 9.63 | 2 | 97 | **0.01*** | - | -0.64 | 0.53 | - | -0.2 | -2.11 | **0.03** | 0.10 | -0.58 | -2.89 | 0.003 | **0.01*** | -0.71 |
| Amplitude | 122.26 | 30.17 | 124.16 | 37.02 | 104.61 | 28.41 | 9.45 | 2 | 97 | **0.01*** | - | 0.05 | 0.96 | - | -0.06 | 2.48 | 0.01 | **0.04*** | 0.61 | 2.6 | 0.01 | **0.03*** | 0.6 |
| Frequency | 0.76 | 0.06 | 0.73 | 0.05 | 0.7 | 0.07 | 16.24 | 2 | 97 | < 0.001 | **0.001*** | 1.5 | 0.13 | - | 0.46 | 2.18 | **0.03** | 0.09 | 0.57 | 3.95 | **< 0.001*** | | 0.92 |
| **Cz** | | | | | | | | | | | | | | | | | | | | | | | |
| Count | 2069.21 | 463.22 | 1853.04 | 595.28 | 1978.89 | 490.25 | 2.9 | 2 | 97 | 0.23 | - | 1.7 | 0.08 | - | 0.4 | -1.05 | 0.29 | - | -0.24 | 0.86 | 0.38 | - | 0.19 |
| Density | 3.61 | 0.73 | 3.62 | 0.84 | 3.67 | 0.65 | 0.21 | 2 | 97 | 0.90 | - | -0.03 | 0.97 | - | -0.01 | -0.36 | 0.72 | - | -0.06 | -0.4 | 0.69 | - | -0.08 |
| Duration | 1.4 | 0.06 | 1.4 | 0.06 | 1.43 | 0.07 | 3.81 | 2 | 97 | 0.15 | - | 0.02 | 0.98 | - | 0 | -1.6 | 0.11 | - | -0.38 | -1.62 | 0.10 | - | -0.38 |
| Amplitude | 113.86 | 27.74 | 128.27 | 46.86 | 97.87 | 21.82 | 13.02 | 2 | 97 | 0.001 | **0.006*** | -0.93 | 0.35 | - | -0.37 | 3.38 | 0.0007 | **0.002*** | 0.91 | 2.4 | 0.016 | **0.049*** | 0.65 |
| Frequency | 0.71 | 0.06 | 0.7 | 0.05 | 0.67 | 0.06 | 5.92 | 2 | 97 | 0.05 | - | 0.63 | 0.53 | - | 0.19 | 1.56 | 0.12 | - | 0.39 | 2.31 | **0.02** | 0.06 | 0.54 |
| **Pz** | | | | | | | | | | | | | | | | | | | | | | | |
| Count | 1954.36 | 446.47 | 1840.65 | 566.78 | 1891.49 | 487.88 | 0.63 | 2 | 97 | 0.54 | - | 0.84 | 0.40 | - | 0.22 | 0.53 | 0.68 | - | -0.1 | 0.53 | 0.60 | - | 0.13 |
| Density | 3.41 | 0.73 | 3.62 | 0.86 | 3.5 | 0.67 | 1.27 | 2 | 97 | 0.53 | - | -1.12 | 0.26 | - | -0.26 | 0.59 | 0.55 | - | 0.16 | -0.67 | 0.50 | - | -0.13 |
| Duration | 1.43 | 0.07 | 1.43 | 0.06 | 1.46 | 0.06 | 4.68 | 2 | 97 | 0.09 | - | 0.21 | 0.84 | - | 0.08 | -1.88 | 0.06 | - | -0.46 | -1.69 | 0.09 | - | -0.36 |
| Amplitude | 96.2 | 26.29 | 102.85 | 32.2 | 84.67 | 21.39 | 7.86 | 2 | 97 | **0.02** | 0.08 | -0.43 | 0.67 | - | -0.22 | 2.51 | 0.01 | **0.04*** | 0.7 | 2.08 | **0.04** | 0.11 | 0.49 |
| Frequency | 0.68 | 0.06 | 0.67 | 0.06 | 0.65 | 0.07 | 5.61 | 2 | 97 | 0.06 | - | 0.28 | 0.78 | - | 0.09 | 1.76 | 0.08 | - | 0.41 | 2.12 | **0.03** | 0.10 | 0.49 |
| **MASSIMINI** | | | | | | | | | | | | | | | | | | | | | | | |
| ***SO characteristics*** | | | | | | | | | | | | | | | | | | | | | | | |
| **Fz** | | | | | | | | | | | | | | | | | | | | | | | |
| Count | 341.14 | 306.52 | 316.15 | 342.08 | 170.79 | 220.54 | 8.79 | 2 | 97 | 0.012 | **0.02*** | 0.55 | 0.57 | - | 0.08 | 2.06 | **0.039** | 0.12 | 0.53 | 2.74 | 0.006 | **0.02*** | 0.66 |
| Density | 0.58 | 0.52 | 0.66 | 0.79 | 0.29 | 0.38 | 10.15 | 2 | 97 | 0.006 | **0.015*** | 0.33 | 0.73 | - | -0.12 | 2.4 | 0.017 | **0.049*** | 0.65 | 2.83 | 0.005 | **0.01*** | 0.65 |
| Duration | 1.19 | 0.13 | 1.176 | 0.11 | 1.3 | 0.19 | 11.21 | 2 | 97 | 0.004 | **0.015*** | -0.04 | 0.96 | - | 0.08 | -2.69 | 0.007 | **0.02*** | -0.7 | -2.83 | 0.005 | **0.01*** | -0.63 |
| Amplitude | 209.48 | 20.18 | 217 | 32.16 | 204.44 | 21.83 | 4.74 | 2 | 97 | 0.09 | - | -0.11 | 0.91 | - | -0.28 | 1.83 | 0.067 | - | 0.48 | 1.76 | 0.08 | - | 0.23 |
| Frequency | 0.75 | 0.09 | 0.76 | 0.08 | 0.71 | 0.11 | 5.30 | 2 | 97 | 0.07 | - | -0.2 | 0.84 | - | -0.07 | 1.97 | **0.048** | 0.14 | 0.48 | 1.82 | 0.07 | - | 0.4 |
| **Cz** | | | | | | | | | | | | | | | | | | | | | | | |
| Count | 156.5 | 210.09 | 153.65 | 215.45 | 59.6 | 86.48 | 8.45 | 2 | 97 | 0.01 | **0.025*** | -0.59 | 0.55 | - | 0.01 | 2.66 | 0.007 | **0.02*** | 0.64 | 2.06 | **0.04** | 0.11 | 0.66 |
| Density | 0.27 | 0.37 | 0.33 | 0.52 | 0.09 | 0.14 | 10.34 | 2 | 97 | 0.006 | **0.025*** | -0.57 | 0.56 | - | -0.13 | 2.91 | 0.004 | **0.01*** | 0.72 | 2.33 | **0.02** | 0.06 | 0.7 |
| Duration | 1.21 | 0.17 | 1.18 | 0.03 | 1.29 | 0.026 | 8.15 | 2 | 97 | 0.017 | **0.028*** | -0.02 | 0.98 | - | 0.18 | -2.34 | **0.02** | 0.06 | -0.67 | -2.4 | 0.02 | **0.049*** | -0.46 |
| Amplitude | 201.06 | 15.47 | 218.2 | 50.24 | 205.75 | 75.86 | 5.08 | 2 | 97 | 0.08 | - | -0.02 | 0.98 | - | -0.46 | 1.87 | 0.06 | - | 0.18 | 1.87 | 0.06 | - | -0.08 |
| Frequency | 0.72 | 0.10 | 0.72 | 0.10 | 0.68 | 0.12 | 4.50 | 2 | 97 | 0.10 | - | 0.3 | 0.76 | - | -0.01 | 1.56 | 0.12 | - | 0.39 | 1.92 | 0.055 | - | 0.37 |
| **Pz** | | | | | | | | | | | | | | | | | | | | | | | |
| Count | 82.68 | 121.32 | 122.5 | 234.63 | 30.87 | 54.8 | 3.60 | 2 | 97 | 0.16 | - | -0.72 | 0.46 | - | -0.21 | 1.85 | 0.06 | - | 0.62 | 1.07 | 0.29 | - | 0.6 |
| Density | 0.14 | 0.23 | 0.25 | 0.49 | 0.04 | 0.09 | 3.44 | 2 | 97 | 0.17 | - | -0.09 | 0.92 | - | -0.28 | 1.56 | 0.12 | - | 0.67 | 1.5 | 0.13 | - | 0.62 |
| Duration | 1.29 | 0.16 | 1.31 | 0.17 | 1.42 | 0.17 | 11.37 | 2 | 97 | 0.003 | **0.01*** | -0.31 | 0.75 | - | -0.13 | -2.65 | 0.008 | **0.02*** | -0.64 | -2.95 | 0.003 | **0.009*** | -0.8 |
| Amplitude | 188.79 | 13.91 | 198.73 | 49.94 | 194.97 | 29.80 | 0.10 | 2 | 97 | 0.95 | - | 0.3 | 0.76 | - | -0.26 | -0.25 | 0.8 | - | 0.1 | 0.09 | 0.92 | - | -0.24 |
| Frequency | 0.69 | 0.11 | 0.67 | 0.10 | 0.62 | 0.095 | 7.26 | 2 | 97 | **0.026** | 0.053 | 0.66 | 0.51 | - | 0.23 | 1.83 | 0.07 | - | 0.46 | 2.53 | 0.01 | **0.03*** | 0.68 |

SO, slow oscillation

#### Table S11 – Chronic sedative-hypnotic use affects SO-spindle association and SO-sigma PAC during NREM

| Outcome measure | GS | | INS | | MED | | GS vs INS vs MED Kruskal-Wallis test or ANOVA | | | | | GS vs INS post hoc - Dunn test or t test | | | effect size | INS vs MED post hoc - Dunn test or t test | | | effect size | GS vs MED post hoc - Dunn test or t test | | | effect size |
|---|---|---|---|---|---|---|---|---|---|---|---|---|---|---|---|---|---|---|---|---|---|---|---|
| | Mean | SD | Mean | SD | Mean | SD | F | df | residual | *p* | *q* | *t* | *p* | *q* | g' | *t* | *p* | *q* | g' | *t* | *p* | *q* | g' |
| ***Temporal association*** | | | | | | | | | | | | | | | | | | | | | | | |
| **Fz** | | | | | | | | | | | | | | | | | | | | | | | |
| Recall | 0.12 | 0.03 | 0.11 | 0.03 | 0.12 | 0.04 | 4.78 | 2 | 97 | 0.09 | - | - | - | - | - | - | - | - | - | - | - | - | - |
| Precision | 0.39 | 0.10 | 0.37 | 0.07 | 0.35 | 0.11 | 1.33 | 2 | 97 | 0.27 | - | - | - | - | - | - | - | - | - | - | - | - | - |
| **Cz** | | | | | | | | | | | | | | | | | | | | | | | |
| Recall | 0.1 | 0.02 | 0.09 | 0.03 | 0.09 | 0.03 | 6.86 | 2 | 97 | **0.032** | 0.053 | 1.64 | 0.10 | - | 0.29 | 0.72 | 0.47 | - | 0.22 | 2.61 | 0.009 | **0.03*** | 0.52 |
| Precision | 0.29 | 0.06 | 0.3 | 0.09 | 0.25 | 0.07 | 5.86 | 2 | 97 | 0.053 | - | 0.07 | 0.94 | - | -0.08 | 1.94 | 0.052 | - | 0.54 | 2.06 | **0.04** | 0.12 | 0.52 |
| **Pz** | | | | | | | | | | | | | | | | | | | | | | | |
| Recall | 0.1 | 0.02 | 0.09 | 0.03 | 0.09 | 0.03 | 7.02 | 2 | 97 | **0.03** | 0.06 | 1.61 | 0.11 | - | 0.35 | 0.79 | 0.42 | - | 0.2 | 2.64 | 0.008 | **0.02*** | 0.55 |
| Precision | 0.24 | 0.05 | 0.24 | 0.07 | 0.21 | 0.07 | 1.75 | 2 | 97 | 0.18 | - | - | - | - | - | - | - | - | - | - | - | - | - |
| ***SO-sigma PAC*** | | | | | | | | | | | | | | | | | | | | | | | |
| Modulation Index | | | | | | | | | | | | | | | | | | | | | | | |
| Fz | 1.63 | 1.12 | 1.22 | 1.32 | 0.99 | 1 | 6.21 | 2 | 97 | **0.04*** | - | 1.71 | 0.09 | - | 0.33 | 0.49 | 0.62 | - | 0.2 | 2.45 | 0.014 | **0.04*** | 0.61 |
| Cz | 2.75 | 1.33 | 2.36 | 1.46 | 1.66 | 1.22 | 12.80 | 2 | 97 | **0.002*** | - | 1.16 | 0.24 | - | 0.27 | 2.09 | **0.037** | 0.11 | 0.53 | 3.47 | < 0.001 | **0.002*** | 0.85 |
| Pz | 3.52 | 1.62 | 2.67 | 1.56 | 1.94 | 1.45 | 17.88 | 2 | 97 | **< 0.001*** | - | 1.84 | 0.07 | - | 0.53 | 2.04 | **0.04** | 0.12 | 0.49 | 4.19 | **< 0.001*** | - | 1.04 |
| Coupling preferred-phase (degree) | | | | | | | | | | | | Watson's Two-Sample Test | | | | Watson's Two-Sample Test | | | | Watson's Two-Sample Test | | | |
| Fz | 279 | 53 | 288 | 34 | 301 | 55 | - | - | - | - | - | 0.07 | > 0.10 | - | - | 0.07 | > 0.10 | - | - | 0.18 | > 0.05 | - | - |
| Cz | 232 | 31 | 228 | 37 | 263 | 33 | - | - | - | - | - | 0.05 | > 0.10 | - | - | 0.41 | **< 0.01*** | - | -1.01 | 0.26 | **< 0.05*** | - | -0.97 |
| Pz | 236 | 27 | 238 | 31 | 265 | 27 | - | - | - | - | - | 0.05 | > 0.10 | - | -0.06 | 0.38 | **< 0.01*** | - | -0.95 | 0.30 | **< 0.01*** | - | -1.07 |

SO, slow oscillation; PAC, phase-amplitude coupling

#### Table S12 : Association between the Modulation Index and changes in SOs and spindles characteristics during NREM

| Outcome measure | MODULATION INDEX Spearman correlation test | | |
|---|---|---|---|
| | *p* | *q* | r |
| Frontal | | | |
| ***Spindle characteristics*** | | | |
| Density | 0.35 | - | - |
| Amplitude | **0.03** | 0.08 | 0.31 |
| ***SO characteristics*** | | | |
| Duration | 0.50 | - | - |
| Amplitude | <0.001 | **0.003*** | 0.49 |
| Frequency | 0.38 | - | - |
| Parietal | | | |
| ***Spindle characteristics*** | | | |
| Amplitude | **<0.001** | **<0.001*** | 0.56 |
| ***SO characteristics*** | | | |
| Amplitude | <0.001 | **<0.001*** | 0.63 |

SO, slow oscillation

#### Figure S2: Chronic sedative-hypnotic use is associated with alteration in the modulation index

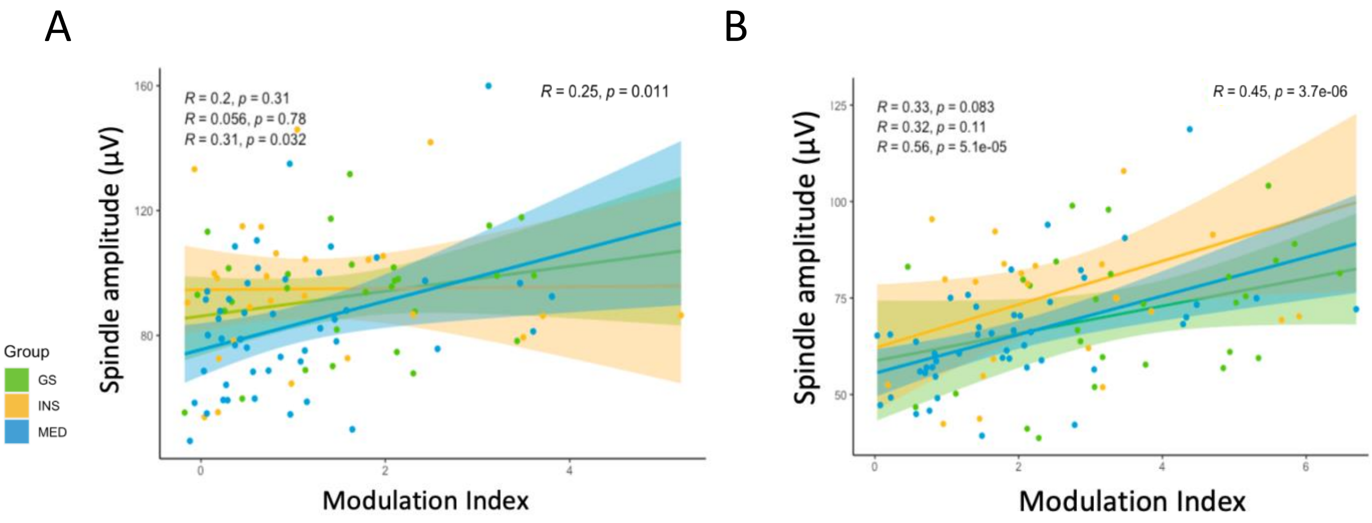


(A) Scatterplot showing correlation between the change in the Modulation Index and the change in the spindle amplitude in Fz for the GS group (green), the INS group (orange), and the MED (blue) group.
(B) Scatterplot showing correlation between the change in the Modulation Index and the change in the spindle amplitude in Pz for the GS group (green), the INS group (orange), and the MED (blue) group.

#### Subgroup analysis: effect on the use and type of chronic sedative-hypnotic exposure on sleep.

A subgroup analysis was conducted within the MED group, where individuals were classified into BZD (n=18) or BZRA (n=29) group. While we found a group effect driven by BZD which displayed greater sedative-hypnotic dose equivalent in diazepam (*p*=.002) compared to the BZRA group, we did not find any significant group effect on age, sex or duration of hypnotics consumption between the two groups (all p>.05).

Within the MED group, we found correlations between higher sedative-hypnotic dosage and decreased TST (*p*=.02, *q*=.07, *r*=-.33), increased SOL (*p*=.003, *q*=.01, *r*=.45), and latency to reach stage N2 (*p*=.002, *q*=.01, *r*=.44), N3 (*p*<.001, *r*=.51 – **Figure 4A**) and REM (*p*=.04, *q*=.08, *r*=.31). Additionally, we found associations between higher hypnotics dosage, and decreased TSP (*p*=.009, *q*=.04, *r*=-.38), as increased arousal density in REM (*p*=.03, *q*=.08, *r*=.31) although it did not pass multiple comparison correction. Additionally, we observed a correlation between higher sedative-hypnotic dosage and increased high-frequency bands relative power as sigma (*p*=.004, *q*=.01, *r*=.41 – **Figure 4B**) and both low and high beta (all *p*=.003, all *q*=.01, *r*=.42 – **Figure 4C**) in frontal area during NREM. Higher sedative-hypnotic dosage were also associated with increased low beta relative power (*p*=.04, *q*=.2, *r*=.29) in parietal region; although it did not pass multiple comparisons correction. We observed a correlation between higher sedative-hypnotic dosage and lower low-frequency bands beta ratio as SO (*p*=.01, *q*=.03, *r*=-.37), delta (*p*=.003, *q*=.02, *r*=-.43) in frontal and theta in both frontal (*p*=.007, *q*=.02, *r*=-.39) and parietal (*p*=.02, *q*=.1, *r*=-.34); although it did not pass multiple comparisons correction.

We found correlations in Fz between higher sedative-hypnotic dosage and increased SO duration (*p*=.046, *q*=.09, *r*=.29) and decreased SO amplitude (*p*=.016, *q*=.06, *r*=-.34); although it did not pass multiple comparisons correction. We did not find any association between sedative-hypnotic dosage and spindle characteristics, as well as temporal co-occurrence of SO and spindle, and SO-sigma PAC. Although it did not pass multiple comparisons, we observed association between higher dose and decreased SFI (*p*=.021, *q*=.38, *r*=-.34), and SSI (*p*=.045, *q*=.41, *r*=-.29). We observed a correlation between longer sedative-hypnotic consumption duration and decreased spindle duration (*p*=.004, *q*=.02, *r*=-.41 – **Figure 4D**) in Pz. We found no correlation between the duration of sedative-hypnotic and others measures of sleep architecture, SOs and spindles characteristics as their temporal co-occurrence and SO-sigma PAC.

We observed that the BZD group drove the alterations in sleep architecture compared to the BZRA group, where they displayed lower TST (*F*(2,44)=6.1; *p*=0.02, *q*=0.2), SE (*F*(2,44)=4.4; *p*=0.04, *q*=0.24), and SL to N3 (*W*(2,44)=143; *p*=0.03, *q*=0.2), although it did not pass multiple comparison correction (**Table S13**). We found a group effect on high-frequency bands in the frontal area driven by the BZD group, which displayed greater low and high beta (all *p*=.02, all *q*=.07) relative power compared to the BZRA group (**Table S14**). We also found a decrease in both delta- and theta-beta ratio in Fz driven by the BZD group, although it did not pass multiple comparison correction (all *p*<.02, all *q*=.07). We noticed a group effect on SO amplitude (*F*(2,44)=161; *p*=0.03, *q*=0.08) in Fz and SO frequency (*F*(2,44)=176; *p*=0.04, *q*=0.10) in Pz driven by the BZD group, where those characteristics were lower compared to the BZRA group, although it did not pass multiple comparison correction (**Table S15**). We did not find significative difference between BZD and BZRA regarding spindle characteristics, as well as SO-spindle coupling (**Table S16**).

#### Table S13 – Effect of chronic sedative-hypnotic exposure on sleep architecture

| Outcome measure | BZD n=18 | | BZRA n=29 | | MEDICATION TYPE (BZD vs BZRA): ANOVA or *Wilcoxon test* | | | | MEDICATION TYPE (BZD vs BZRA): effect size | Covariable: MEDICATION EQUIVALENT DOSE Spearman correlation test or ANCOVA: BZD group | | Covariable: MEDICATION EQUIVALENT DOSE Spearman correlation test or ANCOVA: BZRA group | | MEDICATION DOSE Spearman correlation test | | | MEDICATION DURATION Spearman correlation test | | |
|---|---|---|---|---|---|---|---|---|---|---|---|---|---|---|---|---|---|---|---|
| | Mean | SD | Mean | SD | F | df | p | q | g' | p | r | p | r | p | q | r | p | q | r |
| ***Sleep duration*** | | | | | | | | | | | | | | | | | | | |
| TSP (min) | 440.17 | 68.2 | 480.05 | 49.66 | 0.31 | 44 | 0.58 | - | - | **F(2,44)=6.7; p=0.02*** | | | | **0.041*** | - | -0.38 | 0.38 | - | 0.13 |
| TST (min) | 342.36 | 56.21 | 408.1 | 47.01 | 6.06 | 44 | **0.02** | 0.24 | -0.9 | **F(2,44)=4.9; p=0.03*** | | | | **0.02** | 0.07 | -0.33 | 0.95 | - | -0.01 |
| N1 (% TSP) | 5.01 | 3.16 | 4.68 | 1.96 | *261* | *44* | *1* | - | - | 0.39 | -0.22 | 0.31 | 0.20 | 0.75 | - | 0.05 | 0.39 | - | -0.13 |
| N2 (% TSP) | 50.79 | 11.78 | 51.47 | 6.99 | 0.01 | 44 | 0.91 | - | - | F(2,44)=0.02; p=0.88 | | | | 0.80 | - | -0.04 | 0.79 | - | 0.0 |
| N3 (% TSP) | 5.31 | 4.65 | 8.69 | 5.49 | 2.76 | 44 | 0.10 | - | - | F(2,44)=0.04; p=0.85 | | | | 0.13 | - | -0.22 | 0.29 | - | -0.16 |
| REM (% TSP) | 17.7 | 6.95 | 20.25 | 5.73 | 2.69 | 44 | 0.11 | - | - | F(2,44)=0.83; p=0.37 | | | | 0.91 | - | -0.02 | 0.39 | - | 0.13 |
| Wake (% TSP) | 21.19 | 13.21 | 14.91 | 5.66 | 3.82 | 44 | 0.06 | - | - | F(2,44)=0.04; p=0.84 | | | | 0.39 | - | 0.13 | 0.72 | - | 0.05 |
| ***Sleep Initiation (min)*** | | | | | | | | | | | | | | | | | | | |
| SOL | 40.47 | 51.08 | 18.62 | 21.77 | *185* | *44* | *0.1* | - | - | 0.47 | 0.18 | **0.04*** | 0.38 | **0.01*** | - | 0.45 | 0.67 | - | 0.06 |
| SL to N2 | 42.83 | 50.8 | 21.16 | 21.63 | *183* | *44* | *0.09* | - | - | 0.36 | 0.23 | **0.03*** | 0.40 | **0.01*** | - | 0.44 | 0.74 | - | 0.05 |
| SL to N3 | 75.21 | 59.93 | 45.23 | 31.42 | *143* | *44* | ***0.03*** | 0.24 | 0.67 | **0.04*** | 0.49 | 0.13 | 0.29 | **< 0.001*** | - | 0.51 | 0.61 | - | 0.08 |
| SL to REM | 167.39 | 101.5 | 121.12 | 54.29 | *190* | *44* | *0.12* | - | - | 0.33 | 0.24 | 0.99 | 0 | **0.04** | 0.08 | 0.31 | 0.71 | - | -0.06 |
| ***Sleep Fragmentation*** | | | | | | | | | | | | | | | | | | | |
| WASO (% TIB) | 18.92 | 12.31 | 14.03 | 5.5 | *218.5* | *44* | *0.36* | - | - | 0.70 | -0.09 | 0.94 | 0.01 | 0.67 | - | 0.06 | 0.79 | - | 0.04 |
| SE | 70.41 | 14.5 | 79.96 | 6.76 | 4.38 | 44 | **0.04** | 0.24 | -0.9 | F(2,44)=0.54; p=0.47 | | | | **0.02** | 0.07 | -0.33 | 0.95 | - | -0.01 |
| Arousal density Total | 14.92 | 6.68 | 14.51 | 6.86 | 0.05 | 44 | 0.82 | - | - | F(2,44)=0.50; p=0.49 | | | | 0.48 | - | 0.11 | 0.75 | - | -0.05 |
| Arousal density NREM | 0.23 | 0.1 | 0.25 | 0.13 | 0.15 | 44 | 0.70 | - | - | F(2,44)=0.04; p=0.84 | | | | 0.79 | - | 0.04 | 0.72 | - | -0.05 |
| Arousal density REM | 0.28 | 0.19 | 0.21 | 0.12 | *193* | *44* | *0.14* | - | - | 0.49 | 0.18 | 0.08 | 0.33 | **0.03** | 0.08 | 0.31 | 0.39 | - | 0.13 |
| SFI | 9.64 | 4.2 | 9.16 | 1.85 | *269* | *44* | *0.87* | - | - | 0.92 | -0.02 | 0.71 | -0.07 | 0.87 | - | -0.03 | **0.02** | 0.38 | -0.34 |
| SSI | 18.07 | 7.19 | 19.09 | 3.89 | 0.14 | 44 | 0.71 | - | - | F(2,44)=0.07; p=0.80 | | | | 0.49 | - | -0.10 | **0.045** | 0.41 | -0.29 |

TSP, total sleep period; TiB, time in bed; TST, total sleep time; SOL, sleep onset latency; SL, sleep latency; WASO, wake after sleep onset; SE, sleep efficiency; SFI, sleep fragmentation index; SSI, stage switch index; NREM, non-rapid eye movement; REM, rapid eye movement

#### Table S14 – Effect of chronic sedative-hypnotic exposure on spectral activity during NREM

| Outcome measure | BZD n=18 | | BZRA n=29 | | MEDICATION TYPE (BZD vs BZRA) ANOVA or *Wilcoxon test* | | | | effect size | Covariable: MEDICATION EQUIVALENT DOSE Spearman correlation test or ANCOVA BZD group | | BZRA group | | MEDICATION DOSE Spearman correlation test | | | MEDICATION DURATION Spearman correlation test | | |
|---|---|---|---|---|---|---|---|---|---|---|---|---|---|---|---|---|---|---|---|
| | Mean | SD | Mean | SD | F | df | p | q | g' | p | r | p | r | p | q | r | p | q | r |
| ***Relative power spectrum*** | | | | | | | | | | | | | | | | | | | |
| **Fz** | | | | | | | | | | | | | | | | | | | |
| SO | 0.6 | 0.1 | 0.61 | 0.1 | 250 | 44 | 0.82 | - | 0.04 | 0.61 | -0.13 | 0.99 | -0.002 | 0.84 | | 0.03 | 0.25 | - | 0.17 |
| Delta | 0.73 | 0.08 | 0.74 | 0.09 | 0.21 | 44 | 0.65 | - | 0.18 | F(2,44)=2.8; p=0.10 | | | | 0.14 | - | -0.22 | 0.27 | - | 0.16 |
| Theta | 0.06 | 0.02 | 0.06 | 0.03 | 1.07 | 44 | 0.31 | - | 0.2 | F(2,44)=0.7; p=0.42 | | | | 0.96 | - | -0.007 | 0.36 | - | -0.14 |
| Alpha | 0.05 | 0.03 | 0.05 | 0.03 | 244 | 44 | 0.72 | - | -0.05 | 0.13 | 0.37 | 0.34 | 0.18 | 0.10 | - | 0.24 | 0.33 | - | -0.14 |
| Sigma | 0.04 | 0.02 | 0.03 | 0.02 | 0.8 | 44 | 0.38 | - | -0.75 | **F(2,44)=5.4; p=0.03*** | | | | **0.01*** | - | 0.41 | 0.93 | - | -0.01 |
| Low Beta | 0.01 | 0.01 | 0 | 0 | 152 | 44 | **0.02** | 0.07 | -0.78 | 0.48 | 0.18 | 0.05 | 0.37 | **0.01*** | - | 0.42 | 0.91 | - | -0.02 |
| High Beta | 0.01 | 0.01 | 0.01 | 0 | 153 | 44 | **0.02** | 0.07 | -0.82 | 0.45 | 0.19 | 0.05 | 0.36 | **0.01*** | - | 0.42 | 0.77 | - | -0.05 |
| **Pz** | | | | | | | | | | | | | | | | | | | |
| SO | 0.61 | 0.13 | 0.59 | 0.11 | 0.68 | 44 | 0.41 | - | -0.22 | F(2,44)=0.2; p=0.70 | | | | 0.83 | - | 0.032 | 0.56 | - | -0.09 |
| Delta | 0.68 | 0.07 | 0.69 | 0.09 | 0.23 | 44 | 0.63 | - | 0.11 | F(2,44)=1.9; p=0.17 | | | | 0.14 | - | -0.216 | 0.96 | - | -0.007 |
| Theta | 0.06 | 0.02 | 0.08 | 0.04 | 2 | 44 | 0.16 | - | 0.46 | F(2,44)=0.06; p=0.82 | | | | 0.46 | - | -0.11 | 0.96 | - | 0.007 |
| Alpha | 0.04 | 0.03 | 0.05 | 0.03 | 294 | 44 | 0.47 | - | 0.23 | 0.25 | 0.28 | 0.39 | 0.16 | 0.38 | - | 0.131 | 0.96 | - | 0.007 |
| Sigma | 0.06 | 0.05 | 0.04 | 0.03 | 201 | 44 | 0.19 | - | -0.53 | 0.75 | 0.07 | 0.09 | 0.31 | 0.17 | - | 0.287 | 0.66 | - | 0.06 |
| Low Beta | 0.01 | 0.01 | 0.01 | 0 | 207 | 44 | 0.24 | - | -0.51 | 0.47 | 0.17 | 0.06 | 0.34 | **0.04** | 0.17 | 0.298 | 0.37 | - | 0.13 |
| High Beta | 0.01 | 0.01 | 0.01 | 0.01 | 214 | 44 | 0.30 | - | -0.45 | 0.76 | 0.07 | 0.11 | 0.30 | 0.11 | - | 0.239 | 0.99 | - | 0.002 |
| ***Beta ratio*** | | | | | | | | | | | | | | | | | | | |
| **Fz** | | | | | | | | | | | | | | | | | | | |
| SO | 40.7 | 25.76 | 61.71 | 51.4 | 347 | 44 | 0.06 | - | 0.04 | 0.34 | -0.23 | 0.09 | -0.31 | **0.03*** | - | -0.36 | 0.54 | - | 0.09 |
| Delta | 48.89 | 32.01 | 71.57 | 39.8 | 369 | 44 | **0.02** | 0.07 | 0.18 | 0.28 | -0.26 | 0.06 | -0.35 | **0.02*** | - | -0.43 | 0.70 | - | 0.06 |
| Theta | 3.18 | 1.41 | 5.52 | 3.66 | 374 | 44 | **0.01** | 0.07 | 0.2 | 0.37 | -0.22 | 0.11 | -0.29 | **0.02*** | - | -0.39 | 0.44 | - | -0.12 |
| Alpha | 2.49 | 1.14 | 3.93 | 2.95 | 330 | 44 | 0.13 | - | -0.05 | 0.30 | 0.25 | 0.49 | -0.13 | 0.44 | - | -0.12 | 0.96 | - | 0.008 |
| Sigma | 2.15 | 0.68 | 2.21 | 0.87 | 271 | 44 | 0.84 | - | -0.75 | 0.43 | 0.19 | 0.39 | -0.16 | 0.92 | - | -0.02 | 0.75 | - | 0.05 |
| Low Beta | 0.34 | 0.06 | 0.34 | 0.05 | 0.99 | 44 | 0.33 | - | -0.78 | F(2,44)=2.6; p=0.12 | | | | 0.46 | - | 0.11 | 0.37 | - | 0.13 |
| High Beta | 0.66 | 0.06 | 0.66 | 0.05 | 0.99 | 44 | 0.33 | - | -0.82 | F(2,44)=2.6; p=0.12 | | | | 0.46 | - | -0.11 | 0.37 | - | -0.13 |
| **Pz** | | | | | | | | | | | | | | | | | | | |
| SO | 47.91 | 41.77 | 54.12 | 56.65 | 288 | 44 | 0.56 | - | 0.12 | 0.90 | -0.03 | 0.06 | -0.36 | 0.14 | - | -0.22 | 0.71 | - | -0.06 |
| Delta | 49.22 | 33.32 | 58.26 | 37.09 | 310 | 44 | 0.28 | - | 0.25 | 0.66 | -0.11 | 0.06 | -0.35 | 0.06 | - | -0.29 | 0.66 | - | -0.07 |
| Theta | 3.53 | 1.66 | 5.65 | 3.56 | 351 | 44 | 0.05 | - | 0.7 | 0.39 | -0.21 | 0.35 | -0.17 | **0.02** | 0.12 | -0.34 | 0.48 | - | -0.11 |
| Alpha | 2.05 | 0.98 | 3.19 | 2.58 | 337 | 44 | 0.09 | - | 0.53 | 0.43 | 0.19 | 0.95 | 0.01 | 0.62 | - | -0.08 | 0.79 | - | 0.04 |
| Sigma | 2.95 | 1.03 | 2.74 | 0.98 | 0.16 | 44 | 0.69 | - | -0.21 | F(2,44)=0.09; p=0.76 | | | | 0.68 | - | 0.06 | 0.88 | - | 0.02 |
| Low Beta | 0.34 | 0.06 | 0.34 | 0.06 | 0.65 | 44 | 0.42 | - | -0.04 | F(2,44)=2.6; p=0.12 | | | | 0.24 | - | 0.18 | **0.03** | 0.09 | 0.33 |
| High Beta | 0.66 | 0.06 | 0.66 | 0.06 | 0.65 | 44 | 0.42 | - | 0.04 | F(2,44)=2.6; p=0.12 | | | | 0.24 | - | -0.18 | **0.03** | 0.09 | -0.33 |

SO, slow oscillation

#### Table S15 – Effect of chronic sedative-hypnotic exposure on SOs and spindles during NREM

| Outcome measure | BZD n=18 | | BZRA n=29 | | MEDICATION TYPE (BZD vs BZRA) ANOVA or *Wilcoxon test* | | | | effect size | Covariable: MEDICATION EQUIVALENT DOSE Spearman correlation test or ANCOVA BZD group | | BZRA group | | MEDICATION DOSE Spearman correlation test | | | MEDICATION DURATION Spearman correlation test | | |
|---|---|---|---|---|---|---|---|---|---|---|---|---|---|---|---|---|---|---|---|
| | Mean | SD | Mean | SD | F | df | p | q | g' | p | r | p | r | p | q | r | p | q | r |
| ***SO characteristics*** | | | | | | | | | | | | | | | | | | | |
| **Fz** | | | | | | | | | | | | | | | | | | | |
| Density (Nber/30 sec) | 3.88 | 0.45 | 3.79 | 0.84 | 0.57 | 44 | 0.45 | - | 0.13 | F(2,44)=0.5; p=0.50 | | | | 0.26 | - | -0.17 | 0.83 | - | 0.03 |
| Duration (sec) | 1.43 | 0.07 | 1.39 | 0.08 | 342.5 | 44 | 0.05 | - | 0.56 | 0.44 | 0.14 | 0.38 | 0.21 | 0.05 | - | 0.29 | 0.83 | - | 0.03 |
| Amplitude (µV) | 95.63 | 31.23 | 110.19 | 25.5 | 161 | 44 | **0.03** | 0.08 | -0.51 | 0.06 | -0.34 | 0.56 | -0.14 | **0.02** | 0.06 | -0.35 | 0.49 | - | -0.10 |
| Frequency (Hz) | 0.67 | 0.05 | 0.71 | 0.08 | 184 | 44 | 0.06 | - | -0.6 | 0.22 | -0.23 | 0.88 | -0.03 | 0.07 | - | -0.27 | 0.77 | - | -0.04 |
| **Pz** | | | | | | | | | | | | | | | | | | | |
| Density (Nber/30 sec) | 3.53 | 0.56 | 3.48 | 0.74 | 0.14 | 44 | 0.71 | - | 0.07 | F(2,44)=0.09; p=0.77 | | | | 0.36 | - | -0.14 | 0.56 | - | -0.08 |
| Duration (sec) | 1.47 | 0.05 | 1.44 | 0.07 | 322 | 44 | 0.13 | - | 0.44 | 0.84 | 0.04 | 0.55 | 0.14 | 0.25 | - | 0.17 | 0.43 | - | -0.12 |
| Amplitude (µV) | 80.44 | 27.02 | 87.29 | 17.03 | 172 | 44 | 0.05 | - | -0.31 | 0.10 | -0.30 | 0.86 | -0.04 | 0.21 | - | -0.28 | 0.20 | - | -0.19 |
| Frequency (Hz) | 0.62 | 0.04 | 0.66 | 0.07 | 176 | 44 | **0.04** | .10 | -0.62 | 0.54 | -0.11 | 0.71 | 0.09 | 0.18 | - | -0.19 | 0.94 | - | -0.01 |
| ***Spindle characteristics*** | | | | | | | | | | | | | | | | | | | |
| **Fz** | | | | | | | | | | | | | | | | | | | |
| Density (Nber/30 sec) | 1.36 | 0.28 | 1.24 | 0.23 | 0.05 | 44 | 0.82 | - | 0.46 | F(2,44)=3.7; p=0.06 | | | | 0.08 | - | 0.26 | 0.27 | - | -0.16 |
| Duration (sec) | 0.78 | 0.05 | 0.76 | 0.05 | 339.5 | 44 | 0.08 | - | 0.45 | 0.71 | -0.07 | 0.76 | 0.07 | 0.46 | - | 0.11 | 0.06 | - | -0.27 |
| Amplitude (µV) | 77.27 | 20.93 | 86.78 | 21.53 | 178 | 44 | 0.07 | - | -0.44 | 0.05 | -0.36 | 0.93 | -0.02 | 0.07 | - | -0.26 | 0.27 | - | -0.16 |
| Frequency (Hz) | 11.42 | 0.49 | 11.32 | 0.42 | 1.07 | 44 | 0.30 | - | 0.22 | F(2,44)=0.6; p=0.45 | | | | 0.96 | - | 0.009 | 0.97 | - | 0.01 |
| **Pz** | | | | | | | | | | | | | | | | | | | |
| Density (Nber/30 sec) | 1.6 | 0.36 | 1.41 | 0.34 | 2.05 | 44 | 0.15 | - | 0.54 | F(2,44)=0.01; p=0.91 | | | | 0.21 | - | 0.19 | 0.15 | - | -0.21 |
| Duration (sec) | 0.77 | 0.07 | 0.74 | 0.05 | 305.5 | 44 | 0.33 | - | 0.42 | 0.69 | 0.07 | 0.65 | -0.11 | 0.75 | - | 0.05 | 0.004 | **0.02*** | -0.41 |
| Amplitude (µV) | 62.75 | 16.48 | 66.73 | 12.83 | 187 | 44 | 0.10 | - | -0.27 | 0.18 | -0.25 | 0.96 | -0.01 | 0.24 | - | -0.18 | 0.16 | - | -0.21 |
| Frequency (Hz) | 13.63 | 0.67 | 13.4 | 0.53 | 314 | 44 | 0.25 | - | 0.38 | 0.55 | 0.11 | 0.53 | 0.15 | 0.14 | - | 0.22 | 0.92 | - | -0.02 |

SO, slow oscillation

#### Table S16 – Effect of chronic sedative-hypnotic exposure on SO-spindle association and SO-sigma PAC during NREM

| Outcome measure | BZD n=18 | | BZRA n=29 | | MEDICATION TYPE (BZD vs BZRA) ANOVA or *Wilcoxon test* | | | | effect size | Covariable: MEDICATION EQUIVALENT DOSE Spearman correlation test or ANCOVA BZD group | | BZRA group | | MEDICATION DOSE Spearman correlation test | | | MEDICATION DURATION Spearman correlation test | | |
|---|---|---|---|---|---|---|---|---|---|---|---|---|---|---|---|---|---|---|---|
| | Mean | SD | Mean | SD | F | df | *p* | *q* | g' | *p* | *r* | *p* | *r* | *p* | *q* | r | *p* | *q* | *r* |
| ***Temporal association*** | | | | | | | | | | | | | | | | | | | |
| **Fz** | | | | | | | | | | | | | | | | | | | |
| Recall (SO$^+$) | 0.12 | 0.04 | 0.11 | 0.04 | 278 | 44 | 0.71 | - | 0.19 | 0.49 | -0.13 | 0.40 | 0.20 | 0.81 | - | 0.03 | 0.23 | - | -0.17 |
| Precision (Spindle$^+$) | 0.34 | 0.08 | 0.35 | 0.12 | 0.02 | 44 | 0.89 | - | -0.1 | F(2,44)=0.5; p=0.49 | | | | 0.18 | - | -0.19 | 0.96 | - | -0.01 |
| **Pz** | | | | | | | | | | | | | | | | | | | |
| Recall (SO$^+$) | 0.1 | 0.04 | 0.08 | 0.02 | 332.5 | 44 | 0.11 | - | 0.49 | 0.52 | 0.12 | 0.54 | -0.15 | 0.47 | - | 0.10 | 0.06 | - | -0.28 |
| Precision (Spindle$^+$) | 0.21 | 0.08 | 0.2 | 0.07 | 0.61 | 44 | 0.43 | - | 0.06 | F(2,44)=1.2; p=0.30 | | | | 0.36 | - | -0.13 | 0.28 | - | -0.15 |
| ***Phase-Amplitude coupling*** | | | | | | | | | | | | | | | | | | | |
| **Fz** | | | | | | | | | | | | | | | | | | | |
| Modulation Index | 0.71 | 0.87 | 1.16 | 1.04 | 333 | 44 | 0.11 | - | 0.45 | 0.17 | -0.33 | 0.95 | -0.01 | 0.22 | - | -0.18 | 0.78 | - | 0.04 |
| **Pz** | | | | | | | | | | | | | | | | | | | |
| Modulation Index | 1.74 | 1.55 | 2.06 | 1.39 | 318 | 44 | 0.21 | - | 0.22 | 0.90 | 0.03 | 0.93 | -0.01 | 0.62 | - | -0.07 | 0.26 | - | -0.16 |

SO, slow oscillation

#### Table S17 – Effect of chronic BZRA use on sleep architecture

| Outcome measure | GS | | INS | | BZRA | | GS vs INS vs MED Kruskal-Wallis test or ANOVA | | | | | GS vs INS post hoc - Dunn test or t test | | | effect size | INS vs MED post hoc - Dunn test or t test | | | effect size | GS vs MED post hoc - Dunn test or t test | | | effect size |
|---|---|---|---|---|---|---|---|---|---|---|---|---|---|---|---|---|---|---|---|---|---|---|---|
| | Mean | SD | Mean | SD | Mean | SD | F | df | esidua | *p* | *q* | *t* | *p* | *q* | g' | *t* | *p* | *q* | g' | *t* | *p* | *q* | g' |
| Sleep architecture | | | | | | | | | | | | | | | | | | | | | | | |
| ***Sleep duration*** | | | | | | | | | | | | | | | | | | | | | | | |
| TSP (% TIB) | 94.59 | 3.96 | 92.12 | 7.5 | 93.65 | 5.42 | 0.48 | 2 | 75 | 0.78 | - | - | - | - | - | - | - | - | - | - | - | - | - |
| SE (%) | 85.12 | 8.47 | 75.52 | 10.8 | 80 | 6.89 | 15.16 | 2 | 75 | < 0.002 | **0.001*** | 3.78 | **< 0.001*** | - | 0.98 | -1.15 | 0.25 | - | -0.49 | 2.63 | 0.008 | **0.025*** | 0.65 |
| N1 (% TSP) | 2.49 | 1.32 | 1.98 | 0.97 | 4.75 | 1.91 | 34.70 | 2 | 75 | **< 0.001*** | - | 1.19 | 0.23 | - | 0.42 | -5.55 | **< 0.001*** | - | -1.79 | -4.51 | **< 0.001*** | - | -1.36 |
| N2 (% TSP) | 46.17 | 8.53 | 35.08 | 7.7 | 51.79 | 7.23 | *29.83* | 2 | 75 | **< 0.001*** | - | *5.128* | **< 0.001*** | - | *1.34* | *-2.62* | **< 0.001*** | - | *-2.2* | *-2.62* | *0.01* | **0.03*** | *-0.7* |
| N3 (% TSP) | 19.8 | 9.2 | 23.25 | 6.32 | 8.73 | 5.52 | *25.5* | 2 | 75 | **< 0.001*** | - | *-1.726* | *0.08* | - | *-0.43* | *5.59* | **< 0.001*** | - | *2.41* | *5.59* | **< 0.001*** | - | *1.43* |
| REM (% TSP) | 21.37 | 5.24 | 21.57 | 6.76 | 20.14 | 5.74 | *0.02* | 2 | 75 | *0.97* | - | - | - | - | - | - | - | - | - | - | - | - | - |
| Wake (% TSP) | 10.17 | 6.47 | 18.12 | 9.04 | 14.59 | 5.38 | 15.62 | 2 | 75 | < 0.001 | **0.001*** | -3.77 | **< 0.001*** | - | -1.01 | 0.96 | 0.34 | - | 0.47 | -2.83 | 0.005 | **0.01*** | -0.73 |
| ***Sleep Initiation*** (min) | | | | | | | | | | | | | | | | | | | | | | | |
| SOL | 3.35 | 3.17 | 4.4 | 5.65 | 3.8 | 4.59 | 0.16 | 2 | 75 | 0.92 | - | - | - | - | - | - | - | - | - | - | - | - | - |
| SL to N2 | 17.37 | 15.49 | 23.37 | 33.61 | 21.65 | 22.66 | 0.58 | *2* | 75 | 0.74 | - | - | - | - | - | - | - | - | - | - | - | - | - |
| SL to N3 | 32.41 | 25.13 | 42.27 | 39.78 | | | 4.87 | 2 | 75 | 0.08 | - | - | - | - | - | - | - | - | - | - | - | - | - |
| SL to REM | 90.79 | 33.53 | 94.97 | 42.83 | 120.12 | 55.47 | 5.96 | *2* | 75 | **0.051** | - | -3.77 | **< 0.001*** | - | -1.01 | 0.96 | 0.33 | - | 0.47 | -2.83 | 0.005 | **0.01*** | -0.73 |
| ***Sleep Fragmentation*** | | | | | | | | | | | | | | | | | | | | | | | |
| WASO (% TIB) | 9.47 | 5.61 | 16.6 | 8.22 | 13.66 | 5.14 | 15.57 | 2 | 75 | < 0.001 | **0.001*** | -3.76 | **< 0.001*** | - | -1.01 | 0.94 | 0.34 | - | 0.42 | -2.84 | 0.005 | **0.01*** | -0.77 |
| Arousal density (Nber/30 sec) | | | | | | | | | | | | | | | | | | | | | | | |
| in NREM+REM | 14.7 | 5.52 | 20.38 | 7.42 | 14.67 | 7.19 | 9.68 | 2 | 75 | 0.008 | **0.01*** | -2.62 | 0.009 | **0.026*** | -0.86 | 2.79 | 0.005 | **0.015*** | 0.77 | 0.22 | 0.82 | - | 0.01 |
| in NREM | 0.25 | 0.11 | 0.34 | 0.14 | 0.25 | 0.14 | 6.88 | 2 | 75 | 0.032 | **0.049*** | -2.05 | **0.04** | 0.12 | -0.73 | 2.46 | 0.01 | **0.04*** | 0.68 | 0.46 | 0.64 | - | 0.03 |
| in REM | 0.21 | 0.09 | 0.31 | 0.14 | 0.21 | 0.13 | 12.13 | 2 | 75 | 0.002 | **0.004*** | -2.97 | 0.003 | **0.009*** | -0.88 | 3.09 | 0.002 | **0.006*** | 0.75 | 0.18 | 0.85 | - | -0.02 |
| SFI | 8.15 | 2.51 | 6.76 | 2.33 | 9.32 | 1.83 | 19.33 | 2 | 75 | < 0.001 | **0.001*** | 2.1 | **0.035** | 0.10 | 0.56 | -4.39 | **< 0.001*** | - | -1.2 | -2.39 | 0.017 | **0.05*** | -0.52 |
| SSI | 17.49 | 3.89 | 13.78 | 4.29 | 19.31 | 3.75 | 22.31 | 2 | 75 | **< 0.001*** | - | 3.3 | 0.001 | **0.003*** | 0.89 | -4.6 | **< 0.001*** | - | -1.36 | -1.4 | 0.16 | - | -0.47 |

TSP, total sleep period; TIB, time in bed; SOL, sleep onset latency; SL, sleep latency; WASO, wake after sleep onset; SE, sleep efficiency; SFI, sleep fragmentation index; SSI, stage switch index; NREM, non-rapid eye movement; REM, rapid eye movement

### Table S18 – Effect of chronic BZRA use on spectral activity during NREM

| Outcome measure | GS | | INS | | BZRA | | GS vs INS vs MED Kruskal-Wallis test or ANOVA | | | | | GS vs INS post hoc - Dunn test or t test | | | effect size | INS vs MED post hoc - Dunn test or t test | | | effect size | GS vs MED post hoc - Dunn test or t test | | | effect size |
|---|---|---|---|---|---|---|---|---|---|---|---|---|---|---|---|---|---|---|---|---|---|---|---|
| | Mean | SD | Mean | SD | Mean | SD | F | df | residual | *p* | *q* | *t* | *p* | *q* | g' | *t* | *p* | *q* | g' | *t* | *p* | *q* | g' |
| ***Relative power spectrum*** | | | | | | | | | | | | | | | | | | | | | | | |
| **Fz** | | | | | | | | | | | | | | | | | | | | | | | |
| SO | 0.59 | 0.1 | 0.59 | 0.1 | 0.6 | 0.1 | 0.96 | 2 | 75 | 0.6 | - | - | - | - | - | - | - | - | - | - | - | - | - |
| Delta | 0.76 | 0.07 | 0.78 | 0.08 | 0.75 | 0.08 | 2.80 | 2 | 75 | 0.25 | - | - | - | - | - | - | - | - | - | - | - | - | - |
| Theta | 0.08 | 0.03 | 0.07 | 0.03 | 0.06 | 0.03 | *1.9* | 2 | 75 | *0.15* | - | - | - | - | - | - | - | - | - | - | - | - | - |
| Alpha | 0.04 | 0.02 | 0.03 | 0.02 | 0.05 | 0.03 | 1.32 | 2 | 75 | 0.51 | - | - | - | - | - | - | - | - | - | - | - | - | - |
| Sigma | 0.02 | 0.01 | 0.02 | 0.01 | 0.03 | 0.02 | 6.48 | 2 | 75 | **0.039** | 0.10 | -0.25 | 0.80 | - | -0.1 | -2.04 | **0.04** | 0.12 | -0.69 | -2.35 | **0.019** | 0.056 | -0.79 |
| Low Beta | 0 | 0 | 0 | 0 | 0 | 0 | 8.02 | 2 | 75 | **0.018** | 0.11 | -0.17 | 0.86 | - | -0.08 | -2.33 | **0.019** | 0.06 | -0.55 | -2.56 | 0.01 | **0.03*** | -0.69 |
| High Beta | 0.01 | 0 | 0.01 | 0 | 0.01 | 0 | 6.28 | 2 | 75 | **0.04** | 0.12 | 0.34 | 0.73 | - | 0.1 | -2.3 | **0.02** | 0.06 | -0.61 | -2.02 | **0.04** | 0.13 | -0.52 |
| **Cz** | | | | | | | | | | | | | | | | | | | | | | | |
| SO | 0.52 | 0.11 | 0.54 | 0.11 | 0.54 | 0.11 | 0.42 | 2 | 75 | 0.80 | - | - | - | - | - | - | - | - | - | - | - | - | - |
| Delta | 0.73 | 0.07 | 0.72 | 0.08 | 0.71 | 0.09 | 0.63 | 2 | 75 | 0.72 | - | - | - | - | - | - | - | - | - | - | - | - | - |
| Theta | 0.1 | 0.03 | 0.09 | 0.03 | 0.08 | 0.03 | 3.37 | 2 | 75 | 0.18 | - | - | - | - | - | - | - | - | - | - | - | - | - |
| Alpha | 0.04 | 0.02 | 0.04 | 0.02 | 0.05 | 0.04 | 1.13 | 2 | 75 | 0.56 | - | - | - | - | - | - | - | - | - | - | - | - | - |
| Sigma | 0.03 | 0.01 | 0.03 | 0.01 | 0.04 | 0.02 | 2.63 | 2 | 75 | 0.26 | - | - | - | - | - | - | - | - | - | - | - | - | - |
| Low Beta | 0 | 0 | 0 | 0 | 0.01 | 0 | 4.60 | 2 | 75 | 0.10 | - | - | - | - | - | - | - | - | - | - | - | - | - |
| High Beta | 0.01 | 0 | 0.01 | 0 | 0.01 | 0.01 | 5.03 | 2 | 75 | 0.08 | - | - | - | - | - | - | - | - | - | - | - | - | - |
| **Pz** | | | | | | | | | | | | | | | | | | | | | | | |
| SO | 0.56 | 0.12 | 0.57 | 0.11 | 0.58 | 0.11 | 1.14 | 2 | 75 | 0.56 | - | - | - | - | - | - | - | - | - | - | - | - | - |
| Delta | 0.69 | 0.08 | 0.72 | 0.1 | 0.69 | 0.08 | *0.67* | 2 | 75 | *0.50* | - | - | - | - | - | - | - | - | - | - | - | - | - |
| Theta | 0.11 | 0.05 | 0.1 | 0.05 | 0.08 | 0.04 | *2.92* | 2 | 75 | *0.06* | - | - | - | - | - | - | - | - | - | - | - | - | - |
| Alpha | 0.05 | 0.02 | 0.04 | 0.03 | 0.05 | 0.03 | 0.71 | 2 | 75 | 0.69 | - | - | - | - | - | - | - | - | - | - | - | - | - |
| Sigma | 0.04 | 0.02 | 0.03 | 0.02 | 0.04 | 0.03 | 1.01 | 2 | 75 | 0.60 | - | - | - | - | - | - | - | - | - | - | - | - | - |
| Low Beta | 0 | 0 | 0 | 0 | 0.01 | 0 | *1.08* | 2 | 75 | *0.34* | - | - | - | - | - | - | - | - | - | - | - | - | - |
| High Beta | 0.01 | 0 | 0.01 | 0 | 0.01 | 0.01 | 4.69 | 2 | 75 | 0.09 | - | - | - | - | - | - | - | - | - | - | - | - | - |
| ***Beta ratio*** | | | | | | | | | | | | | | | | | | | | | | | |
| **Fz** | | | | | | | | | | | | | | | | | | | | | | | |
| SO | 80.27 | 61.4 | 77.74 | 43.38 | 62.77 | 53.69 | 4.97 | 2 | 75 | 0.08 | - | - | - | - | - | - | - | - | - | - | - | - | - |
| Delta | 97.15 | 49.52 | 100 | 52.84 | 72.82 | 40.9 | 6.95 | 2 | 75 | **0.03** | 0.11 | -0.31 | 0.75 | - | -0.06 | 2.4 | 0.016 | **0.049*** | 0.57 | 2.16 | **0.03** | 0.09 | 0.53 |
| Theta | 9.35 | 4.03 | 9.16 | 6.5 | 5.79 | 3.77 | 12.65 | 2 | 75 | 0.001 | **0.01*** | 1.04 | 0.29 | - | 0.04 | 2.37 | 0.018 | **0.05*** | 0.63 | 3.49 | <0.001 | **0.002*** | 0.9 |
| Alpha | 4.78 | 4.64 | 4.21 | 2.65 | 4.06 | 3.09 | 0.95 | 2 | 75 | 0.62 | - | - | - | - | - | - | - | - | - | - | - | - | - |
| Sigma | 2.11 | 0.74 | 2.18 | 0.65 | 2.26 | 0.9 | 0.26 | 2 | 75 | 0.87 | - | - | - | - | - | - | - | - | - | - | - | - | - |
| Low Beta | 0.33 | 0.04 | 0.35 | 0.03 | 0.34 | 0.05 | *0.86* | 2 | 75 | *0.43* | - | - | - | - | - | - | - | - | - | - | - | - | - |
| High Beta | 0.67 | 0.04 | 0.65 | 0.03 | 0.66 | 0.05 | *0.86* | 2 | 75 | *0.43* | - | - | - | - | - | - | - | - | - | - | - | - | - |
| **Cz** | | | | | | | | | | | | | | | | | | | | | | | |
| SO | 51.39 | 40.2 | 48.79 | 29.77 | 42.5 | 42.4 | *0.39* | 2 | 75 | *0.68* | - | - | - | - | - | - | - | - | - | - | - | - | - |
| Delta | 66.01 | 32.41 | 62.4 | 25.36 | 51.4 | 29.56 | 4.442 | 2 | 75 | 0.10 | - | - | - | - | - | - | - | - | - | - | - | - | - |
| Theta | 8.13 | 3.24 | 7.72 | 3.95 | 5.55 | 3.47 | 11.38 | 2 | 75 | 0.003 | **0.02*** | 0.74 | 0.46 | - | 0.11 | 2.45 | 0.01 | **0.04*** | 0.58 | 3.25 | 0.001 | **0.003*** | 0.76 |
| Alpha | 3.59 | 2.34 | 3.37 | 1.73 | 3.36 | 2.66 | 1.72 | 2 | 75 | 0.42 | - | - | - | - | - | - | - | - | - | - | - | - | - |
| Sigma | 2.41 | 0.89 | 2.29 | 0.71 | 2.21 | 0.61 | 0.23 | 2 | 75 | 0.89 | - | - | - | - | - | - | - | - | - | - | - | - | - |
| Low Beta | 0.35 | 0.04 | 0.36 | 0.05 | 0.34 | 0.05 | 1.07 | 2 | 75 | 0.58 | - | - | - | - | - | - | - | - | - | - | - | - | - |
| High Beta | 0.65 | 0.04 | 0.64 | 0.05 | 0.66 | 0.05 | 1.07 | 2 | 75 | 0.58 | - | - | - | - | - | - | - | - | - | - | - | - | - |
| **Pz** | | | | | | | | | | | | | | | | | | | | | | | |
| SO | 57.64 | 45.81 | 61.85 | 45.92 | 53.12 | 57.04 | 2.74 | 2 | 75 | 0.25 | - | - | - | - | - | - | - | - | - | - | - | - | - |
| Delta | 65.86 | 33.86 | 73.5 | 35.36 | 57.02 | 34.28 | 3.98 | 2 | 75 | 0.14 | - | - | - | - | - | - | - | - | - | - | - | - | - |
| Theta | 9.23 | 3.27 | 9.04 | 5.39 | 5.87 | 3.69 | 13.72 | 2 | 75 | 0.001 | **0.007*** | 0.96 | 0.33 | - | 0.04 | 2.56 | 0.01 | **0.03*** | 0.68 | 3.6 | <0.001 | **0.001*** | 0.95 |
| Alpha | 3.66 | 1.69 | 3.76 | 2.76 | 3.28 | 2.71 | 4.31 | 2 | 75 | 0.11 | - | - | - | - | - | - | - | - | - | - | - | - | - |
| Sigma | 3.1 | 1.25 | 2.98 | 0.97 | 2.79 | 0.97 | 0.19 | 2 | 75 | 0.90 | - | - | - | - | - | - | - | - | - | - | - | - | - |
| Low Beta | 0.35 | 0.04 | 0.37 | 0.04 | 0.34 | 0.06 | 4.44 | 2 | 75 | 0.10 | - | - | - | - | - | - | - | - | - | - | - | - | - |
| High Beta | 0.65 | 0.04 | 0.63 | 0.04 | 0.66 | 0.06 | 4.44 | 2 | 75 | 0.10 | - | - | - | - | - | - | - | - | - | - | - | - | - |

SO, slow oscillation

### Table S19 – Effect of chronic BZRA use on SOs and spindles, their co-occurrence and SO-sigma PAC during NREM

| Outcome measure | GS | | INS | | BZRA | | GS vs INS vs MED Kruskal-Wallis test or ANOVA | | | | | GS vs INS post hoc - Dunn test or t test | | | effect size | INS vs MED post hoc - Dunn test or t test | | | effect size | GS vs MED post hoc - Dunn test or t test | | | effect size |
|---|---|---|---|---|---|---|---|---|---|---|---|---|---|---|---|---|---|---|---|---|---|---|---|
| | Mean | SD | Mean | SD | Mean | SD | F | df | residual | *p* | *q* | *t* | *p* | *q* | g' | *t* | *p* | *q* | g' | *t* | *p* | *q* | g' |
| ***Spindle characteristics*** | | | | | | | | | | | | | | | | | | | | | | | |
| **Fz** | | | | | | | | | | | | | | | | | | | | | | | |
| Density (Nber/30 sec) | 1.19 | 0.22 | 1.1 | 0.19 | 1.26 | 0.22 | *4.3* | 2 | 75 | ***0.017*** | 0.07 | 1.52 | 0.13 | 0.39 | 0.42 | -1.31 | 0.007 | **0.021*** | -0.78 | -1.31 | 0.19 | 0.58 | -0.34 |
| Duration (sec) | 0.77 | 0.04 | 0.75 | 0.03 | 0.76 | 0.05 | 3.06 | 2 | 75 | 0.21 | - | - | - | - | - | - | - | - | - | - | - | - | - |
| Amplitude (µV) | 92.7 | 18.29 | 95.02 | 23.72 | 88.18 | 21.29 | 1.79 | 2 | 75 | 0.40 | - | - | - | - | - | - | - | - | - | - | - | - | - |
| Frequency (Hz) | 11.36 | 0.5 | 11.26 | 0.5 | 11.33 | 0.44 | *0.28* | 2 | 75 | *0.75* | - | - | - | - | - | - | - | - | - | - | - | - | - |
| **Cz** | | | | | | | | | | | | | | | | | | | | | | | |
| Density (Nber/30 sec) | 1.26 | 0.24 | 1.16 | 0.31 | 1.27 | 0.23 | 3.15 | 2 | 75 | 0.20 | - | - | - | - | - | - | - | - | - | - | - | - | - |
| Duration (sec) | 0.73 | 0.04 | 0.73 | 0.04 | 0.73 | 0.04 | 0.67 | 2 | 75 | 0.71 | - | - | - | - | - | - | - | - | - | - | - | - | - |
| Amplitude (µV) | 84.78 | 17.9 | 109.35 | 72.44 | 79.89 | 14.12 | 4.90 | 2 | 75 | 0.08 | - | - | - | - | - | - | - | - | - | - | - | - | - |
| Frequency (Hz) | 13.21 | 0.43 | 13.26 | 0.58 | 13.09 | 0.48 | 2.42 | 2 | 75 | 0.29 | - | - | - | - | - | - | - | - | - | - | - | - | - |
| **Pz** | | | | | | | | | | | | | | | | | | | | | | | |
| Density (Nber/30 sec) | 1.45 | 0.27 | 1.36 | 0.32 | 1.42 | 0.32 | *0.59* | 2 | 75 | *0.55* | - | - | - | - | - | - | - | - | - | - | - | - | - |
| Duration (sec) | 0.74 | 0.04 | 0.73 | 0.04 | 0.74 | 0.05 | *0.38* | 2 | 75 | *0.68* | - | - | - | - | - | - | - | - | - | - | - | - | - |
| Amplitude (µV) | 71.25 | 17.26 | 76.91 | 22.06 | 67.7 | 12.86 | *1.04* | 2 | 75 | *0.36* | - | - | - | - | - | - | - | - | - | - | - | - | - |
| Frequency (Hz) | 13.53 | 0.56 | 13.82 | 0.47 | 13.45 | 0.52 | 7.84 | 2 | 75 | **0.019** | 0.08 | -2.1 | **0.035** | 0.10 | -0.55 | 2.67 | 0.008 | **0.02*** | 0.73 | 0.62 | 0.53 | - | 0.14 |
| ***SO characteristics*** | | | | | | | | | | | | | | | | | | | | | | | |
| **Fz** | | | | | | | | | | | | | | | | | | | | | | | |
| Density (Nber/30 sec) | 3.83 | 0.78 | 3.99 | 0.84 | 3.83 | 0.8 | *0.26* | 2 | 75 | *0.76* | - | - | - | - | - | - | - | - | - | - | - | - | - |
| Duration (sec) | 1.35 | 0.06 | 1.36 | 0.05 | 1.39 | 0.08 | 2.40 | 2 | 75 | 0.30 | - | - | - | - | - | - | - | - | - | - | - | - | - |
| Amplitude (µV) | 122.26 | 30.17 | 124.15 | 37.78 | 111.61 | 26 | *0.31* | 2 | 75 | *0.76* | - | - | - | - | - | - | - | - | - | - | - | - | - |
| Frequency (Hz) | 0.76 | 0.06 | 0.74 | 0.05 | 0.72 | 0.07 | 5.917 | 2 | 75 | 0.052 | - | - | - | - | - | - | - | - | - | - | - | - | - |
| **Cz** | | | | | | | | | | | | | | | | | | | | | | | |
| Density (Nber/30 sec) | 3.61 | 0.73 | 3.64 | 0.86 | 3.69 | 0.75 | 0.30 | 2 | 75 | 0.85 | - | - | - | - | - | - | - | - | - | - | - | - | - |
| Duration (sec) | 1.4 | 0.06 | 1.4 | 0.06 | 1.41 | 0.08 | *0.01* | 2 | 75 | *0.98* | - | - | - | - | - | - | - | - | - | - | - | - | - |
| Amplitude (µV) | 113.86 | 27.74 | 128.67 | 47.78 | 103.02 | 19 | 4.91 | 2 | 75 | 0.08 | - | - | - | - | - | - | - | - | - | - | - | - | - |
| Frequency (Hz) | 0.71 | 0.06 | 0.7 | 0.05 | 0.69 | 0.07 | *0.26* | 2 | 75 | *0.77* | - | - | - | - | - | - | - | - | - | - | - | - | - |
| **Pz** | | | | | | | | | | | | | | | | | | | | | | | |
| Density (Nber/30 sec) | 3.41 | 0.73 | 3.64 | 0.87 | 3.52 | 0.71 | 1.61 | 2 | 75 | 0.44 | - | - | - | - | - | - | - | - | - | - | - | - | - |
| Duration (sec) | 1.43 | 0.07 | 1.42 | 0.06 | 1.44 | 0.07 | 1.01 | 2 | 75 | 0.60 | - | - | - | - | - | - | - | - | - | - | - | - | - |
| Amplitude (µV) | 96.2 | 26.29 | 102.58 | 32.83 | 88.03 | 17.62 | 2.27 | 2 | 75 | 0.32 | - | - | - | - | - | - | - | - | - | - | - | - | - |
| Frequency (Hz) | 0.68 | 0.06 | 0.68 | 0.06 | 0.67 | 0.07 | 0.55 | 2 | 75 | 0.76 | - | - | - | - | - | - | - | - | - | - | - | - | - |
| ***Temporal association*** | | | | | | | | | | | | | | | | | | | | | | | |
| **Fz** | | | | | | | | | | | | | | | | | | | | | | | |
| Recall | 0.12 | 0.03 | 0.11 | 0.03 | 0.12 | 0.04 | *2.09* | 2 | 75 | *0.13* | - | - | - | - | - | - | - | - | - | - | - | - | - |
| Precision | 0.39 | 0.1 | 0.38 | 0.08 | 0.36 | 0.12 | *0.6* | 2 | 75 | *0.55* | - | - | - | - | - | - | - | - | - | - | - | - | - |
| **Cz** | | | | | | | | | | | | | | | | | | | | | | | |
| Recall (SO') | 0.1 | 0.02 | 0.09 | 0.03 | 0.09 | 0.02 | 7.41 | 2 | 75 | 0.02 | **0.049*** | 1.93 | 0.054 | - | 0.34 | 0.65 | 0.51 | - | 0.27 | 2.61 | 0.009 | **0.027*** | 0.65 |
| Precision (Spindle') | 0.29 | 0.06 | 0.3 | 0.09 | 0.26 | 0.08 | 3.36 | 2 | 75 | 0.18 | - | - | - | - | - | - | - | - | - | - | - | - | - |
| **Pz** | | | | | | | | | | | | | | | | | | | | | | | |
| Recall (SO') | 0.1 | 0.02 | 0.09 | 0.03 | 0.08 | 0.02 | 10.34 | 2 | 75 | **0.01*** | - | 1.9 | 0.06 | 0.17 | 0.45 | 1.23 | 0.22 | 0.66 | 0.36 | 3.19 | 0.002 | **0.004*** | 0.89 |
| Precision (Spindle') | 0.24 | 0.05 | 0.24 | 0.07 | 0.21 | 0.07 | *1.66* | 2 | 75 | *0.19* | - | - | - | - | - | - | - | - | - | - | - | - | - |
| ***SO-sigma PAC*** | | | | | | | | | | | | | | | | | | | | | | | |
| **Fz** | | | | | | | | | | | | | | | | | | | | | | | |
| Modulation Index | 1.63 | 1.12 | 1.22 | 1.34 | 1.18 | 1.09 | 3.42 | 2 | 75 | 0.18 | - | - | - | - | - | - | - | - | - | - | - | - | - |
| **Cz** | | | | | | | | | | | | | | | | | | | | | | | |
| Modulation Index | 2.75 | 1.33 | 2.41 | 1.47 | 1.7 | 1.09 | 8.70 | 2 | 75 | **0.01*** | - | 1.11 | 0.26 | - | 0.24 | 1.76 | 0.07 | - | 0.54 | 2.93 | 0.003 | **0.01*** | 0.84 |
| **Pz** | | | | | | | | | | | | | | | | | | | | | | | |
| Modulation Index | 3.52 | 1.62 | 2.69 | 1.59 | 2.16 | 1.43 | 10.42 | 2 | 75 | **0.006*** | - | 1.84 | 0.06 | - | 0.51 | 1.31 | 0.19 | - | 0.35 | 3.21 | 0.001 | **0.004*** | 0.88 |

SO, slow oscillation; PAC, phase-amplitude coupling

#### Chronic use of BZRA alters sleep quality

Additional analyses were carried out to balance age between groups and also evaluate the impact of BZRA on sleep quality. This process involved the comparison of the two same GS and INS groups, and a third composed of individuals exclusively using BZRA medication (BZRA group).

We found a group effect on sleep architecture measures, such as N1 ($F$(2,75)=34.7, $p$<.001), N2 ($F$(2,75)=29.8, $p$<.001), N3 ($F$(2,75)=25.5, $p$<.001), and wake duration ($F$(2,75)=15.2, $p$<.001, $q$=.001), WASO ($F$(2,75)=15.6, $p$<.001, $q$=.001) and SE ($F$(2,75)=15.2, $p$=.002, $q$=.001). Post-hoc tests showed that both INS ($p$<.001) and BZRA ($p$=.008, $q$=.025) groups presented lower SE compared to the GS group. Both INS (all $p$<.001) and BZRA (all $p$=.005, all $q$=.01) group displayed greater wake and WASO duration compared to the GS group. Participants taking sedative-hypnotics displayed greater N1 duration compared to both GS ($p$<.001) and INS ($p$<.001) groups. When compared to the GS group, we observed greater N2 duration in the BZRA ($p$=.01, $q$=.03) group, and lower in the INS ($p$<.001) group. N3 duration was lower in both GS ($p$<.001) and INS ($p$<.001) groups compared to the BZRA group. We observed a group effect driven by the INS group, where they displayed greater arousal density in both NREM and REM ($F$(2,75)=9.68, $p$=.008, $q$=.01) compared to both GS ($p$=.01, $q$=.03) and BZRA ($p$=.005, $q$=.02) groups. We also found a group effect on the SFI ($F$(2,75)=19.33, $p$<.001, $q$=.001), where the GS group displayed lower SFI compared to the BZRA ($p$=.02, $q$=.05) group, as well as greater SFI compared to the INS ($p$=.035, $q$=.10) group.

During NREM, we found a group effects in frontal region for relative spectral power in sigma ($F$(2,75)=6.5, $p$=.04, $q$=.10), low beta ($F$(2,75)=8.02, $p$=.02, $q$=.11) and high beta ($F$(2,75)=6.3, $p$=.04, $q$=.12), although it did not pass multiple comparison correction. Post-hoc tests showed that the GS group ($p$=.01, $q$=.03) presented lower low beta relative power compared to the BZRA group. Both the GS (all $p$<.04, all $q$<.06) and the INS (all $p$<.02, all $q$<.06) groups displayed lower sigma and high beta relative power compared to the BZRA group, although it did not pass multiple comparison correction. We also found a group effect on theta-beta ratio in both frontal ($F$(2,75)=12.65, $p$=.001, $q$=.01) and parietal ($F$(2,75)=13.72, $p$=.001, $q$=.01) regions, driven by the BZRA group. Post-hoc test showed that the BZRA group displayed lower theta-beta ratio compared to both GS (all $p$<.001, all $q$<.002) and INS (all $p$<.02, all $q$<.05) groups. Additionally, we found a group effect on delta-beta ratio ($F$(2,75)=6.95, $p$=.03, $q$=.11) driven by the BZRA group, although it did not pass multiple comparison correction. Delta-theta beta ratio was found lower in the BZRA group compared to the INS ($p$=.02, $q$=.05) group.

We found a group effect on spindle density ($F$(2,75)=4.3, $p$=.02, $q$=.07) in frontal and frequency ($F$(2,75)=7.8, $p$=.02, $q$=.08) in the parietal region driven by the BZRA group, although it did not pass multiple comparison. The BZRA group shown greater density compared to the INS ($p$=.007, $q$=.02) group, and lower frequency compared to the INS ($p$=.008, $q$=.02) and the GS groups before adjusted ($p$=.035, $q$=.1). SO characteristics were non significantly different between the three groups. Furthermore, we found a group effect on the temporal co-occurrence of SO and spindle ($F$(2,75)=10.34, $p$=.01), and on their phase-amplitude coupling strength (MI) ($F$(2,75)=10.42, $p$=.006) solely in the parietal region, which where both lower in the BZRA group compared to the GS (all $p$<.002, all $q$=.004) group.
